# Asyndromic Surveillance of New York City Emergency Department Diagnoses with the Tree-Temporal Scan Statistic

**DOI:** 10.1101/2025.11.11.25339953

**Authors:** Sharon K. Greene, Alison Levin-Rector, Martin Kulldorff, Ramona Lall

## Abstract

**Objectives:** Illness trends are typically monitored by reportable disease and syndromic surveillance systems, but unanticipated health issues might not be captured. Using diagnosis codes, the New York City Health Department developed a data mining method to detect unusual increases in emergency department (ED) visits for any reason.

**Methods:** We applied the tree-temporal scan statistic in TreeScan software to ICD-10-CM diagnosis codes for ED visits. We searched for unusual citywide increases in ED visits or hospital admissions, over any recent time period, and at any part of and level on the ICD-10-CM tree. We conducted proof-of-concept analyses for March 2020 when COVID-19 emerged, then investigated signals detected in daily, automated analyses during April–August 2025.

*Results:* If TreeScan analyses had been in place, then increasing hospital admissions for viral pneumonia (J12) would have triggered a signal on March 13, 2020, two days before widespread COVID-19 community transmission was announced. An extreme heat event in June 2025 triggered a signal for admissions for acute kidney failure (N17), prompting outreach to dialysis networks. A sustained signal for hand, foot, and mouth disease (B08.4) prompted outreach to child care programs. Other signals supported situational awareness, including a seasonal increase for swimmer’s ear (H60.33) and burns (T30.0) related to consumer fireworks.

*Practice Implications:* TreeScan quickly detected credible increases in various diagnoses without pre-specification, from minor to severe, rare to common, acute to sustained, and foreseen to unforeseen. TreeScan can strengthen surveillance for health issues related to new pathogens, non-notifiable conditions, environmental exposures, and mass gatherings.

---

To detect and monitor outbreaks and other conditions of public health importance, the New York City (NYC) Health Department receives near-real time data on reportable diseases^1^ and emergency department (ED) visits.^2^ To conduct syndromic surveillance, syndromes like influenza-like illness or heat-related illness are defined using ED diagnoses and chief complaint terms.^3^ While useful for public health response and decision-making,^2^ reportable disease and syndromic surveillance systems have limitations. If a new health issue arises that is not a reportable disease or predefined syndrome, then any chance for a public health response might be delayed. Illnesses due to novel pathogens might not be captured, as before SARS-CoV-2 testing became accessible^4,5^ and new diagnostic codes were introduced and incorporated into a COVID-19–like illness syndrome.^6^ Syndromes might need updating as clinical knowledge improves, such as removing a fever requirement from a COVID-19–like illness syndrome definition.^7^ In prioritizing specificity over sensitivity, definitions might be narrow. For instance, a heat-related illness syndrome relies on clinicians to code visits as directly heat-related, such as for heat exhaustion or sun stroke,^8,9^ and omits indirect effects, such as dehydration or exacerbation of kidney disease.^10^ Classifying some ED visits into syndromes and ignoring all other visits discards potentially informative data, limiting visibility into real-time population trends. In contrast, the goal of asyndromic surveillance is to search for any unusual increases in illness data, without prespecifying diseases or syndromes of concern.^11,12^

Previously introduced asyndromic surveillance methods have substantial limitations. The first such method implemented by the NYC Health Department, in 2013, detected relative increases in chief complaint keywords.^2,13^ Such word alerts tend to preferentially detect obscure terms and can fail to group related visits with different keywords due to typos, abbreviations, and synonyms.^12–15^ Although improvements using natural language processing, generative topic modeling, and machine learning have been proposed,^16–19^ their complexity and opacity can impede adoption by public health authorities. Another approach entails assessing weekly trends in *International Classification of Diseases, Tenth Revision, Clinical Modification* (ICD-10-CM) diagnosis codes.^20,21^ Treating every ICD-10-CM code separately fails to leverage their clinically meaningful, nested structure. Additionally, using fixed time windows fails to account for different types of health issues emerging abruptly or gradually over different time frames.

To detect emerging health issues that might not be captured by reportable disease or syndromic surveillance, we used the tree-temporal scan statistic^22,23^ for asyndromic surveillance to simultaneously monitor thousands of ICD-10-CM codes and related code groupings, over varying time frames, while adjusting for the multiplicity of codes and time frames evaluated. A similar approach is used in pharmacovigilance to detect diagnoses indicating possible adverse events, with the distinction of scanning over person-time since vaccine or drug administration^24,25^ rather than scanning over calendar time. We report results of daily, prospective analyses of NYC ED data, first as mimicked for proof-of-concept for March 2020 when COVID-19 emerged, and then as applied in real-time during April–August 2025.

## Methods

### Emergency Department Data

The NYC Health Code authorizes the Health Department to collect ED visit data from NYC hospitals.^26^ These data include an anonymized patient identifier, demographic characteristics, visit date, disposition, and ICD-10-CM diagnosis codes.^26,27^ From the latter field, we excluded any ED-provided codes other than ICD-10-CM codes, such as SNOMED and CPT codes, and standardized by removing extra spaces and decimal points. We determined all diagnoses per patient per visit that were eligible (acute health issues of interest), incident (first diagnosis per ICD-10-CM tree subchapter within a 1-year period), and unique (one diagnosis per subchapter per day) (eMethods 1 in the Supplement).

A daily input file for the past 90 days consisted of three columns: (1) a hyphen-delimited concatenation of hospital inpatient admission status (1 = admitted; 0 = not admitted) and ICD-10-CM code, (2) visit date, and (3) the citywide count of patients with that admission status and ICD-10-CM diagnosis on that date.

The NYC Health Department Institutional Review Board determined this study (#24-084) was exempt human subjects research under 45 CFR §46.104 (d)(4)(iii) because it involved secondary use of clinical data. We followed reporting guidelines for studies using observational routinely-collected health data.^28^

### Tree File

A tree file consisted of every eligible ICD-10-CM code and its parents (eAppendix 1 in the Supplement). ICD-10-CM codes are grouped in a hierarchical tree structure, reflecting general or specific disease conditions affecting different body systems, with related diagnoses on the same tree branch. We downloaded the tree structure provided by the Centers for Medicare & Medicaid Services.^29^

We assigned three nodes to every ICD-10-CM code, distinguished by whether the patient was admitted (“1-” prefix) versus not admitted (“0-” prefix), as well as a parent node (“2-” prefix) indicating any ED visit for that diagnosis, regardless of admission status. This allowed us to simultaneously evaluate diagnoses for all ED visits (“2-” prefix) and for admissions only (“1-” prefix), without also evaluating diagnoses for non-admissions only (“0-” prefix).

We added supplemental nodes connecting ICD-10-CM codes on different parts of the tree sharing a common etiology (eAppendix 1 in the Supplement). When infections increase for a particular pathogen, for example, *Mycoplasma pneumoniae*, some patients might have pneumonia (J12-J18) while others might have acute bronchitis (J20) or another illness manifestation.^30,31^ To improve the signal-to-noise ratio to quickly detect outbreaks, we added pathogen-specific supplemental nodes to scan not only for increases affecting a common body system on the same tree branch, but also for increases in illnesses caused by a common pathogen (Figure 1).

**Figure 1.**
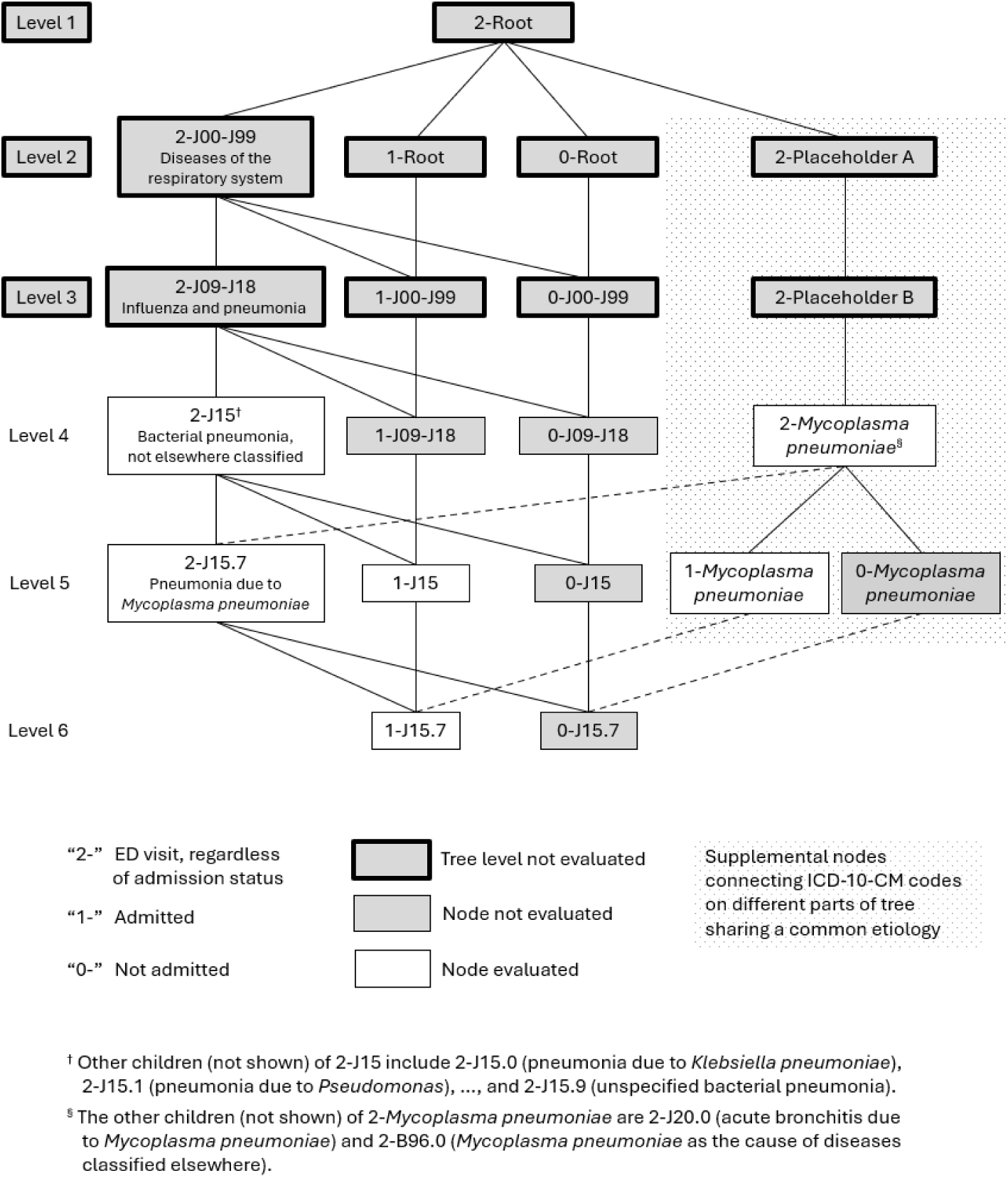
Excerpt of an International Classification of Diseases, Tenth Revision, Clinical Modification (ICD-10-CM)-based tree structure for an emergency department diagnosis of J15.7 (pneumonia due to *Mycoplasma pneumoniae*). In tree level 6, patients are classified as admitted (“1-” prefix) or not admitted (“0-” prefix). Both nodes have 3 parents in level 5. First, both nodes share the parent of ED visits for J15.7 regardless of admission status (“2-” prefix). Second, a parent for 1-J15.7 is 1-J15 (admissions for bacterial pneumonia, not elsewhere classified); correspondingly, a parent for 0-J15.7 is 0-J15. Third, a parent for 1-J15.7 is 1-*Mycoplasma pneumoniae* (admissions for *Mycoplasma pneumoniae*); correspondingly, a parent for 0-J15.7 is 0-*Mycoplasma pneumoniae.* Shaded nodes are not evaluated because either the entire tree level is not evaluated (too general) or the individual node is not evaluated (not of interest). By applying the tree-based scan statistic to this structure, we simultaneously searched for increases in any of the 6 unshaded nodes, i.e., pneumonia due to *Mycoplasma pneumoniae*; bacterial pneumonia, not elsewhere classified; and *Mycoplasma pneumoniae* infections affecting any organ system — each in hospital admissions only and ED visits overall.

### Tree-Temporal Scan Statistic

We conducted prospective analyses with the tree-temporal scan statistic^22,23,32^ (Table 1) using the TreeScan^TM^ software v2.3.^33^ We flexibly searched for unusual increases in diagnoses emerging over any length of recent time and at any part of and level on the ICD-10-CM tree, from general to specific diagnoses. Under the null hypothesis, the proportion of each diagnosis among all ED visits and hospital admissions for a given day of the week was constant over time. Under the alternative hypothesis, for one diagnosis or group of related diagnoses, its proportion among all ED visits or hospital admissions was higher during some recent period.

**Table 1.** Specifications for asyndromic anomaly detection using the tree-temporal scan statistic applied to New York City emergency department diagnosis codes.

| Feature | Selection | Notes |
| --- | --- | --- |
| Source population | Persons in NYC on any day during the time range. | We used an open population instead of a closed cohort to detect emergent health issues affecting anyone physically present in NYC, including residents and visitors. |
| Database population | Persons from the source population who presented for care at any reporting ED. | ED data provide comprehensive source population coverage, reflecting visits for any reason. Patients who are low-income or under-insured might be under-represented in other health data sources while disproportionately accessing EDs for primary care services.<sup>72</sup><br>As of August 2025, 49 EDs were included. The 3 remaining NYC EDs, which were administered by the U.S. Department of Veterans Affairs, were excluded because they did not provide ICD-10-CM codes.<br>Occasionally, file transmissions from a given ED to the NYC Health Department might be interrupted by technical issues. An ED was temporarily considered “non-reporting” and omitted from daily analysis if the hospital reported 0 ED visits for the most recent $\geq 2$ days of the time range. This data cleaning step assured that until any technical issues were resolved, EDs did not misleadingly weaken or strengthen emerging signals by contributing data to the baseline but not the cluster period. |
| Study population | Persons from the database population with an eligible, incident, and unique diagnosis. | One investigator (R.L.) had unrestricted access to the database population and generated input files daily.<br>Diagnoses received within 14 days of the ED visit were included. ED visits with no diagnosis were excluded. |
|  |  | This occurred, for example, when patients left the ED against medical advice. |
| Visit severity | Admitted (yes/no). | It is of considerable interest to detect serious, emerging health issues resulting in hospitalization. Using the discharge disposition field, we distinguished diagnoses of ED patients who were “admitted as an inpatient to this hospital” or “expired”<sup>27</sup> (i.e., died) (“1-” prefix) versus not admitted (“0-” prefix).<br>Of 49 included EDs, 26 (53%) had a data quality issue as of August 2025 in which the disposition field was blank instead of populated as admitted. As a data cleaning step until this issue can be corrected, any visit at these 26 EDs with a missing discharge disposition code as of 2 days after the visit was imputed as admitted. |
| Time precision | Day | We used daily resolution for precise cluster start dates. |
| Data time range of ED visits | Rolling 90-day period, ending day prior to analysis | The data time range should be at least 3 times as long as the maximum temporal size (specified below).<sup>49</sup><br>Analogously, case-control studies have diminishing returns in statistical power with control-to-case ratios above 3.<sup>73</sup> By keeping the time range short, the cluster and baseline periods were more comparable and robust to administrative changes in input data, such as when a hospital adopts new ICD-10-CM codes.<br>After a major health event, to avoid difficulty in detecting subsequent increases in the same diagnoses, we excluded from the baseline period all ED visits citywide during the days of the major health event. For example, starting July 23, 2025, we excluded from the baseline period all ED visits during a spike in heat-related illness (June 23–June 26, 2025) to preserve the ability to detect health effects from subsequent heat waves. |
| Only allow data on leaves of tree | No | We used ICD-10-CM codes as provided by EDs, without requiring codes to be at the most specific node. |
| Allow multiple root nodes | No | The ICD-10-CM tree had only one root, which is a node without a parent. |
| Allow multiple parents for the same node | Yes | We allowed multiple parents per node to: (a) nest every node for a diagnosis resulting in hospital admission within a node for all ED visits for that diagnosis, and (b) connect diagnoses on different parts of the tree sharing a common etiology. |
| Tree levels excluded from evaluation | 1, 2, 3 | All diagnoses in these tree levels were excluded from evaluation as too broad to be actionable (Figure 1). |
| Specify separate file with nodes not to evaluate | Yes | We specified certain nodes in tree levels $\geq 4$ not to evaluate (eAppendix 2 in the Supplement). Nodes were included in input files but not evaluated if: (a) diagnoses were for the subset of ED patients who were not admitted (“0-” prefix), because we were only interested in signals of diagnoses for ED patients overall (“2-” prefix) or the subset admitted (“1-” prefix), or (b) diagnoses were too specific to plausibly reflect population-level emergent health issues, such as diagnoses distinguishing whether a health issue was for the left, right, bilateral, or unspecified sides of the body. Such diagnoses were rolled up and evaluated only at less specific nodes.<br>Additionally, when new ICD-10-CM codes were added annually in October,<sup>29</sup> we incorporated the new codes into the tree file for system maintenance and temporarily did not evaluate them until the data time range of 90 days (specified above) had elapsed since adoption. This step assured that we did not get false signals due to the introduction of new codes that were not present during the baseline period. |
| Type of scan | Tree and time | We scanned for increases in ED visits for any diagnosis or group of related diagnoses and over any recent time period. |
| Temporal size | Minimum = 1 day.<br>Maximum = 28 days. | We searched for diagnoses with increases during the most recent 1, 2, 3, ..., 27, or 28 days. Our goal was to detect not only health issues emerging abruptly, as is the focus of ESSENCE alerting algorithms,<sup>74</sup> but also issues emerging stealthily over a few weeks, such as due to intermittent or prolonged source exposures, person-to-person transmission, or care-seeking delays. More slowly emerging health issues would be more difficult for health care personnel to notice manually and are thus an important use case for data mining methods. |
| Conditional analysis | Node and time | We conditioned on time, representing the ED visit date, to adjust nonparametrically for any citywide purely temporal patterns. These patterns include data lags, reduced ED volume on weekends and holidays, and any increasing or decreasing secular trends in ED visits, such as periods when virtual visits with health care providers were encouraged.<sup>75</sup><br>We also conditioned on node, representing the ICD-10-CM code, to account for whether each diagnosis had been common or rare during the baseline period. This allowed us to detect diagnoses with abnormal increases and not just the most common diagnoses throughout the time range. |
| Perform node by day-of-week adjustment | Yes | We adjusted for diagnosis by day-of-week interaction to account for variation in the distribution of ICD-10-CM codes by day-of-week of ED visit. This accounts for some diagnoses being approximately equally common on all days of the week (e.g., A09 [infectious gastroenteritis and colitis, unspecified]), while other |
|  |  | diagnoses are more common on weekends (e.g., W34 [accidental discharge and malfunction from other and unspecified firearms and guns]).<br>With this adjustment, the dates in which ED visits occurred were shuffled, restricting to the same day of the week, and assigned to the original ICD-10-CM codes. The 90-day time range included several baseline weeks, avoiding unstable day-of-week-specific estimates. |
| Scan for branches with: | High rates | Our goal was to detect when diagnoses became an increasing percentage of ED visits. One could also scan for diagnoses that became less common. |
| Prospective evaluation | Yes | We used prospective analyses to only consider temporal windows reaching through the end of the time range. We were only interested in detecting current problems, not historical health issues that occurred some weeks or months ago. |
| Monte Carlo replications | 99,999 | We used Monte Carlo hypothesis testing to assess the statistical strength of clusters, controlling for the multiplicity of overlapping diagnoses and time windows evaluated.<br>This number of replications can yield sufficiently large recurrence intervals to distinguish weaker and stronger signals (specified below) while maintaining reasonable computing time, as determined by the number of tree nodes and time intervals evaluated. |
| Inference method | Sequential Monte Carlo, with early termination cutoff of 5,000 | To save computational time, we allowed runs to terminate early if no potential clusters were detected after >5% of the total replications. |
| Prospective analysis frequency | Daily at 10 a.m. | We ran daily analyses to quickly detect emergent health concerns. |
|  |  | Scheduling analyses to run at 10 a.m. allowed reporting to plateau for the prior day's ED visits, while also allowing same-day results review and signal escalation as warranted. |
| Minimum number of cases | 3 | We selected a low minimum so we could detect increases in rare diagnoses. |
| Signal definition | Recurrence interval (RI) $\geq 365$ days and relative risk (RR) $\geq 1.3$ | We considered RI $\geq 365$ days to RI $< 5$ years as moderate strength, RI $\geq 5$ years to RI $< 100$ years as strong, and RI $\geq 100$ years as very strong.<sup>49</sup> We used RIs instead of *P*-values. Applying the typical threshold of $P < 0.05$ would not adequately control type I error in prospective analyses as it would, on average, generate a false signal every 20 days.<sup>49</sup><br>We ignored nodes with only slightly elevated RR because with a large volume of ED visits, clusters can have high RI but with a RR close to 1 that is not meaningful from a public health perspective. |
| Signal classification | New, increasing, maximum, stable, or decreasing | In daily analyses, the set of emerging diagnoses can be very similar from one day to the next. To focus attention on new and emergent health issues and minimize signal fatigue, we compared signals for today versus prior days and classified as follows:<br>*New*: Yesterday's RI $< 365$, RR $< 1.3$, or node missing.<br>(Over 149 days of analyses conducted during April 5–August 31, 2025, the number of new signals per day was 0 for 74 days [50%], 1 for 40 days [27%], 2–3 for 26 days [17%], and 4–6 for 9 days [6%.])<br>*Increasing*: If signal persists 2 or 3 days, then RI monotonically increasing. If signal persists $\geq 4$ days, then slope of trendline for RI by day is positive and $P < 0.05$. |
|  |  | *Maximum:* RI = 100,000 days today and yesterday, as determined by the number of Monte Carlo replications (99,999).<br>*Stable:* If signal persists 2 or 3 days, then RI neither monotonically increasing nor decreasing. If signal persists $\geq 4$ days, then slope of trendline for RI by day is $P \geq 0.05$.<br>*Decreasing:* If signal persists 2 or 3 days, then RI monotonically decreasing. If signal persists $\geq 4$ days, then slope of trendline for RI by day is negative and $P < 0.05$. |

Abbreviations: ED, emergency department; ICD-10-CM, *International Classification of Diseases, Tenth Revision, Clinical Modification*; NYC, New York City; RI, recurrence interval; RR, relative risk.

We used Monte Carlo hypothesis testing to control overall type I error across the multiplicity of overlapping sets of diagnoses and time windows evaluated on any analysis day. Statistical details are in the TreeScan user guide.^34^ We defined signals based on relative risk and, to account for the daily analytic cadence, the recurrence interval (RI = 1 / *P*-value). The RI represented the duration of daily surveillance required for the expected number of clusters at least as unusual as the observed cluster to be equal to 1 by chance.^35^ We tracked RI trends as signals strengthened, held steady, and weakened, mirroring epidemic patterns (Table 1).

### Proof-of-Concept: COVID-19 Emergence

We mimicked daily surveillance for February 29, 2020 (when the first laboratory-confirmed COVID-19 case was diagnosed in NYC^5^ and prior to which no increase in COVID-19–like illness syndrome in EDs was observed^36^) through March 15, 2020 (when widespread community transmission was announced^37^). SARS-CoV-2 testing had been restricted at public health laboratories and not yet commercially available, curtailing situational awareness.^5^ We assessed if and when increases in COVID-19–related diagnoses could have been detected by our new approach had it been applied at the time.

### Real-Time Prospective Analyses

We launched daily, automated analyses on April 4, 2025. When interpreting each signaling diagnosis, we reviewed summary data, including weekly trends for the most recent four years to assess seasonality; frequent co-diagnoses; and differences between baseline and cluster periods in patient demographic characteristics (sex, age, race, ethnicity, and borough of residence) and in the distribution of EDs to which patients presented. When patients disproportionately presented to certain EDs, we determined whether the health issue was truly geographically focused versus an artifact of hospital-specific changes, such as delayed adoption of new ICD-10-CM codes. We additionally considered patient-level records, including chief complaints and triage notes. As warranted, we escalated signals to relevant subject matter experts at the NYC Health Department for situational awareness and possible public health action. We compiled illustrative examples of TreeScan signals detected during the first five months of daily analyses conducted during April–August 2025. Additionally, we investigated why TreeScan analyses did not detect a large Legionnaires’ disease outbreak in July 2025.

## Results

### Proof-of-Concept: COVID-19 Emergence

If the NYC Health Department had been using TreeScan for daily analyses of ED visit diagnoses when COVID-19 emerged, then the first signals would have been detectable on March 10, 2020 (Table 2), two days before a state of emergency declaration.^38^ Signals detected during March 10–15 by data mining and with no pre-specification reflected a breadth of diagnoses, including relevant symptoms (e.g., ICD-10-CM code R05 [cough]) and factors increasing the risk of severe complications (e.g., F17.210 [cigarette smoking], E11.9 [type 2 diabetes]). As the COVID-19–specific diagnosis code U07.1 was not effective until April 1, 2020,^39^ certain EDs instead leveraged available codes for coronavirus infections (B97.2, B34.2), as also reflected in signals for the supplemental node “other coronavirus” connecting ICD-10-CM codes on different parts of the tree sharing a common etiology. Hospital admissions for J12 (viral pneumonia) signaled on March 13, indicating severe illness. By the time widespread community transmission was announced on March 15,^37^ rapid increases in both rare and common diagnoses would have been readily apparent (eFigure 1 in the Supplement).

**Table 2.**
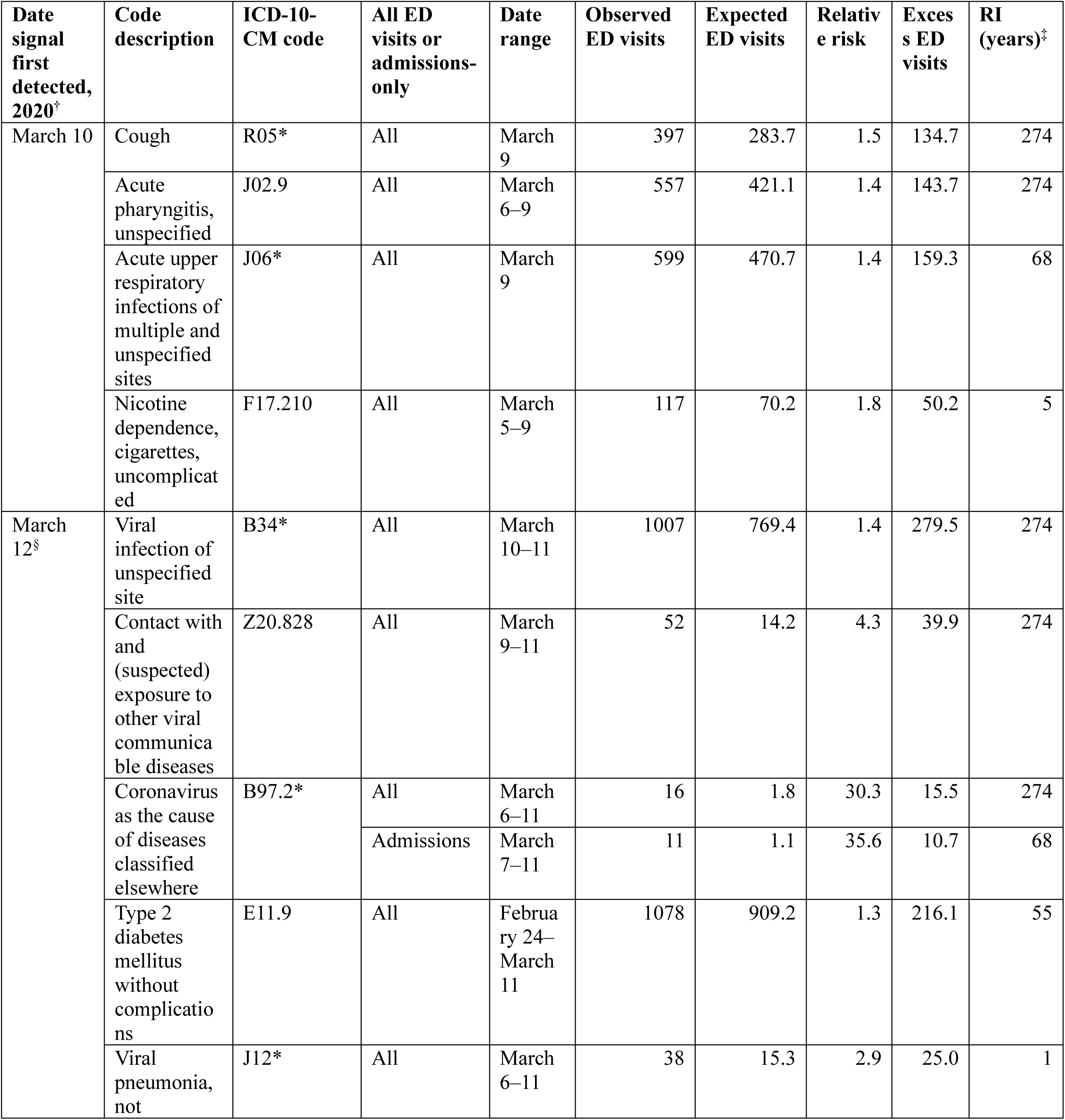
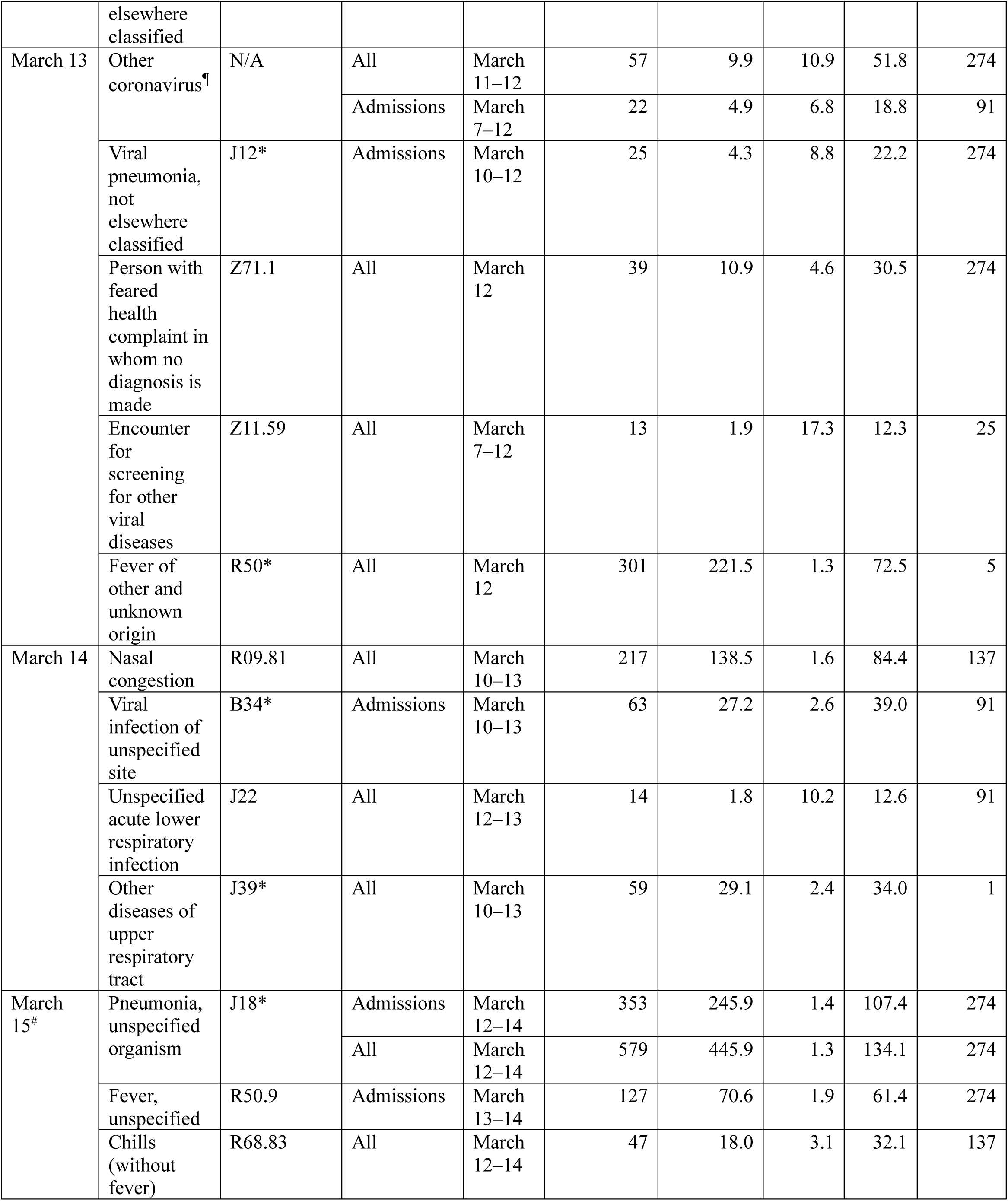
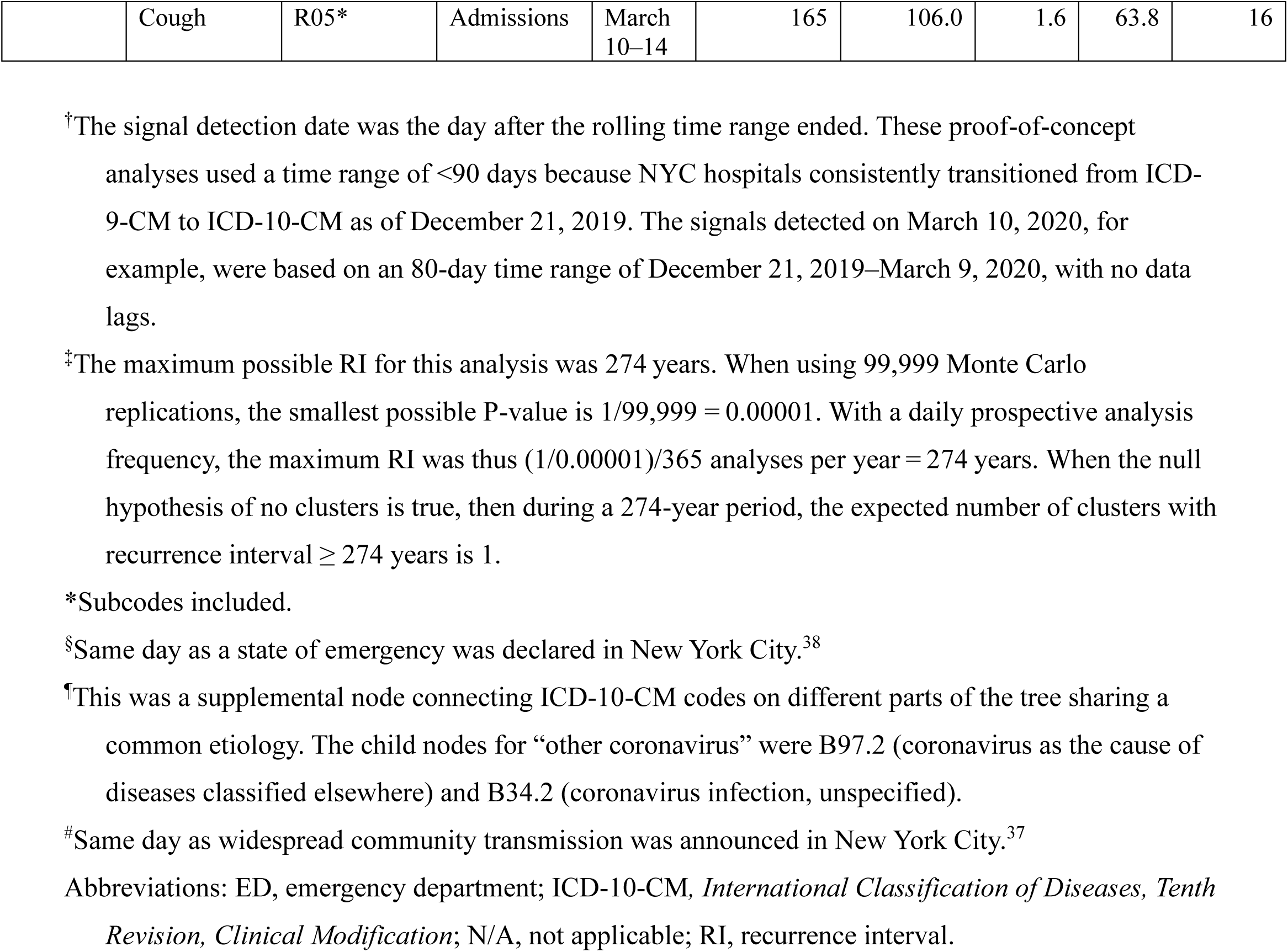
Signals for COVID-19-related diagnoses in New York City emergency departments detected by mimicking prospective daily analyses using the tree-temporal scan statistic, February 29–March 15, 2020.

### Heat-Related Illness

On May 18, 2025, TreeScan detected a spike for ICD-10-CM code T67 (effects of heat and light) on the prior day. The chief complaints indicated that patients presented with heat stroke and heat exhaustion after participating in a half marathon under unseasonably hot and humid conditions.^40^

An extreme heat event occurred during June 22–25, 2025, when the daily heat index was 95–103°F.^41,42^ ED visits for the heat-related illness syndrome^10^ spiked June 23–26.^42^ The only ICD-10-CM codes included in this syndrome definition were T67 and X30 (exposure to excessive natural heat).^10^ TreeScan quickly detected an increase in T67 on June 22, and then detected increases in several other heat-related diagnoses during June 24–29 (Table 3, eFigure 2 in the Supplement). Hospital admissions for acute kidney failure signaled on June 27, indicating severe illness. Of 1,094,275 eligible, incident, and unique diagnoses during the 90-day period ending June 28, 1,047,210 (95.7%) were available in near-real time (Figure 2), so signals were timely and actionable. In response and in advance of a subsequent heat wave, the NYC Health Department alerted dialysis networks and connected them to messaging from the NYC Health Department and NYC Emergency Management to proactively prepare patients for heat events.

**Table 3.** Signals on June 29, 2025^†^ related to the June 22–25 extreme heat event in New York City detected by applying the tree-temporal scan statistic to emergency department diagnosis data from March 31 to June 28.

| Code description | ICD-10-CM code | All ED visits or admissions -only | Date range | Observed ED visits | Expected ED visits | Relative risk | Excess ED visits | RI (years) | Days ago signal first detected |
| --- | --- | --- | --- | --- | --- | --- | --- | --- | --- |
| Effects of heat and light | T67* | All | June 22–28 | 282 | 31.6 | 24.5 | 270.5 | 274 | 7 |
|  |  | Admissions | June 22–28 | 44 | 6.0 | 14.8 | 41.0 | 274 | 4 |
| Acute kidney failure | N17* | All | June 23–28 | 519 | 344.3 | 1.6 | 190.9 | 274 | 4 |
|  |  | Admissions | June 23–28 | 322 | 233.1 | 1.4 | 97.5 | 46 | 2 |
| Volume depletion | E86* | All | June 23–28 | 281 | 159.4 | 1.9 | 131.4 | 274 | 4 |
| Miliaria rubra | L74.0 | All | June 20–28 | 46 | 11.9 | 6.6 | 39.0 | 274 | 5 |
| Syncope and collapse | R55* | All | June 19–28 | 1513 | 1285.7 | 1.2 | 261.1 | 274 | 3 |
| Other symptoms and signs involving cognitive functions and awareness | R41.8* | All | June 23–28 | 433 | 321.5 | 1.4 | 116.3 | 137 | 3 |
|  |  | Admissions | June 23–28 | 262 | 191.3 | 1.4 | 72.7 | 0.8 | 2 |
| Malaise and fatigue | R53* | All | June 23–28 | 616 | 488.5 | 1.3 | 147.8 | 46 | 2 |
| Hypo-osmolality and hyponatremia | E87.1 | All | June 21–28 | 239 | 163.7 | 1.5 | 82.2 | 46 | 3 |
| Sunburn | L55* | All | June 20–28 | 31 | 9.6 | 3.9 | 23.0 | 34 | 3 |
| Rhabdomyolysis | M62.82 | All | June 23–28 | 69 | 35.2 | 2.1 | 35.5 | 3 | 0 |

<sup>†</sup>The TreeScan analysis on this date analyzed 1,047,210 ICD-10-CM diagnoses, with a computer running time of 20 minutes 57 seconds.
\*Subcodes included.

**Figure 2.**
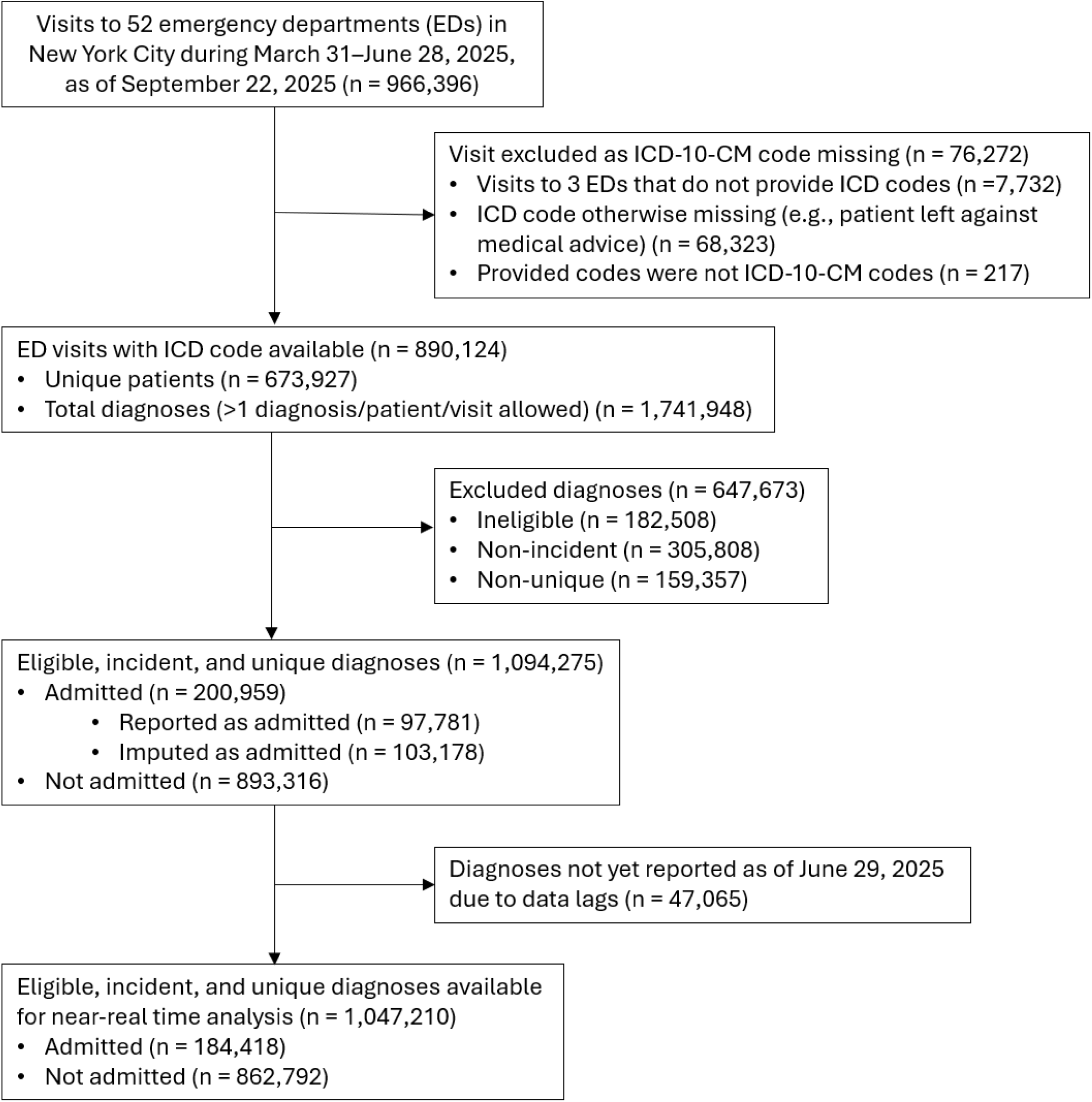
Population flow diagram for input file for analysis run on June 29, 2025.

### Hand, Foot, and Mouth Disease and Other Seasonal Increases

On April 5, 2025, TreeScan detected the start of a seasonal increase in diagnoses for B08.4 (enteroviral vesicular stomatitis with exanthem), also known as hand, foot, and mouth disease (HFMD) (eFigure 3 in the Supplement). In the United States, HFMD is a common, typically mild viral illness that is easily transmitted among young children, peaks seasonally from spring to fall, and is not notifiable to health departments.^43^ HFMD continued to signal for >4 months of daily TreeScan analyses, with the last signal on August 19. A comparison of weekly trends showed the seasonal increase in 2025 was markedly larger than that of the three prior years (eFigure 3 in the Supplement).

NYC child care providers are required to report HFMD clusters (≥3 cases within a single child care facility) to the NYC Health Department.^44^ An unusual increase in reported HFMD clusters in child care facilities independently corroborated the increase in ED diagnoses. Unusual HFMD increases were concurrently noted in multiple states.^45^ In response, the NYC Health Department disseminated HFMD messaging to all child care programs regulated under NYC Health Code Article 47 for group day care and Article 43 for school-based child care, highlighting HFMD prevention through handwashing, disinfection, and temporary exclusion of infected children.

TreeScan also detected diagnoses with seasonal increases of similar magnitude as prior years. These included L23.7 (allergic contact dermatitis due to plants, except food [commonly poison ivy exposure]), H60.33 (swimmer’s ear), and T63.44 (toxic effect of venom of bees). These signals provided situational awareness, but did not warrant public health action.

### Injuries

TreeScan detected signals on July 5, 2025 for T30.0 (burn of unspecified body region, unspecified degree) and on July 9 for W39 (discharge of firework) (eFigure 4 in the Supplement). For T30.0, patient chief complaints typically did not mention the burn source, although for three patients with no W39 co-diagnosis, “fireworks” or “firecrackers” were mentioned. The NYC Health Department issues annual press releases around Independence Day encouraging New Yorkers to avoid consumer fireworks, citing counts of fireworks-related ED visits^46^ as determined by W39 or “fireworks” in the chief complaint. During July 4–8, 2025, 35 ED visits were classified as fireworks-related, while TreeScan estimated 40.4 excess visits for T30.0, indicating W39 was likely under-coded. To lessen reliance on clinicians documenting fireworks-related visits, future public health messaging could account not only for visits directly attributed to fireworks, but also concurrent excess visits for burns.

On August 18, TreeScan detected a spike for W34.00 (accidental discharge from unspecified firearms or gun) (eFigure 4 in the Supplement). The spike was attributable to a mass shooting at a bar on August 17,^47^ demonstrating TreeScan can support situational awareness during mass casualty events.

### A missed cluster

A community cluster of Legionnaires’ disease in central Harlem was detected on July 25, 2025 by applying the space-time scan statistic to electronic laboratory reports.^48,49^ This cluster, which grew to 114 cases diagnosed during July 22–August 19, 2025,^50^ was not detected by TreeScan. Of 100 outbreak-linked patients with an ED visit, only 6 patients were diagnosed with A48.1 (Legionnaires’ disease). The most common diagnosis, with 72 patients, was J18.9 (pneumonia, unspecified organism), for which the background rate was high, with 3,269 diagnoses citywide during the outbreak period. Thus, the signal-to-noise ratio in citywide temporal analyses was low for a nonspecific diagnosis and a geographically localized outbreak.

## Discussion

By applying the tree-temporal scan statistic to ED diagnoses, we detected various emerging health issues without pre-specification that affected NYC residents and visitors. Within the first five months of daily analyses, TreeScan detected increases in diagnoses that were minor (e.g., allergic contact dermatitis due to poison ivy exposure) and severe (e.g., hospital admissions for acute kidney failure); rare in the ED (e.g., miliaria rubra [heat rash]) and common (e.g., syncope and collapse); acute (a 1-day increase in heat exhaustion from a half marathon) and sustained (a prolonged HFMD increase); and foreseen (injuries from consumer fireworks around Independence Day) and unforeseen (gunshot wounds).

Two signals prompted focused outreach separately to dialysis networks and child care programs. If TreeScan analyses had been in place during COVID-19 emergence, then policymakers might have had timelier knowledge of widespread community transmission and illness burden and severity. Factors increasing risk of severe complications, including cigarette smoking and type 2 diabetes, might have been characterized by mid-March 2020, before more resource-intensive studies could be conducted.^51,52^ When many diagnoses signaled simultaneously, as during COVID-19 emergence (Table 2) and extreme heat events (Table 3), signal definition and classification criteria (Table 1) helped focus attention on new and emerging health issues.^53^

### Limitations

Repurposing administrative ICD-10-CM codes for public health surveillance carries limitations. First, patients with no ICD-10-CM codes assigned were excluded from analysis. These patients might have left against medical advice or discontinued care for other reasons, and their missingness could reduce population representativeness^54^ and contribute to missed or delayed signal detection. Second, where assigned, ICD-10-CM codes might misclassify patients’ true health conditions.^55^ Diagnoses are assigned during ED visits and at discharge, prior to potential later adjustment by hospital billing departments, so likely reflect clinical impressions. Validating all diagnosis codes for population surveillance is impractical, but even with baseline misclassification and variation in coding processes across clinicians and institutions contributing noise,^56^ signals of increases above baseline should be detectable. Third, although we provided examples of detecting health issues the immediate day after an exposure, illnesses requiring more time to diagnose and data delays can contribute to missed or delayed signal detection. We are unaware of systematic differences in ICD-10-CM code timeliness by nature of diagnosis, hospital system, or admission status in NYC, but timeliness could vary within and across jurisdictions.^57^

While an important addition to the surveillance toolkit, TreeScan did not detect every emerging health issue in NYC, including a Legionnaires’ disease outbreak. Depending on the signal-to-noise ratio, a geographically limited outbreak might not be apparent in citywide temporal analyses, especially when patients are assigned nonspecific and common diagnoses such as for pneumonia caused by an unspecified organism. TreeScan analyses of ED visits should thus supplement and not replace other aberration detection methods, such as spatiotemporal cluster detection of reportable diseases.^49^ Health issues primarily affecting a subpopulation (e.g., <5 year-olds) might not be detected in population-wide analyses. Stratified analyses restricting to selected subpopulations of special interest could improve the signal-to-noise ratio for such issues.

### Practice Implications

A unified, practical, and timely approach for asyndromic surveillance has been heretofore elusive.^58^ By applying data mining to ED diagnoses, authorities now can automatically detect credible increases in visits for any cause, without preselection, and use findings in near-real time to quickly detect emerging health issues, launch investigations, mitigate hazards, support situational awareness, and allocate resources. The approach could be applied to any data source with ICD-10 codes, including urgent care visits^59^ and hospitalization claims,^60^ as well as cause-of-death mortality data.^61^

In addition to infectious disease outbreak detection, TreeScan can be used to detect health issues during mass gatherings, such as for sporting, political, religious, or cultural events^62,63^ and health effects of environmental exposures, including extreme heat,^64^ extreme cold,^65^ hurricanes,^66^ flooding,^67^ wildfire smoke inhalation,^68^ and power outages.^69^ TreeScan can also detect population-level increases in newly emerging pathogens (e.g., COVID-19 emergence) and in conditions not required to be reported to health departments, such as HFMD and *Mycoplasma pneumoniae* infections,^31^ which otherwise can be difficult to monitor in near-real time. With adjustments to time frames, this approach could also be useful in detecting longer-term effects of changes in health policy and services.^70,71^

## Supporting information

eAppendices

Preparing TreeScan input files

Reproducing a TreeScan analysis

## Data Availability

Sample code for generating TreeScan input files from a user's own data source is provided in the Supplement. For transparency and reproducibility of signals related to an extreme heat event, TreeScan input and output files (which contain no confidential patient data) for prospective surveillance on June 29, 2025 are provided in the Supplement. The TreeScan software (www.treescan.org) and source code (github.com/scanstatistics/treescan) are freely available.

## Acknowledgments

We gratefully acknowledge Leah Seifu, Joel Ackelsberg, and Rachel Paneth-Pollak (NYC Health Department) for contributions to real-time signal interpretation and follow-up. Judy Maro and Katherine Yih (Harvard Pilgrim Health Care Institute) provided helpful guidance for generating input files and automating TreeScan analyses. Drs. Seifu, Ackelsberg, Paneth-Pollak, and Maro also provided constructive suggestions on a previous manuscript draft. We additionally thank the Syndromic Surveillance Team (NYC Health Department) for ED data maintenance and Scott Hostovich (Information Management Services, Inc.) for incorporating updates into TreeScan software v2.3.

## Author Contributions

Greene: Conceptualization (lead), Formal analysis (supporting), Funding acquisition (lead), Investigation (supporting), Methodology (equal), Project administration (lead), Software (supporting), Supervision (lead), Writing – original draft (lead), Writing – review & editing (equal).

Levin-Rector: Data curation (supporting), Formal analysis (supporting), Methodology (supporting), Software (supporting), Writing – review & editing (supporting).

Kulldorff: Conceptualization (supporting), Methodology (equal), Software (lead), Writing – review & editing (equal).

Lall: Conceptualization (supporting), Data curation (lead), Formal analysis (lead), Investigation (lead), Writing – review & editing (supporting).

## Data availability

Sample code for generating TreeScan input files from a user’s own data source is provided in the Supplement. For transparency and reproducibility of signals related to an extreme heat event, TreeScan input and output files (which contain no confidential patient data) for prospective surveillance on June 29, 2025 are provided in the Supplement.

## Software

The TreeScan software (www.treescan.org) and source code (github.com/scanstatistics/treescan) are freely available.

