## Supplement for "Asyndromic Surveillance of New York City Emergency Department Diagnoses with the Tree-Temporal Scan Statistic"

Table of Contents

| | | |
|---|---|---|
| eMethods 1 | Defining eligible, incident, and unique ICD-10-CM diagnoses per patient per visit. | P. 3 |
| eFigure 1 | Temporal graphs for COVID-19 emergence proof of concept. | P. 4 |
| eFigure 2 | Temporal graphs for heat-related illness. | P. 9 |
| eFigure 3 | Temporal graphs for hand, foot, and mouth disease. | P. 11 |
| eFigure 4 | Temporal graphs for selected injuries. | P. 12 |
| eAppendix 1 | ICD-10-CM codes included and excluded from tree file. | Separate file (.xlsx) |
| eAppendix 2 | ICD-10-CM codes evaluated and not evaluated. | Separate file (.xlsx) |
| Preparing TreeScan input files | • How-to overview (How to prepare TreeScan input files.txt)<br>• Code to create tree file for ICD-10-CM codes in use during period of interest according to the Centers for Medicare & Medicaid Services (1_Create_Tree_File.R)<br>• Sample emergency department dataset using artificial data (Synthetic_Dataset.txt)<br>• Code to create sample TreeScan count file from sample artificial data (2_Create_Count_File.R)<br>• Code to automate daily TreeScan analysis (3_Run_TreeScan.R) | Separate files |
| Reproducing a TreeScan analysis | • How-to overview (How to reproduce an analysis.txt)<br>• Modified count file from analysis conducted on June 29, 2025, where all codes not in the top 50 nodes or their children as detected by TreeScan were replaced with "1- | Separate files |

| | |
|---|---|
| | other" or "0-other" to protect patient confidentiality (Count_File_20250629.txt)<br>• Tree file (Tree_File_20250629.csv)<br>• Do not evaluate nodes file (Do_not_evaluate_nodes.csv)<br>• Parameter file (Parameter_File.prm)<br>• TreeScan results, as summarized in Table 3 (Results_20250629.html)<br>• TreeScan temporal graphs, as depicted in eFigure 2 (Results_20250629.temporal.html) |

**eMethods 1:** Defining eligible, incident, and unique ICD-10-CM diagnoses per patient per visit.

*Eligible*: To improve the signal-to-noise ratio to detect acutely emerging health issues of interest and avoid unnecessary statistical testing, we excluded three categories of ICD-10-CM codes (eAppendix 1 in the Supplement). First, we excluded codes for influenza, COVID-19, asthma, and allergies. These high-volume, seasonal conditions are monitored separately through syndromic surveillance. Second, we excluded certain "Z codes" for factors influencing health status and contact with health services. Such codes (e.g., Z67 for blood type) are neither diagnoses per se nor indicators of acute medical needs. Third, we excluded codes for neoplasms and congenital malformations because population-level increases in these diagnoses cannot plausibly emerge acutely over a few days or weeks.

*Incident*: A substantial portion of ED visits are from frequent users.[1] Repeated diagnoses for the same patient for similar conditions contribute noise when searching for signals of acutely emerging health issues. Accordingly, we excluded occurrences of ICD-10-CM codes where the same patient had a prior ED visit within a 1-year period with a diagnosis in the same tree "subchapter" of three-character categories. For example, N17-N19 (acute kidney failure and chronic kidney disease) is one of eleven ICD-10-CM subchapters for chapter N00-N99 (diseases of the genitourinary system).[2] Suppose a patient first visited the ED for N18.9 (unspecified chronic kidney disease) and then again within a 1-year period for N17.9 (unspecified acute kidney failure). We retained only the first, incident diagnosis for that patient in a 1-year period per subchapter, which here was N18.9. The exception was if the patient was admitted at a later visit. In such an instance, we instead retained the first *admission* diagnosis for that patient in a 1-year period per subchapter as more informative for detecting serious, emerging health issues resulting in hospitalization.

*Unique*: Patients with >1 ED visit per calendar day were considered as a single visit, combining all diagnoses and retaining the more serious discharge disposition of admitted, if applicable. If a patient had >1 incident diagnosis per visit in the same tree subchapter, then we retained the more informative diagnosis that was rarer, according to ED visits citywide in the prior year. If codes within the same tree subchapter were equally common, then we randomly selected a code.

**eFigure 1**. Observed (bar) and expected (line) counts during December 21, 2019–March 14, 2020 of emergency department diagnoses related to COVID-19 emergence in New York City, as signaled (grey shading) in TreeScan by mimicking prospective analysis on March 15, 2020.

R05*: Cough

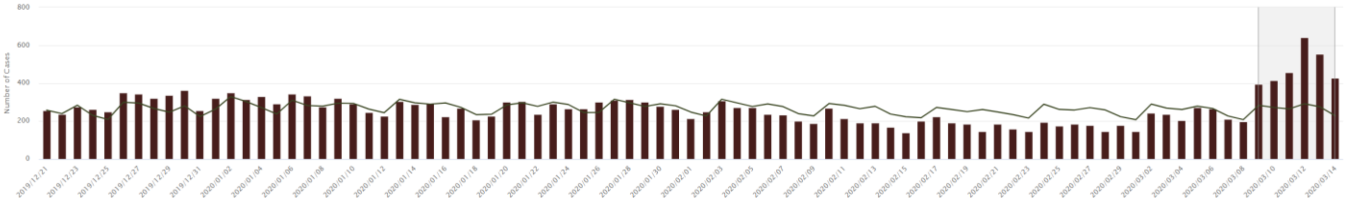

Admissions for R05*: Cough

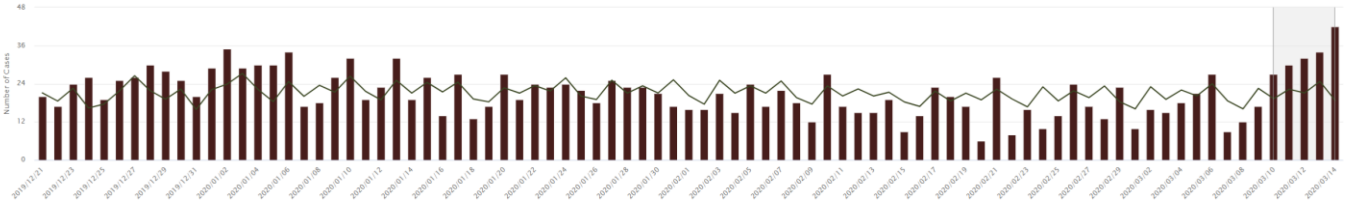

J06.9: Acute upper respiratory infection, unspecified

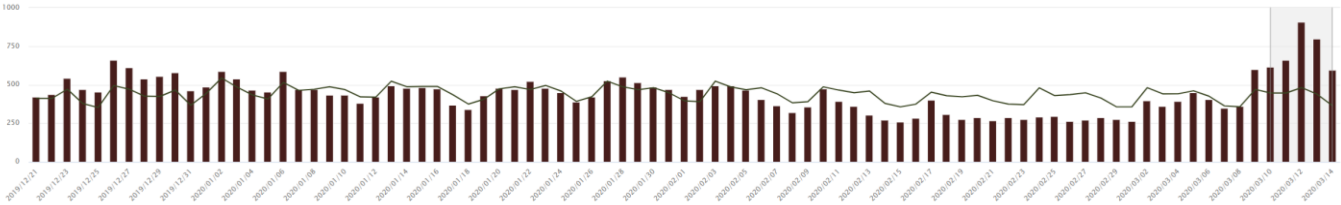

B34*: Viral infection of unspecified site

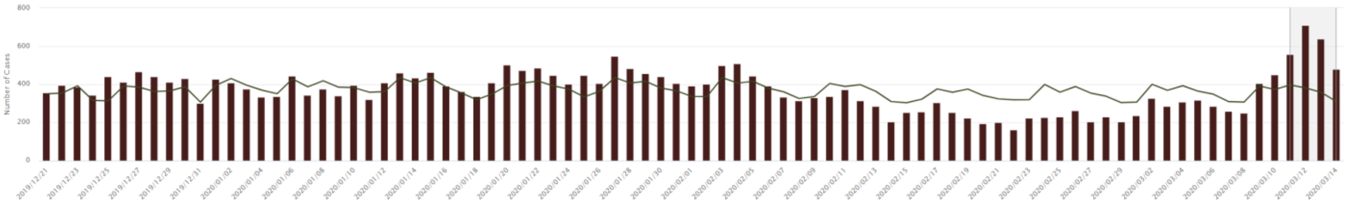

Admissions for B34*: Viral infection of unspecified site

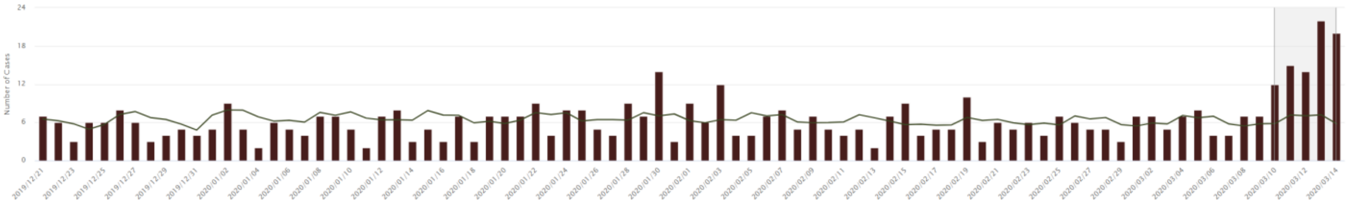

Other coronavirus (supplemental node)

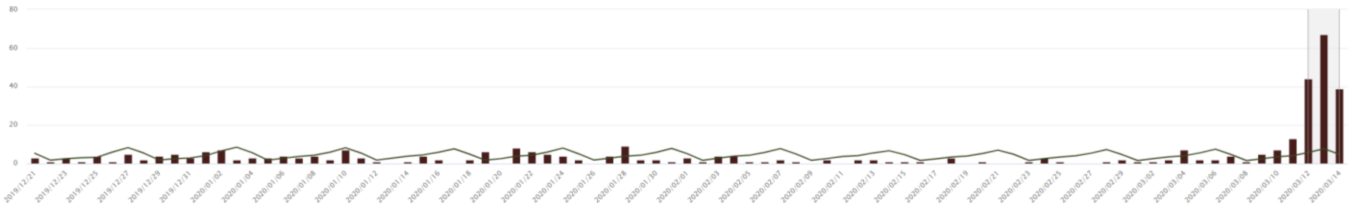

Admissions for other coronavirus (supplemental node)

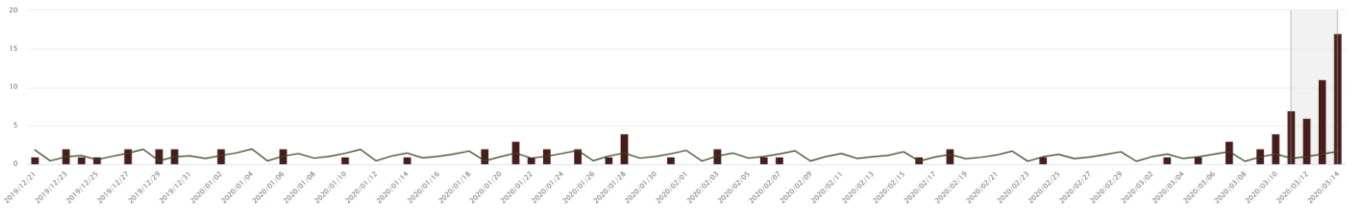

B97.2*: Coronavirus as the cause of diseases classified elsewhere

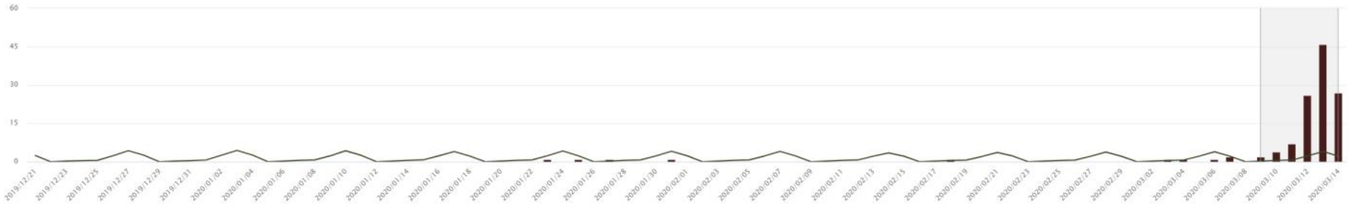

Admissions for B97.2*: Coronavirus as the cause of diseases classified elsewhere

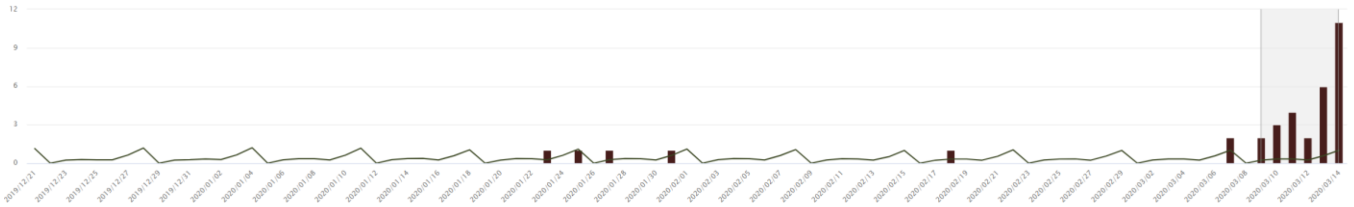

J02.9: Acute pharyngitis, unspecified

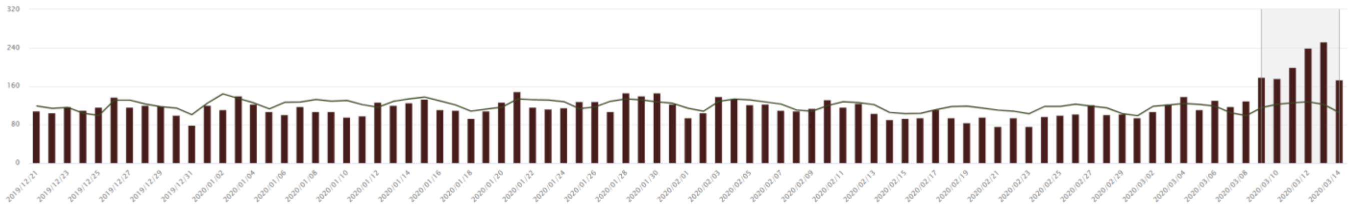

Z20.828: Contact with and (suspected) exposure to other viral communicable diseases

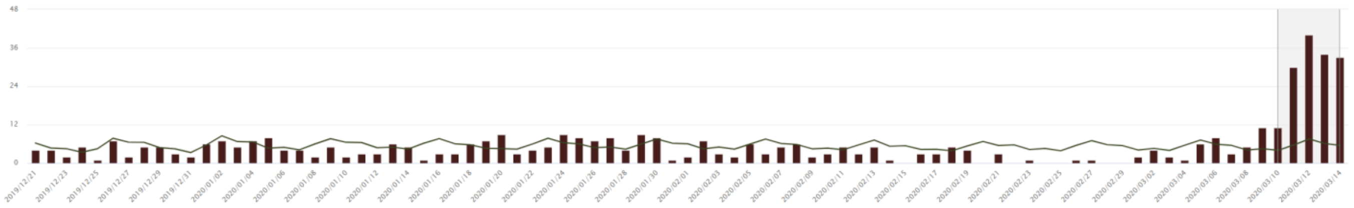

R50*: Fever of other and unknown origin

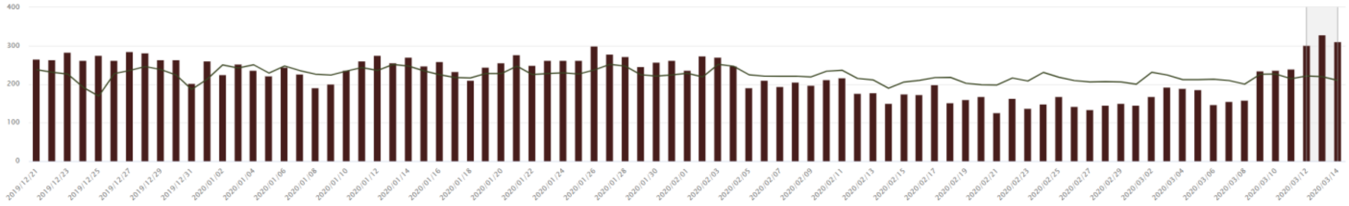

Admissions for R50*: Fever of other and unknown origin

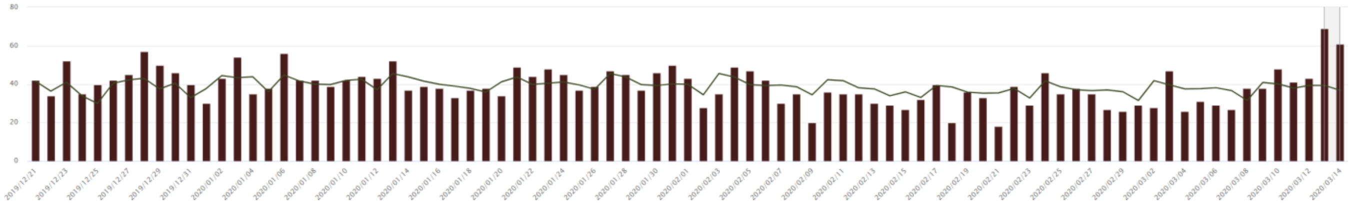

Z71*: Persons encountering health services for other counseling and medical advice, not elsewhere classified

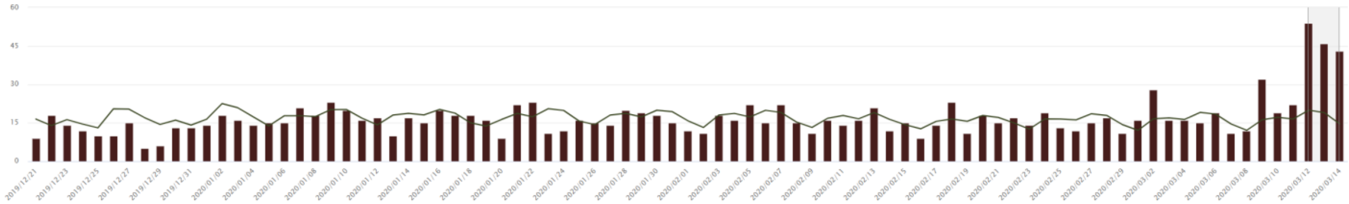

J12*: Viral pneumonia, not elsewhere classified

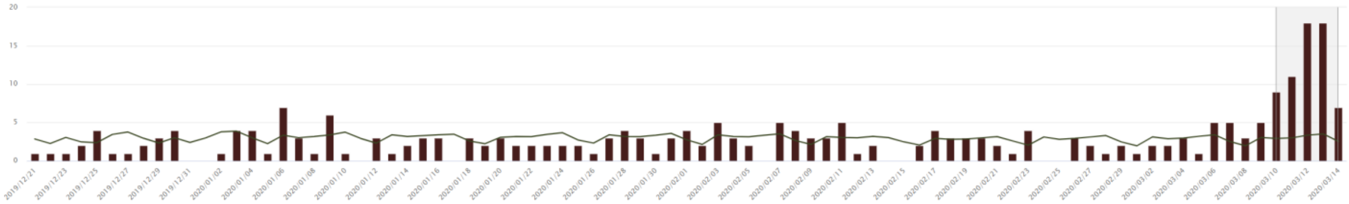

Admissions for J12*: Viral pneumonia, not elsewhere classified

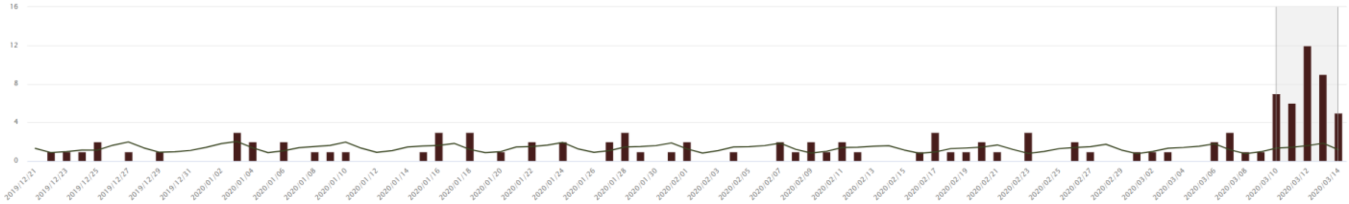

J22: Unspecified acute lower respiratory infection

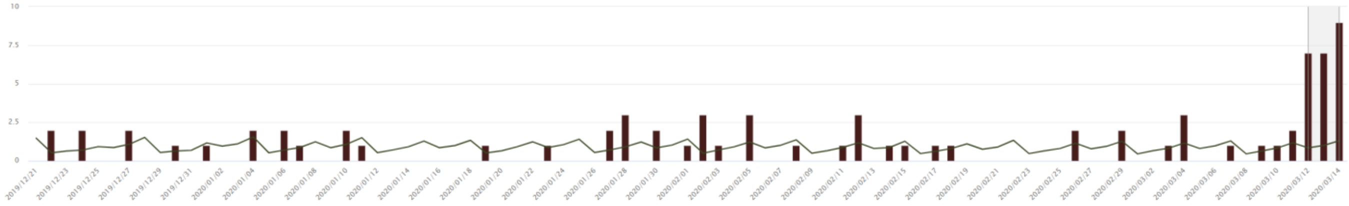

F17.210: Nicotine dependence, cigarettes, uncomplicated

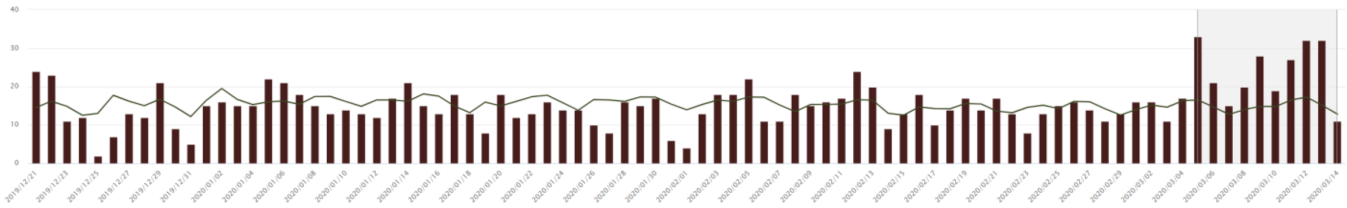

J18*: Pneumonia, unspecified organism

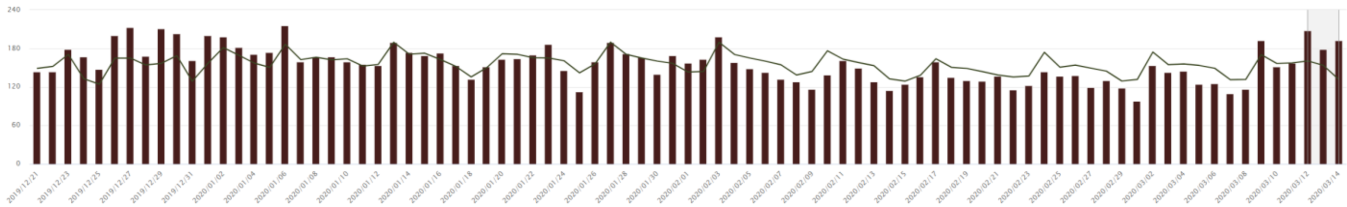

Admissions for J18*: Pneumonia, unspecified organism

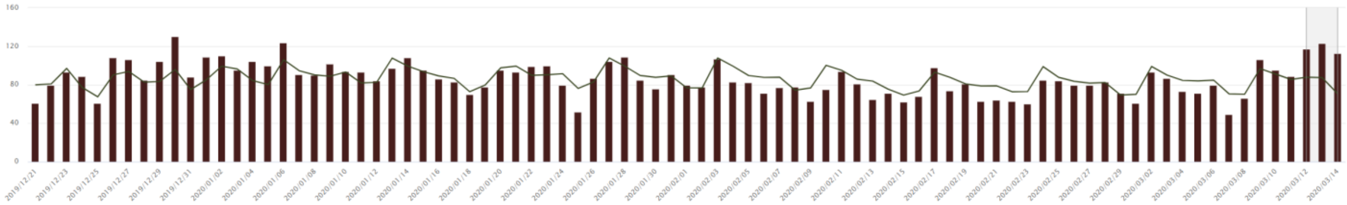

Z11.59: Encounter for screening for other viral diseases

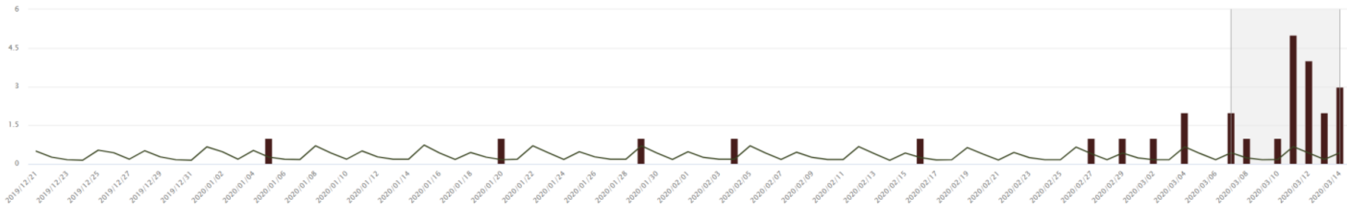

E11.9: Type 2 diabetes mellitus without complications

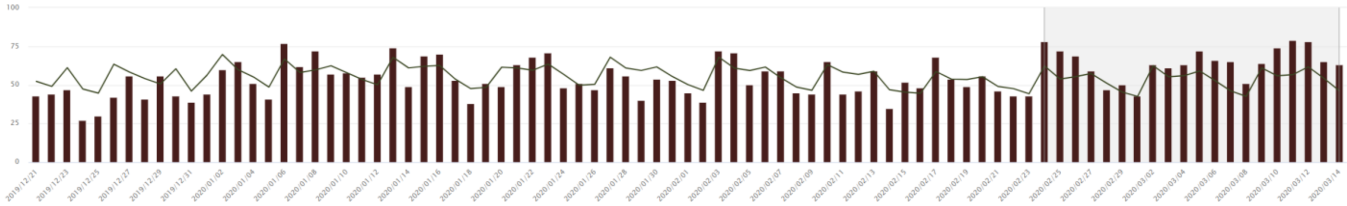

R09.81: Nasal congestion

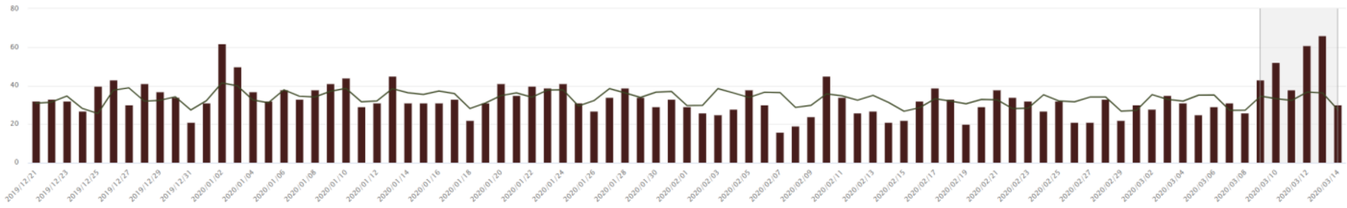

R68.83 : Chills (without fever)

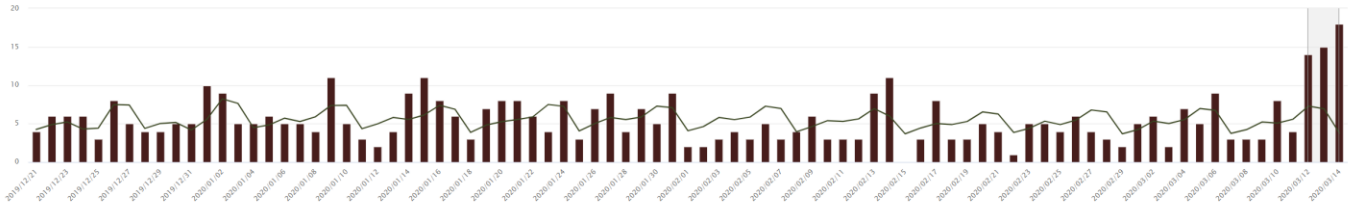

J39.9: Disease of upper respiratory tract, unspecified

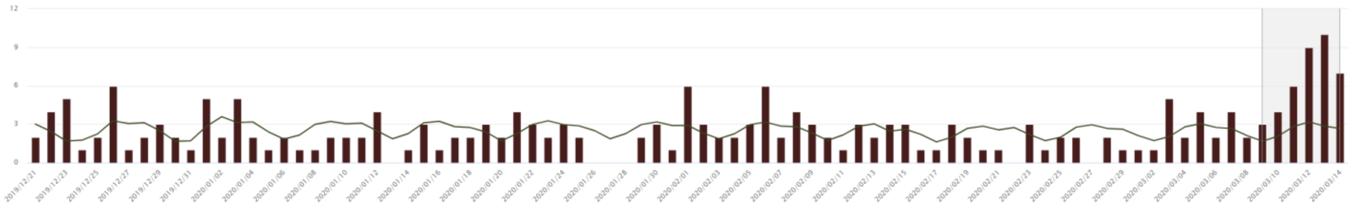

**eFigure 2**. Observed (bar) and expected (line) counts during March 31–June 28, 2025 of emergency department diagnoses related to the June 22–25 extreme heat event in New York City, as signaled (grey shading) in TreeScan on June 29, 2025.

T67*: Effects of heat and light

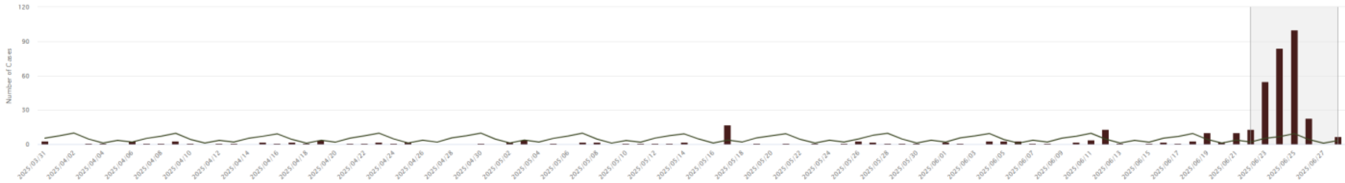

Admissions for T67*: Effects of heat and light

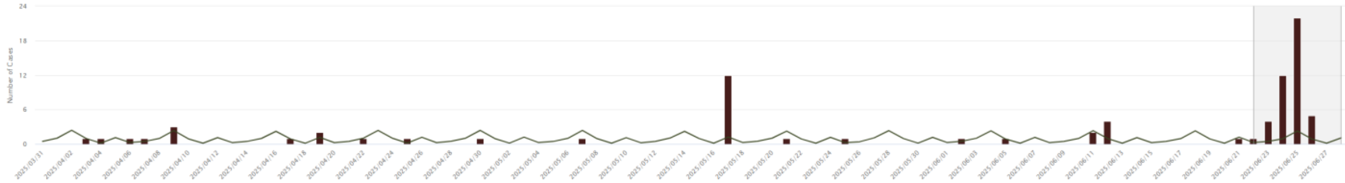

N17*: Acute kidney failure

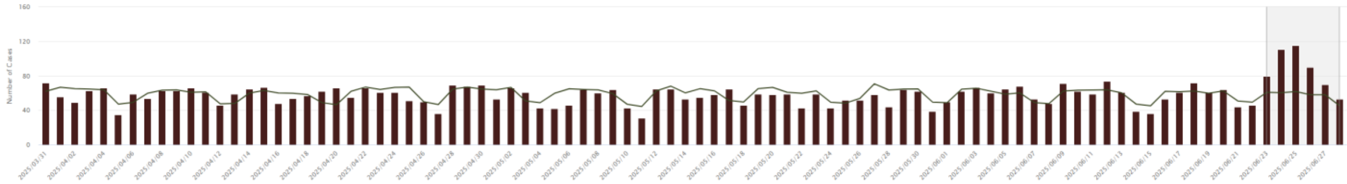

Admissions for N17*: Acute kidney failure

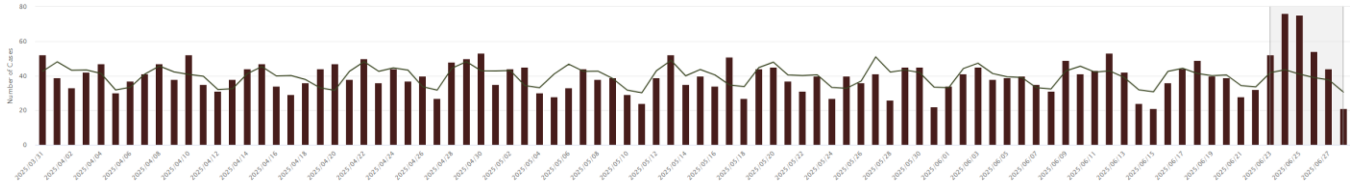

E86*: Volume depletion

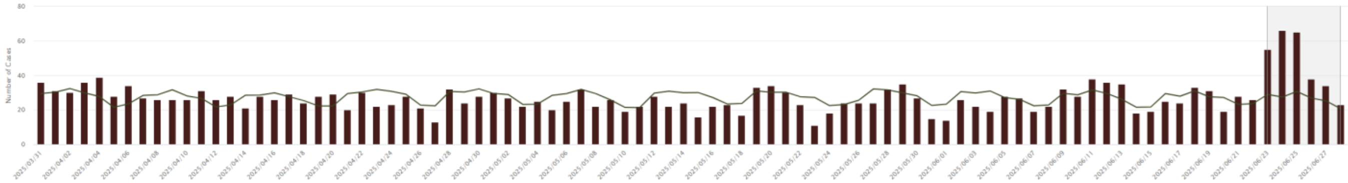

L74.0: Miliaria rubra

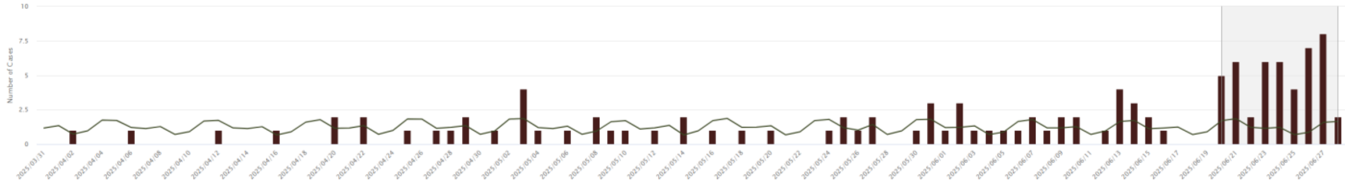

R55*: Syncope and collapse

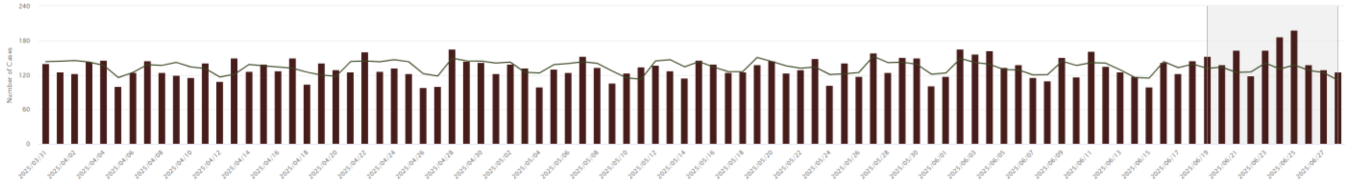

R41.8*: Other symptoms and signs involving cognitive functions and awareness

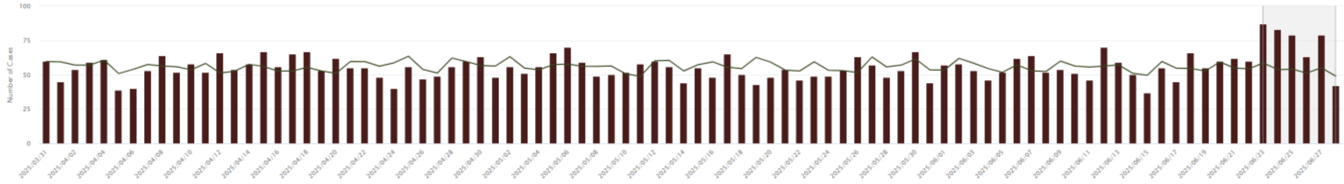

R53*: Malaise and fatigue

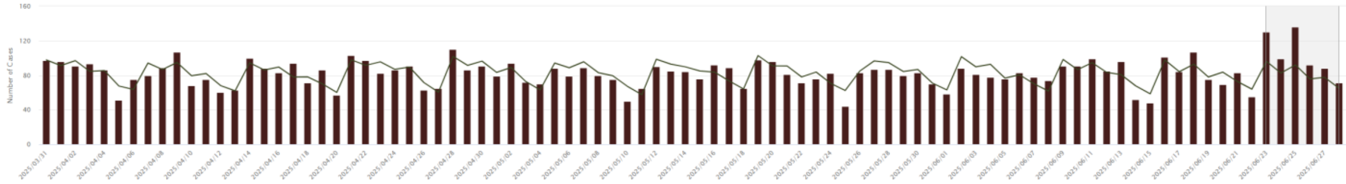

E87.1: Hypo-osmolality and hyponatremia

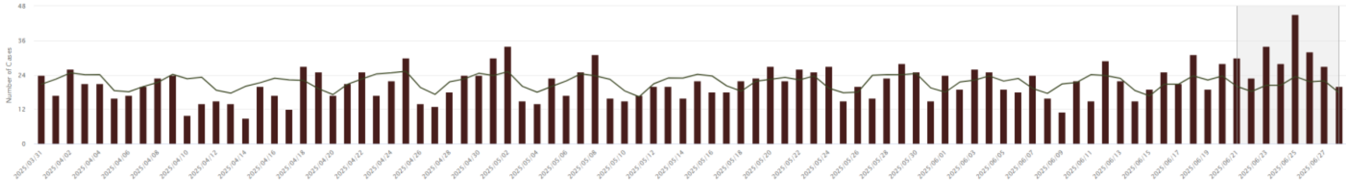

L55*: Sunburn

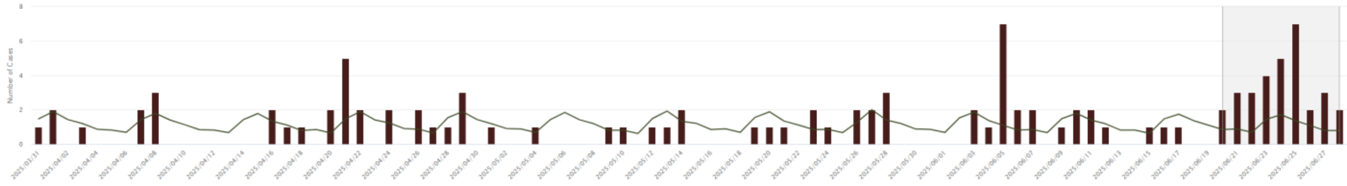

M62.82: Rhabdomyolysis

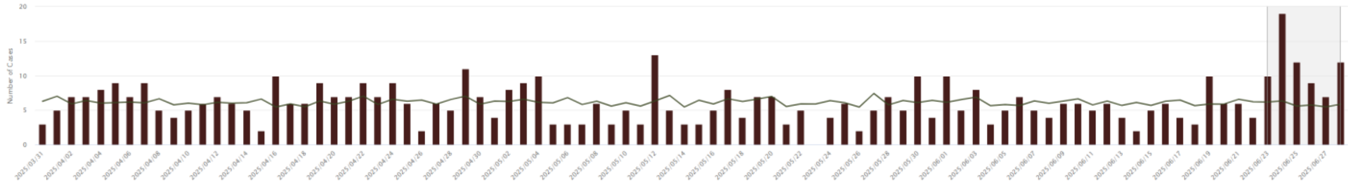

Half marathon-related: T67 (effects of heat and light) during February 17–May 17, 2025, as of May 18.

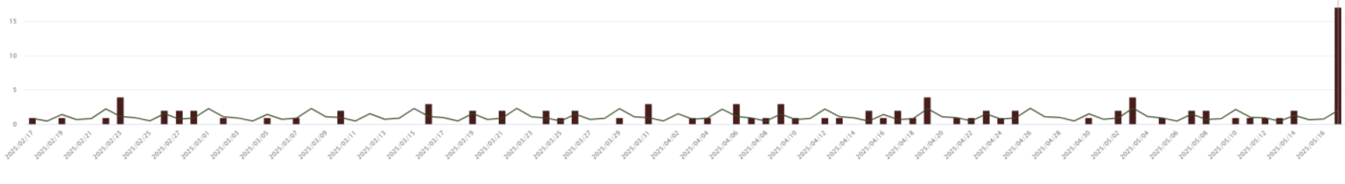

**eFigure 3**. Observed (bar) and expected (line) counts of emergency department diagnoses for B08.4 (enteroviral vesicular stomatitis with exanthem) in New York City, as signaled (grey shading) in TreeScan.

B08.4 diagnoses during January 5–April 4, 2025, as of first signal on April 5, in week 14.

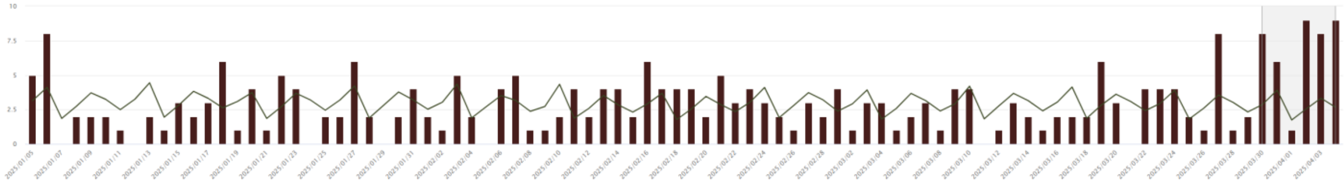

B08.4 diagnoses during May 21–August 18, 2025, as of last signal on August 19, in week 34.

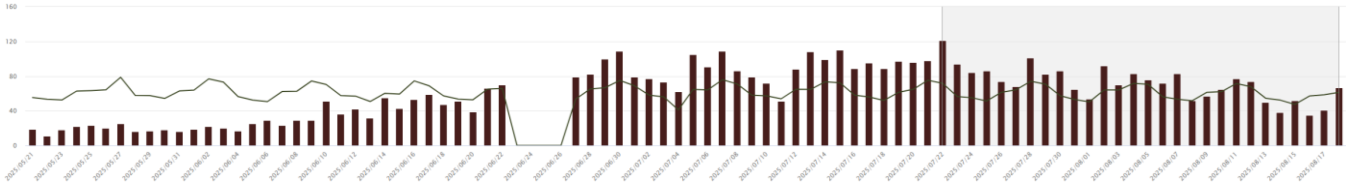

Note above there were zero observed and expected B08.4 diagnoses during June 23–26. This is because we removed all ED visits citywide during the extreme heat event. By removing these days from the baseline, we preserved the ability to detect health effects from subsequent heat waves.

B08.4 diagnoses by week, 2022–2025, as of August 17, 2025, the last full week TreeScan signaled.

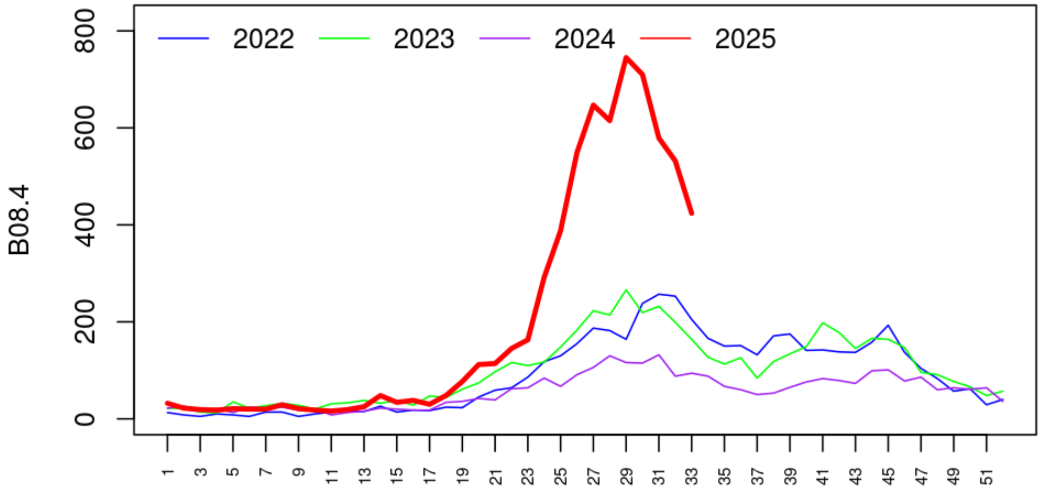

**eFigure 4**. Observed (bar) and expected (line) counts of emergency department diagnoses for selected injuries in New York City, as signaled (grey shading) in TreeScan.

T30.0 (burn of unspecified body region, unspecified degree) during April 6–July 4, 2025, as of July 5.

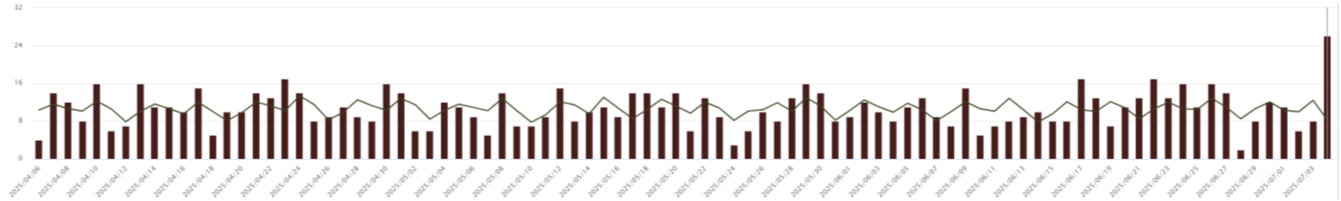

W39 (discharge of firework) during April 10–July 8, 2025, as of July 9.

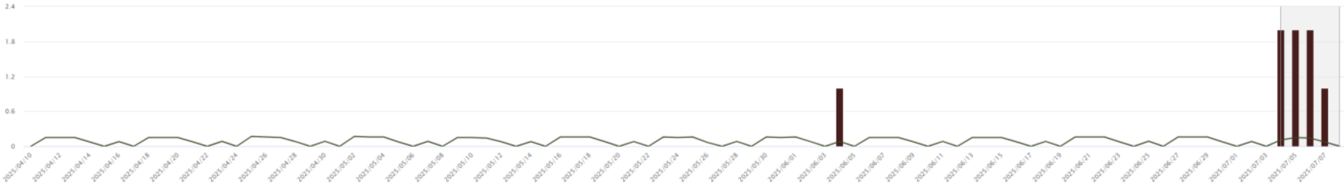

W34.00 (Accidental discharge from unspecified firearms or gun) during May 20–August 17, 2025, as of August 18.

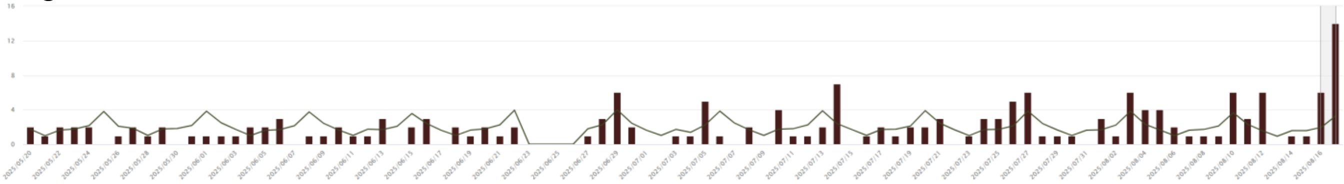
