## Supplementary material for "Asyndromic Surveillance of New York City Emergency Department Diagnoses with the Tree-Temporal Scan Statistic": Preparing TreeScan input files: 1_Create_Tree_File.R

```
## CREATE TREE FILES FOR ASYNDROMIC TREESCAN ANALYSES
## Ramona Lall & Alison Levin-Rector
## October 2025
#
## PART 1:
## READ IN XML FILES TO CREATE TREE FILES IN LONG AND WIDE FORMAT
#
## For PARTS 1 & 2: Download ICD-10 Files from https://www.cms.gov/medicare/coding-billing/icd-10-codes:
# 1) icd10cm_tabular_[year].xml from Code Tables, Tabular and Index ZIP file
# includes all codes except at Level 8
#
# 2) icd10cm_codes_[year].txt from Code Descriptions in Tabular Order ZIP file
# includes all codes from Level 5 to Level 8

library(xml2)
library(dplyr)
library(readr)
library(stringr)

# set your working directory to point to "Preparing TreeScan Input Files" folder
setwd("/run/user/1701674816/gvfs/smb-share:server=nasprgshare220,share=share/DIS/BCD/COMDISshared/Analyst_of_the_week/Asyndromic/Manuscript/Draft/Supplement/Preparing TreeScan Input Files")

year <- 2025

# Read in the XML file
icd10_xml=xml2::read_xml(paste0("files for creating the tree/icd10cm_tabular_",year,".xml"))

# Parse the XML into a list
huge_xml_dict <- xml_find_all(icd10_xml, "//ICD10CM.tabular/chapter")


## Create long tree from xml
# Initialize list to collect data
tree_data <- list()
# Add root manually
tree_data[[length(tree_data) + 1]] <- c("Root", NA, 1, "ICD-10 Root")

# Iterate through each chapter
for (item in huge_xml_dict) {
  desc <- xml_text(xml_find_first(item, "desc"))
  match <- regexpr("(.+)\\((.+)\\)", desc)
  
  if (attr(match, "match.length") > 0) {
    parent_name <- sub("^(.+)\\((.+)\\)$", "\\2", desc)
    full_desc <- sub("^(.+)\\((.+)\\)$", "\\1", desc)
    tree_data[[length(tree_data) + 1]] <- c(parent_name, "Root", 1, full_desc)
    
    sections <- xml_find_all(item, "section")
    
    for (entry in sections) {
      entry_desc <- xml_text(xml_find_first(entry, "desc"))
      entry_id <- xml_attr(entry, "id")
      match <- regexpr("(.+)\\(.+\\)", entry_desc)
      sec_desc <- sub("^(.+)\\((.+)\\)$", "\\1", entry_desc)
      
      if (attr(match, "match.length") > 0) {
        tree_data[[length(tree_data) + 1]] <- c(entry_id, parent_name, 1, sec_desc)
        
        diagnostics <- xml_find_all(entry, "diag")
        if (length(diagnostics) == 0) next
        
        for (diag in diagnostics) {
          diag_name <- xml_text(xml_find_first(diag, "name"))
          diag_desc <- xml_text(xml_find_first(diag, "desc"))
          tree_data[[length(tree_data) + 1]] <- c(diag_name, entry_id, 1, diag_desc)
          
          sub_diagnostics <- xml_find_all(diag, "diag")
          if (length(sub_diagnostics) == 0) next
          
          for (sub_diag in sub_diagnostics) {
            sub_diag_name <- xml_text(xml_find_first(sub_diag, "name"))
            sub_diag_desc <- xml_text(xml_find_first(sub_diag, "desc"))
            tree_data[[length(tree_data) + 1]] <- c(sub_diag_name, diag_name, 1, sub_diag_desc)
            
            sub_diagnostics3 <- xml_find_all(sub_diag, "diag")
            if (length(sub_diagnostics3) == 0) next
            
            for (sub_diag3 in sub_diagnostics3) {
              sub_diag3_name <- xml_text(xml_find_first(sub_diag3, "name"))
              sub_diag3_desc <- xml_text(xml_find_first(sub_diag3, "desc"))
              tree_data[[length(tree_data) + 1]] <- c(sub_diag3_name, sub_diag_name, 1, sub_diag3_desc)
              
              sub_diagnostics4 <- xml_find_all(sub_diag3, "diag")
              if (length(sub_diagnostics4) == 0) next
              
              for (sub_diag4 in sub_diagnostics4) {
                sub_diag4_name <- xml_text(xml_find_first(sub_diag4, "name"))
                sub_diag4_desc <- xml_text(xml_find_first(sub_diag4, "desc"))
                tree_data[[length(tree_data) + 1]] <- c(sub_diag4_name, sub_diag3_name, 1, sub_diag4_desc)
                
                sub_diagnostics5 <- xml_find_all(sub_diag4, "diag")
                if (length(sub_diagnostics5) == 0) next
                
                for (sub_diag5 in sub_diagnostics5) {
                  sub_diag5_name <- xml_text(xml_find_first(sub_diag5, "name"))
                  sub_diag5_desc <- xml_text(xml_find_first(sub_diag5, "desc"))
                  tree_data[[length(tree_data) + 1]] <- c(sub_diag5_name, sub_diag4_name, 1, sub_diag5_desc)
                }
              }
            }
          }
        }
      }
    }
  }
}

# Convert list of rows to data frame
tree_df <- as.data.frame(do.call(rbind, tree_data), stringsAsFactors = FALSE)
colnames(tree_df) <- c("name", "parent", "level", "description")


## Create wide tree from xml
# Initialize list to collect rows
tree_data_wide <- list()
# Define column names
col_names <- c("Name", "Desc", paste0("Level", 1:8))

# Iterate through each chapter
for (item in huge_xml_dict) {
  desc <- xml_text(xml_find_first(item, "desc"))
  match <- regexpr("(.+)\\((.+)\\)", desc)
  
  if (attr(match, "match.length") > 0) {
    parent_name <- sub("^(.+)\\((.+)\\)$", "\\2", desc)
    
    # Iterate through each section in the chapter
    sections <- xml_find_all(item, "section")
    
    for (entry in sections) {
      entry_desc <- xml_text(xml_find_first(entry, "desc"))
      entry_id <- xml_attr(entry, "id")
      match <- regexpr("(.+)\\(.+\\)", entry_desc)
      sec_desc <- sub("^(.+)\\((.+)\\)$", "\\1", entry_desc)
      
      if (attr(match, "match.length") > 0) {
        
        diagnostics <- xml_find_all(entry, "diag")
        if (length(diagnostics) == 0) next
        
        for (diag in diagnostics) {
          diag_name <- xml_text(xml_find_first(diag, "name"))
          diag_desc <- xml_text(xml_find_first(diag, "desc"))
          tree_data_wide[[length(tree_data_wide) + 1]] <- c(
            diag_name, diag_desc, "Root", parent_name, entry_id, diag_name, "", "", "", ""
          )
          
          sub_diagnostics <- xml_find_all(diag, "diag")
          if (length(sub_diagnostics) == 0) next
          
          for (sub_diag in sub_diagnostics) {
            sub_diag_name <- xml_text(xml_find_first(sub_diag, "name"))
            sub_diag_desc <- xml_text(xml_find_first(sub_diag, "desc"))
            tree_data_wide[[length(tree_data_wide) + 1]] <- c(
              sub_diag_name, sub_diag_desc, "Root", parent_name, entry_id, diag_name, sub_diag_name, "", "", ""
            )
            
            sub_diagnostics3 <- xml_find_all(sub_diag, "diag")
            if (length(sub_diagnostics3) == 0) next
            
            for (sub_diag3 in sub_diagnostics3) {
              sub_diag3_name <- xml_text(xml_find_first(sub_diag3, "name"))
              sub_diag3_desc <- xml_text(xml_find_first(sub_diag3, "desc"))
              tree_data_wide[[length(tree_data_wide) + 1]] <- c(
                sub_diag3_name, sub_diag3_desc, "Root", parent_name, entry_id, diag_name, sub_diag_name, sub_diag3_name, "", ""
              )
              
              sub_diagnostics4 <- xml_find_all(sub_diag3, "diag")
              if (length(sub_diagnostics4) == 0) next
              
              for (sub_diag4 in sub_diagnostics4) {
                sub_diag4_name <- xml_text(xml_find_first(sub_diag4, "name"))
                sub_diag4_desc <- xml_text(xml_find_first(sub_diag4, "desc"))
                tree_data_wide[[length(tree_data_wide) + 1]] <- c(
                  sub_diag4_name, sub_diag4_desc, "Root", parent_name, entry_id, diag_name, sub_diag_name, sub_diag3_name, sub_diag4_name, ""
                )
                
                sub_diagnostics5 <- xml_find_all(sub_diag4, "diag")
                if (length(sub_diagnostics5) == 0) next
                
                for (sub_diag5 in sub_diagnostics5) {
                  sub_diag5_name <- xml_text(xml_find_first(sub_diag5, "name"))
                  sub_diag5_desc <- xml_text(xml_find_first(sub_diag5, "desc"))
                  tree_data_wide[[length(tree_data_wide) + 1]] <- c(
                    sub_diag5_name, sub_diag5_desc, "Root", parent_name, entry_id, diag_name, sub_diag_name, sub_diag3_name, sub_diag4_name, sub_diag5_name
                  )
                }
              }
            }
          }
        }
      }
    }
  }
}

# Convert list to data frame
tree_df_wide <- as.data.frame(do.call(rbind, tree_data_wide), stringsAsFactors = FALSE)
colnames(tree_df_wide) <- col_names


## PART 2:
### ADD CODES FROM TXT FILE THAT ARE MISSING FROM XML 

# Read in txt file 
icd10cm.codes <- read.delim(paste0("files for creating the tree/icd10cm_codes_",year,".txt"), header=FALSE)
icd10cm.codes$Name1=substr(icd10cm.codes$V1,1,7)
icd10cm.codes$Name1=gsub(" ","",icd10cm.codes$Name1)

# Read in tree file the wide format with Level1-Level8 for each code
tree_df_wide$Name1=gsub("\\.","",tree_df_wide$Name)

# Merge and identify codes not already in tree
icd10_treefilew_f=merge(tree_df_wide,icd10cm.codes,by="Name1",all=TRUE)
icd10_treefilew_f$added=ifelse(is.na(icd10_treefilew_f$Level2)==T,1,0)

# All codes not in current tree are in Level 8
# Assign all Levels for codes identified as not in Tree
for(i in 1:nrow(icd10_treefilew_f))
{
  if(is.na(icd10_treefilew_f$Level2[i])==T)
  {
    code=paste0(substr(icd10_treefilew_f$Name1[i],1,3),".",substr(icd10_treefilew_f$Name1[i],4,nchar(icd10_treefilew_f$Name1[i])))
    code_len=nchar(code)
    icd10_treefilew_f[i,c("Level1","Level2","Level3","Level4","Level5","Level6","Level7","Level8")][code_len]=code
    for(n in (code_len-1):5)
    {
      if(substr(code,1,n) %in% icd10_treefilew_f[which(icd10_treefilew_f$added==0),c("Level1","Level2","Level3","Level4","Level5","Level6","Level7","Level8")][,n])
      {
        icd10_treefilew_f[i,c("Level1","Level2","Level3","Level4","Level5","Level6","Level7","Level8")][n]=substr(code,1,n)}
    }
    for(n in 4)
    {
      if(substr(code,1,3) %in% icd10_treefilew_f[which(icd10_treefilew_f$added==0),c("Level1","Level2","Level3","Level4","Level5","Level6","Level7","Level8")][,n])
      {
        icd10_treefilew_f[i,c("Level1","Level2","Level3","Level4","Level5","Level6","Level7","Level8")][n]=substr(code,1,3)
      }
    }
    # Add levels 1,2,3
    icd10_treefilew_f[i,c("Level1","Level2","Level3")]=unique(icd10_treefilew_f[which(icd10_treefilew_f$added==0 & icd10_treefilew_f$Level4==icd10_treefilew_f$Level4[i]),c("Level1","Level2","Level3")])  
  }
}

# Some Level8 have missing Level7; for those cases assign last available level
icd10_treefilew_f$Level7[is.na(icd10_treefilew_f$Level8)==F & is.na(icd10_treefilew_f$Level7)==T]=icd10_treefilew_f$Level6[is.na(icd10_treefilew_f$Level8)==F & is.na(icd10_treefilew_f$Level7)==T]
icd10_treefilew_f$Level7[is.na(icd10_treefilew_f$Level8)==F & is.na(icd10_treefilew_f$Level7)==T]=icd10_treefilew_f$Level5[is.na(icd10_treefilew_f$Level8)==F & is.na(icd10_treefilew_f$Level7)==T]
icd10_treefilew_f$Level7[is.na(icd10_treefilew_f$Level8)==F & is.na(icd10_treefilew_f$Level7)==T]=icd10_treefilew_f$Level4[is.na(icd10_treefilew_f$Level8)==F & is.na(icd10_treefilew_f$Level7)==T]

#Fill in Name and Desc of added codes
icd10_treefilew_f$Name[which(nchar(icd10_treefilew_f$Name1)==3 & icd10_treefilew_f$added==1)]=icd10_treefilew_f$Name1[which(nchar(icd10_treefilew_f$Name1)==3 & icd10_treefilew_f$added==1)]
icd10_treefilew_f$Name[which(nchar(icd10_treefilew_f$Name1)>=5 & icd10_treefilew_f$added==1)]=paste0(substr(icd10_treefilew_f$Name1[which(nchar(icd10_treefilew_f$Name1)>=5 & icd10_treefilew_f$added==1)],1,3),".",substr(icd10_treefilew_f$Name1[which(nchar(icd10_treefilew_f$Name1)>=5 & icd10_treefilew_f$added==1)],4,nchar(icd10_treefilew_f$Name1[which(nchar(icd10_treefilew_f$Name1)>=5 & icd10_treefilew_f$added==1)])))

icd10_treefilew_f$Desc[which(icd10_treefilew_f$added==1)]=gsub("^[A-Z][0-9][0-9AB]\\.?[0-9A-TV-Z]{0,4} ","",icd10_treefilew_f$V1[which(icd10_treefilew_f$added==1)])

# Fix Level3 where value is same as Level4
icd10_treefilew_f <- icd10_treefilew_f %>%
  mutate(
    Level3 = if_else(Level3 == Level4, 
                     paste0(Level4, "-", Level4), 
                     Level3)
  )

# Keep Level 7 and Level 8 for codes added to add to Final Tree (Long Format)
codes_to_add=unique(icd10_treefilew_f[which(icd10_treefilew_f$added==1),c("Level8","Level7","Desc")])
codes_to_add$X3=1
codes_to_add=codes_to_add[,c("Level8","Level7","X3","Desc")]


# Add missing codes
colnames(tree_df)[1:4] <- c("X1", "X2", "X3", "X4")
names(codes_to_add)=names(tree_df)
icd10_treefile_added=rbind(tree_df,codes_to_add)

# Fix Level3
# subset codes where parent and child match
temp=icd10_treefile_added$X1[which(icd10_treefile_added$X1==icd10_treefile_added$X2)]

for(i in 1:length(temp))
{
  icd10_treefile_added$X1[which(icd10_treefile_added$X1==temp[i])][1]=paste0(temp[i],"-",temp[i]) 
  icd10_treefile_added$X2[which(icd10_treefile_added$X1==temp[i])]=paste0(temp[i],"-",temp[i]) 
}

# Export wide Tree File, which is used in creating Count File
write.table(icd10_treefilew_f, file = paste0("files for creating the tree/Tree_File_",year,"_wide_format.txt"), quote=FALSE, row.names=FALSE, sep="\t", eol="\r\n")

# Export long Tree File, which is used to update tree files with annual ICD-10 code updates
write.table(icd10_treefile_added, file = paste0("files for creating the tree/Tree_File_",year,"_long_format.txt"), quote=FALSE, row.names=FALSE, sep="\t", eol="\r\n")
## Run this to update tree files with annual ICD-10 code updates:
# lastyear<-year-1
#
### Update wide tree file
## Start with last year's file
#icd10_treefilew_f_lastyear <- read_delim(paste0("files for creating the tree/Tree_File_",lastyear,"_wide_format.txt"), show_col_types=FALSE)
## Merge with this year's file and keep all from both years
#icd10_treefilew_f_bothyears = merge(icd10_treefilew_f,icd10_treefilew_f_lastyear,by="Name",all=TRUE)
#
## .x variables are from this year's file and .y variables are from last year's file
## replace all MISSING .x with all .y 
## prioritizing all new codes and descriptions but not dropping anything from last year's tree
#icd10_treefilew_update <- icd10_treefilew_f_bothyears %>%
#  mutate(
#    Name1.x   = if_else(is.na(Name1.x), Name1.y, Name1.x),
#    Level1.x  = if_else(is.na(Level1.x), Level1.y, Level1.x),
#    Level2.x  = if_else(is.na(Level2.x), Level2.y, Level2.x),
#    Level3.x  = if_else(is.na(Level3.x), Level3.y, Level3.x),
#    Level4.x  = if_else(is.na(Level4.x), Level4.y, Level4.x),
#    Level5.x  = if_else(is.na(Level5.x), Level5.y, Level5.x),
#    Level6.x  = if_else(is.na(Level6.x), Level6.y, Level6.x),
#    Level7.x  = if_else(is.na(Level7.x), Level7.y, Level7.x),
#    Level8.x  = if_else(is.na(Level8.x), Level8.y, Level8.x),
#    V1.x  = if_else(is.na(V1.x), V1.y, V1.x),
#    added.x  = if_else(is.na(added.x), added.y, added.x),
#    Desc.x  = if_else(is.na(Desc.x), Desc.y, Desc.x)
#  ) %>%
#  select(-ends_with(".y")) %>%                         # Drop all .y variables
#  rename_with(~ sub("\\.x$", "", .), ends_with(".x"))  # Remove .x suffixes
#
## save new wide tree file
#write.table(icd10_treefilew_update, file = paste0("files for creating the tree/Tree_File_",year,"_wide_format.txt"), quote=FALSE, row.names=FALSE, sep="\t", eol="\r\n")
#
### Update long tree file
## start with last years file 
#icd10_treefile_added_lastyear <- read_delim(paste0("files for creating the tree/Tree_File_",lastyear,"_long_format.txt"), show_col_types=FALSE)
#
## merge with this years file
## Create source variable that distinguishes whether codes are from last year's file, this year's file, or in both
#icd10_treefile_added$in_current_year <- TRUE
#icd10_treefile_added_lastyear$in_last_year <- TRUE
#icd10_treefile_added_bothyears=merge(icd10_treefile_added,icd10_treefile_added_lastyear,by="X1",all=TRUE)
#icd10_treefile_added_bothyears$in_current_year[is.na(icd10_treefile_added_bothyears$X2.x) & icd10_treefile_added_bothyears$X1!="Root"] <- FALSE
#icd10_treefile_added_bothyears$in_last_year[is.na(icd10_treefile_added_bothyears$X2.y) & icd10_treefile_added_bothyears$X1!="Root"] <- FALSE
#icd10_treefile_added_bothyears$source <- with(icd10_treefile_added_bothyears, 
#    ifelse(in_current_year & in_last_year, "both_years",
#    ifelse(in_current_year, "current_year_only", "last_year_only"))
#)
#icd10_treefile_added_bothyears$in_current_year <- NULL
#icd10_treefile_added_bothyears$in_last_year <- NULL
#
## .x variables are this years and .y variables are from last years file
## replace all MISSING .x with .y
## prioritizing all new codes and descriptions but not dropping anything from tree
## in 2026, only one code (Q00-Q99) was completely dropped - keep in the file but no children assigned to it
#icd10_treefile_update <- icd10_treefile_added_bothyears %>%
#  mutate(
#    X2.x   = if_else(is.na(X2.x), X2.y, X2.x),
#    X3.x  = if_else(is.na(X3.x), X3.y, X3.x),
#    X4.x  = if_else(is.na(X4.x), X4.y, X4.x)
#  ) %>%
#  select(-ends_with(".y")) %>%                         # Drop all .y variables
#  rename_with(~ sub("\\.x$", "", .), ends_with(".x"))  # Remove .x suffixes
#
## save new codes in a separate file for updating the "do not evaluate" file
## Keep rows from current year only
#icd10_treefile_current_year_only <- subset(
#  icd10_treefile_update,
#  source == "current_year_only"
#)
#write.table(icd10_treefile_current_year_only, file = paste0("files for creating the tree/Tree_File_",year,"_current_year_only.txt"), quote=FALSE, row.names=FALSE, sep="\t", eol="\r\n")
#
## save new long tree file
#write.table(icd10_treefile_update, file = paste0("files for creating the tree/Tree_File_",year,"_long_format.txt"), quote=FALSE, row.names=FALSE, sep="\t", eol="\r\n")


## PART 3:
## CREATE FINAL TREE FILE FOR TREESCAN INPUT
# Append supplemental nodes and add in 0-, 1-, 2- prefixes

# import supplemental nodes connecting ICD-10-CM code on different parts of tree that share a common etiology
supplemental_nodes <- read_csv(paste0("files for creating the tree/Common_etiology_nodes.csv"),col_types = cols(X3 = col_character()),show_col_types=FALSE)

# Append supplemental codes
full_w_common <- bind_rows(icd10_treefile_added, supplemental_nodes)

# add in 0- and 1- prefixes for differentiating visits and admits
tree1 <- full_w_common %>%
  transmute(
    child        = str_c("0-",X1),
    parent       = str_c("0-",X2),
    description  = str_c("0-",X4),
    distance_between = X3
  )

tree2 <- full_w_common %>%
  transmute(
    child        = str_c("1-",X1),
    parent       = str_c("1-",X2),
    description  = str_c("1-",X4),
    distance_between = X3
  )

tree3 <- full_w_common %>%
  transmute(
    child        = str_c("0-",X1),
    parent       = str_c("2-",X1),
    description  = str_c("0-",X4),
    distance_between = X3
  )

tree4 <- full_w_common %>%
  transmute(
    child        = str_c("1-",X1),
    parent       = str_c("2-",X1),
    description  = str_c("1-",X4),
    distance_between = X3
  )

tree5 <- full_w_common %>%
  transmute(
    child        = str_c("2-",X1),
    parent       = str_c("2-",X2),
    description  = str_c("2-",X4),
    distance_between = X3
  )

fulltree_final <- bind_rows(tree1, tree2, tree3, tree4, tree5) %>%
  # where parent not in ("0-","1-")
  filter(!parent %in% c("0-", "1-")) %>%
  # keep child parent distance_between description + do same cleaning as SAS
  mutate(
    parent = if_else(parent == "2-", "", parent),
    description = if_else(description %in% c("0-", "1-", "2-"), "", description),
    parent = if_else(child %in% c("0-Root", "1-Root"), "2-Root", parent)
  ) %>%
  select(distance_between, description, child, parent)

# export
write_csv(fulltree_final, paste0("TreeScan input files/Final_Tree_File_",year,".csv"), na = "")
```
