## Supplementary material for "Asyndromic Surveillance of New York City Emergency Department Diagnoses with the Tree-Temporal Scan Statistic": Preparing TreeScan input files: 2_Create_Count_File.R

```
### CREATE COUNT FILE FOR ASYNDROMIC TREESCAN ANALYSIS
## Ramona Lall & Alison Levin-Rector
## October 2025

# Start with synthetic dataset where each row is a unique key (person) and visit date,
# and includes tab delimited diagnosis codes assigned on that visit and 
# an indicator of severity: V=Visit, A=Admitted.
# If using a 90-day study period, require 15 months of data so every day has a year lookback period
# for defining incident diagnoses.

library(lubridate)
library(tidyr)
library(dplyr)

# set your working directory to point to "Preparing TreeScan Input Files" folder
setwd("/run/user/1701674816/gvfs/smb-share:server=nasprgshare220,share=share/DIS/BCD/COMDISshared/Analyst_of_the_week/Asyndromic/Manuscript/Draft/Supplement/Preparing TreeScan Input Files")

year <- 2025

# pull in 15 months of ED visit data ending 8/20/2024
sample_ED_data <- read.delim("synthetic data for count file/Synthetic_Dataset.txt")
sample_ED_data<-mutate(sample_ED_data,date=ymd(date))
class(sample_ED_data$date)
end <- max(sample_ED_data$date)

# Split diagnosis codes into a list (one list per row)
sample_ED_data <- sample_ED_data %>%
  mutate(code = strsplit(diagnosis_codes, split = " "))  # Split diagnosis codes

# Transform so that each row is unique visit-code rather than unique visit 
sample_ED_data_long <- sample_ED_data %>%
  unnest(code) 

# remove spaces
sample_ED_data_long$code=gsub(" |  ","",sample_ED_data_long$code)

# drop diagnosis_codes var
sample_ED_data_long <- sample_ED_data_long %>% 
  select(-diagnosis_codes)

# Define a function to select the most severe outcome
outcome_severity <- function(outcome) {
  # "Admit" is more severe than "Visit"
  if ("A" %in% outcome) {
    return("A")
  }
  return("V")
}

## For repeat visits on same day, keep more severe outcome (visit or admission)
sample_ED_data_long=sample_ED_data_long %>%
  group_by(date, key) %>%
  mutate(
    # Keep the most severe outcome
    severity = outcome_severity(severity)) %>%
  ungroup()

## Remove non-ICD10 codes
# Remove if starts with a digit
sample_ED_data_long=sample_ED_data_long[!grepl("^[0-9]",sample_ED_data_long$code),]
# Remove if no digit in code
sample_ED_data_long=sample_ED_data_long[grepl("\\d", sample_ED_data_long$code),]
# Remove empty values
sample_ED_data_long=sample_ED_data_long[which(sample_ED_data_long$code!=""),]

## Remove repeat rows
sample_ED_data_long <- sample_ED_data_long %>% distinct()

## Remove codes that are ineligible for this analysis
# seasonal codes
eligible_visit_codes=sample_ED_data_long[grepl("U071|J09|J10|J11|\\bJ301\\b|\\bJ302\\b|\\bJ3089\\b|\\bJ309\\b|J45|T7840",sample_ED_data_long$code)==F,]
# certain Z codes
eligible_visit_codes=eligible_visit_codes[grepl("Z0|Z10|Z1152|Z12|Z13|Z14|Z15|Z17|Z18|Z19|Z21|Z28|Z30|Z3|Z4|Z50|Z51|Z52|Z53|Z55|Z56|Z62|Z63|Z64|Z66|Z67|Z68|Z76|Z78|Z8|Z90|Z91|Z92|Z93|Z94|Z95|Z96|Z97|Z98",eligible_visit_codes$code)==F,]
# neoplasms (C00*–D49*) and congenital malformations, deformations and chromosomal abnormalities (Q00*–Q99*)
eligible_visit_codes=eligible_visit_codes[grepl("\\bC|\\bD0|\\bD1|\\bD2|\\bD3|\\bD4|\\bQ",eligible_visit_codes$code)==F,]

## Merge with tree file in wide format to get level2 and level3 parents for each code
icd10_treefile_wide <- read.delim(paste0("files for creating the tree/Tree_File_",year,"_wide_format.txt"))
eligible_visit_codes=merge(eligible_visit_codes, icd10_treefile_wide[,c("Name1","Level2","Level3")],by.x="code",by.y="Name1",all.x=TRUE)

# If code not recognized as belonging to tree file remove
eligible_visit_codes=eligible_visit_codes[is.na(eligible_visit_codes$Level2)==F & is.na(eligible_visit_codes$Level3)==F,]

# Transform again so each row is unique visit rather than unique visit-code
all_visits <- eligible_visit_codes %>%
  group_by(date, key, severity) %>%
  dplyr::summarize(
    Level3 = paste(Level3, collapse = ","),
    code = paste(code, collapse = ","),
    .groups = 'keep'  # keep the grouping for further operations
  )

# Change Level 3 "-" to "_"
all_visits$Level3=gsub("\\-","_",all_visits$Level3)

## Keep only incident diagnoses
# Separate dataset into one for patients with a single visit and one for patients with multiple visits
patient_list=data.frame(table(all_visits$key))
all_visits_single=all_visits[all_visits$key %in% patient_list$Var1[which(patient_list$Freq==1)],c("date","key","severity", "code", "Level3")]
all_visits_multiple=all_visits[all_visits$key %in% patient_list$Var1[which(patient_list$Freq>=2)],c("date","key","severity", "code", "Level3")]

# Define incident diagnosis for patients with multiple visits
# Function to process each unique key
process_patient <- function(patient_data) {

  # Calculate the interval matrix
  interval <- as.matrix(dist(patient_data$date))
  interval[upper.tri(interval, diag = TRUE)] <- NA
  interval <- ifelse(interval <= 365, 1, 0)
  
  # Update Level 3 for visits within 365 days
  for(i in 1:(nrow(interval)-1))
  {interval[,i]=ifelse(interval[,i]==1,gsub(","," ",patient_data$Level3[i]),NA)}

  # Create search strings from prior visits' Level 3 codes
  patient_data$search <- apply(interval, 1, function(x) {
    # If 'x' has no non-NA values, return NONE
    if (all(is.na(x))) {
      return("NONE")
    } else {
      paste0(unique(unlist(strsplit(na.omit(x), " "))), collapse = "|")
    }
  })

  # search = all non admit codes for patient within the past year
  patient_data$search2=NA
  # search2 = all admit codes for patient within the next year
  patient_data$search3=NA
  # searchf = both combined
  patient_data$searchf=NA
  
  # Do this for patients with at least one admit code
  if(nrow(patient_data[which(patient_data$severity=="A"),])>=1)
  {
    admits_only=patient_data[which(patient_data$severity=="A"),]
    
    interval <- as.matrix(dist(admits_only$date))
    interval[lower.tri(interval, diag = TRUE)] <- NA
    interval <- ifelse(interval <= 365, 1, 0)
    
    for(i in 1:nrow(admits_only))
    {
      if(sum(interval[,i],na.rm=T)==0|all(is.na(interval[,i])))
      {patient_data$search2[which(patient_data$severity=="A")[i]]="NONE"}  
      patient_data$search2[which(patient_data$date-patient_data$date[which(patient_data$severity=="A")][i]<0 & patient_data$date-patient_data$date[which(patient_data$severity=="A")][i]>=-365 & patient_data$severity=="V")]=paste0(unique(c(na.omit(unlist(strsplit(patient_data$search2[i],"\\|"))),unlist(strsplit(na.omit(patient_data$Level3[which(patient_data$severity=="A")][i]), ",")))),collapse="|")
    }
    
    if(nrow(admits_only)>=2)
    {
      for(i in 2:nrow(admits_only))
      {
        list_codes=paste0(admits_only$Level3[which((admits_only$date-admits_only$date[i]<0 & admits_only$date-admits_only$date[i]>=-365))],collapse=",")  
        patient_data$search3[which(patient_data$severity=="A")][i]=paste0(setdiff(unlist(strsplit(patient_data$Level3[which(patient_data$severity=="A")][i],",")),unlist(strsplit(list_codes,","))),collapse="|")
      }
    }
  }
  
  for(i in 1:nrow(patient_data))
  {

    patient_data$search[i]=ifelse(is.na(patient_data$search3[i])==F,gsub(patient_data$search3[i],"",patient_data$search[i]),patient_data$search[i])
    
    patient_data$searchf[i]=ifelse(is.na(patient_data$search2[i])==T,patient_data$search[i],
                                   ifelse(is.na(patient_data$search2[i])==F & patient_data$search2[i]!="NONE" & patient_data$search[i]=="NONE",patient_data$search2[i],
                                          ifelse(patient_data$search2[i]=="NONE","NONE",
                                                 paste0(c(patient_data$search[i],na.omit(patient_data$search2[i])),collapse = "|"))))
  }   
  return(patient_data)
}

# Apply the function to each unique key and create 'search' string
# search = all non admit codes for patient within the past year
# search2 = all admit codes for patient within the next year
# searchf = both combined
all_visits_multiple_search <- all_visits_multiple %>%
  group_by(key) %>%
  group_split() %>%
  purrr::map_dfr(process_patient)

# Use 'search string' to find non-incident codes to remove
# remove searchf codes from level3 and subs REMOVE for corresponding code under level3
remove_nonincident <- all_visits_multiple_search %>%
  mutate(Level3 = purrr::map2_chr(Level3, searchf, ~gsub(.y, "REMOVE", .x))) %>%
  mutate(code = purrr::map2_chr(code, Level3, ~{
    codes <- unlist(strsplit(.x, ","))
    lvl3 <- unlist(strsplit(.y, ","))
    codes[lvl3 == "REMOVE"] <- "REMOVE"
    paste0(codes, collapse = ",")
  }))


# merge back with patients with 1 visit
# every row is a unique patient-visit
study_cohort=rbind(all_visits_single[,c("date","key","severity","code")],remove_nonincident[,c("date","key","severity","code")])

# Keep 90-day study period
study_data=study_cohort[which(study_cohort$date>=end-90+1),]

# Make long dataset
# Split diagnosis codes into a list (one list per row)
study_data <- study_data %>%
  mutate(code = strsplit(code, split = ","))  # Split diagnosis codes
# Directly unnest the list into a long format 
study_data_long <- study_data %>%
  unnest(code) 

# remove non-incident codes
study_data_long=study_data_long[which(study_data_long$code!="REMOVE"),]


# Keep unique
# If multiple codes within same visit with same level 3, keep rarest
# string of all codes in original sample dataset, deleting blanks
code=unlist(strsplit(sample_ED_data$diagnosis_codes,split=" "))
code=code[code!=""]
dx_freq_table=data.frame(table(code))

# merge so that we have a column that is the frequency of every code in the original sample dataset
study_data_long=merge(study_data_long, dx_freq_table,by="code",all.x=TRUE)

#if Multiple codes in Level3 keep the rarest
study_data_long=merge(study_data_long,icd10_treefile_wide[,c("Name1","Level3")],by.x="code",by.y="Name1",all.x=TRUE)

# create column n that counts the number of codes with the same level3 during the same visit
# sort in order of increasing frequency, so rarest is first
study_data_long <- study_data_long %>%
  dplyr::arrange(key, date, Level3, Freq) %>%  # Sort the data
  dplyr::group_by(key, date, Level3) %>%       # Group by key, date, Level3
  dplyr::mutate(n = row_number())              # Add row number within each group

# if the >=2 codes in same visit with same level3, keep the first row, which is the rarest
# if there are >=2 visits for the same person with equally rare codes, keep all rarest
keep_rarest <-study_data_long %>%
  dplyr::group_by(key, date, Level3) %>%
  dplyr::filter(n() >= 2 & (Freq == first(Freq))) 
# if codes with same level3 in the same visit are equally rare, keep a random row
tie_breaker <- keep_rarest %>%
  dplyr::group_by(key, date, Level3) %>%
  dplyr::mutate(n = sample(seq_along(Freq), size = n(), replace = FALSE)) %>%  # Randomly shuffle within the group
  dplyr::filter(n == 1)  # Keep only the randomly selected row

# these are all the rest (visits with no repeat level3 codes)
no_repeats <- study_data_long %>%
  dplyr::group_by(key, date, Level3) %>%
  dplyr::filter(!(n() >= 2 & (Freq == first(Freq)))) %>%  
  dplyr::filter(n == 1)  

study_data_long=rbind(no_repeats,tie_breaker)


### CREATE INPUT FILE
# input file is unique visit date, codes and count of that combo
input_file=aggregate(n ~ code+severity+date, data = study_data_long[,c("date","code","severity","n")], FUN = sum)
# add decimal
input_file$code=ifelse(nchar(input_file$code)>=4,paste0(substr(input_file$code,1,3),".",substr(input_file$code,4,nchar(input_file$code))),input_file$code)
# add 0- and 1-
input_file$code1=ifelse(input_file$severity=="V",paste0("0-",input_file$code),paste0("1-",input_file$code))
input_file=input_file[,c("code1","date","n")]
names(input_file)[names(input_file)=="code1"]="code"

write.table(input_file,paste0("TreeScan input files/Analysis_Count_File.txt"), quote=FALSE, row.names=FALSE, sep="\t", eol="\r\n")
```
