## Supplementary material for "Asyndromic Surveillance of New York City Emergency Department Diagnoses with the Tree-Temporal Scan Statistic": Preparing TreeScan input files: 3_Run_TreeScan.R

```
### CREATE PARAMETER FILE FOR ASYNDROMIC ANALYSIS AND RUN TREESCAN IN BATCH MODE
## Ramona Lall & Alison Levin-Rector
## October 2025

library(lubridate)
library(dplyr)

year<-2025

# set your working directory to point to "Preparing TreeScan Input Files" folder
setwd("/run/user/1701674816/gvfs/smb-share:server=nasprgshare220,share=share/DIS/BCD/COMDISshared/Analyst_of_the_week/Asyndromic/Manuscript/Draft/Supplement/Preparing TreeScan Input Files")

# set location of "Preparing TreeScan Input Files" folder
directory <-"//nasprgshare220/Share/DIS/BCD/COMDISshared/Analyst_of_the_week/Asyndromic/Manuscript/Draft/Supplement/Preparing TreeScan Input Files/"

# First set up Param file and save to folder where all files need to be for Run
makeprm_2_3_0<- function (treefile, countfile, date_range, resultsfile, paramfile) {
  prminput <- c('[Analysis]',
                #scan type (TREEONLY=0, TREETIME=1, TIMEONLY=2)
                "scan-type=1",
                #probability model type (POISSON=0, BERNOULLI_TREE=1, UNIFORM=2, Not-Applicable=3)
                "probability-model=0",
                #conditional type (UNCONDITIONAL=0, TOTALCASES=1, NODE=2, NODEANDTIME=3)
                "conditional-type=3",
                #self control design - unconditional Bernoulli only (y/n)
                "self-control-design=n",
                #case probability (integer/integer)
                "event-probability=1/2",
                #variable case probability - unconditional Bernoulli only (y/n)
                "variable-case-probability=n",
                #scan rate type (HIGHRATE=0, LOWRATE=1, HIGHORLOWRATE=2)
                "scan-rate-type=0",
                
                '[Input]',
                #tree structure filename
                paste("tree-filename=",treefile),
                #source type (CSV=0)
                "tree-filename-SourceType=0",
                #source field map (comma separated list of integers)
                "tree-filename-SourceFieldMap=3,4,1,2",
                #csv source delimiter (leave empty for space or tab delimiter)
                "tree-filename-SourceDelimiter=,",
                #csv source group character
                'tree-filename-SourceGrouper="',
                #csv source skip initial lines (i.e. meta data)
                "tree-filename-SourceSkip=0",
                #csv source first row column header
                "tree-filename-SourceFirstRowHeader=y",
                #count data filename
                paste("count-filename=",countfile),
                #source type (CSV=0)
                "count-filename-SourceType=0",
                #source field map (comma separated list of integers)
                "count-filename-SourceFieldMap=1,3,2",
                #csv source delimiter (leave empty for space or tab delimiter)
                'count-filename-SourceDelimiter=" "',
                #csv source group character
                'count-filename-SourceGrouper="',
                #csv source skip initial lines (i.e. meta data)
                "count-filename-SourceSkip=0",
                #csv source first row column header
                "count-filename-SourceFirstRowHeader=y",
                #control data filename
                "control-filename=",
                #date precision type (NONE=0, GENERIC=1, YEAR=2, MONTH=3, DAY=4)
                "date-precision=4",
                #data time ranges: [integer,integer] or [yyyy/mm/dd,yyyy/mm/dd]
                paste0("data-time-range=",date_range),
                
                '[Output]',
                #results filename
                paste("results-filename=",resultsfile),
                #create HTML results (y/n)
                "results-html=y",
                #create CSV results (y/n)
                "results-csv=y",
                #create NCBI Asn results (y/n)
                "results-asn=n",
                #create Newick File (y/n)
                "results-nwk=n",
                
                '[Advanced Input]',
                #cuts filename
                "cut-filename=",
                #default cuts type (SIMPLE=0, PAIRS=1, TRIPLETS=2, ORDINAL=3, COMBINATORIAL=4)
                "cut-type=0",
                #allow data only on tree leaves - (y/n)
                "data-only-on-leaves=n",
                #ignore cases outside study period - (y/n)
                "relaxed-study-data-period-checking=y",
                #allow multi-parent nodes - (y/n)
                "allow-multi-parent-nodes=y",
                #allow multiple root nodes - (y/n)
                "allow-multiple-roots=n",
                #minimum censor time (2 <= x)
                "minimum-censor-time=50",
                #minimum censor time percentage of study period (0 < x <= 100.0)
                "min-censor-percentage=10",
                #apply risk window restriction due to censoring - (y/n)
                "risk-window-restriction-censor=n",
                #risk window alternative censor denominator (integer)
                "risk-window-alt-censor-denominator=2",
                
                '[Temporal Window]',
                #maximum temporal size as percentage of data time range (0 < x <= 50.0)
                "maximum-window-percentage=50",
                #maximum temporal size as fixed time length (integer)
                "maximum-window-fixed=28",
                #maximum temporal size selection (PERCENTAGE_WINDOW=0, FIXED_LENGTH=1)
                "maximum-window-type=1",
                #minimum temporal size as fixed time length (integer)
                "minimum-window-fixed=1",
                #apply risk window restriction - (y/n)
                "apply-risk-window-restriction=n",
                #risk window percentage (0 < x <= 100.0)
                "risk-window-percentage=20",
                #prospective analysis (y/n)
                "prospective-analysis=y",
                #restrict temporal windows (y/n)
                "restricted-window-range=n",
                #start data time range: [integer,integer] or [yyyy/mm/dd,yyyy/mm/dd]
                paste0("window-start-range=",date_range),
                #end data time range: [integer,integer] or [yyyy/mm/dd,yyyy/mm/dd]
                paste0("window-end-range=",date_range),
                
                '[Adjustments]',
                #perform day of week adjustments (y/n)
                "perform-day-of-week-adjustments=y",
                #apply exclusion time ranges (y/n)
                "apply-exclusion-data-ranges=n",
                #exclusion time ranges (semi-colon separated list of ranges: [integer,integer];[integer,integer] or [yyyy/mm/dd,yyyy/mm/dd];[yyyy/mm/dd,yyyy/mm/dd])
                "exclusion-data-ranges=",
                
                '[Inference]',
                #number of simulation replications (0, 9, 999, n999)
                "monte-carlo-replications=99999",
                #restrict tree levels evaluated (y/n)
                "restrict-tree-levels=y",
                #tree levels excluded from evaluation (csv list of unsigned integers, root level is 1)
                "excluded-tree-levels=1,2,3",
                #restrict tree nodes evaluated (y/n)
                "restrict-evaluated-nodes=y",
                #not evaluated tree nodes filename
                "not-evaluated-nodes-file=Do_not_evaluate_nodes.csv",
                #source type (CSV=0)
                "not-evaluated-nodes-file-SourceType=0",
                #source field map (comma separated list of integers)
                "not-evaluated-nodes-file-SourceFieldMap=1",
                #csv source delimiter (leave empty for space or tab delimiter)
                "not-evaluated-nodes-file-SourceDelimiter=,",
                #csv source group character
                'not-evaluated-nodes-file-SourceGrouper="',
                #csv source skip initial lines (i.e. meta data)
                "not-evaluated-nodes-file-SourceSkip=0",
                #csv source first row column header
                "not-evaluated-nodes-file-SourceFirstRowHeader=y",
                #randomization seed (integer)
                "randomization-seed=12345678",
                #generate randomization seed (y/n)
                "random-randomization-seed=n",
                #minimum number of cases in a node (integer)
                "minimum-node-cases=3",
                #p-value reporting type (STANDARD_PVALUE=0, TERMINATION_PVALUE)
                "pvalue-report-type=1",
                #early termination threshold (> 0)
                "early-termination-threshold=5000",
                
                '[Sequential Scan]',
                #perform sequential scan - time-only scan (y/n)
                "sequential-scan=n",
                #sequential scan maximum cases for signal (integer)
                "sequential-maximum-signal=200",
                #sequential scan - minimum cases to signal (integer)
                "sequential-minimum-signal=3",
                #sequential scan filename
                "sequential-filename=",
                #sequential alpha overall
                "sequential-alpha-overall=0.05",
                #sequential alpha spending
                "sequential-alpha-spending=0.01",
                
                '[Power Evaluations]',
                #perform power evaluations (y/n)
                "perform-power-evaluations=n",
                #power evaluation type (0=Analysis And Power Evaluation Together, 1=Only Power Evaluation With Count File, 2=Only Power Evaluation With Defined Total Cases)
                "power-evaluation-type=0",
                #critical values type (0=Monte Carlo, 1=User Specified Values)
                "critical-values-type=0",
                #power evaluation critical value .05 (> 0)
                "critical-value-05=0",
                #power evaluation critical value .01 (> 0)
                "critical-value-01=0",
                #power evaluation critical value .001 (> 0)
                "critical-value-001=0",
                #total cases in power evaluation (integer)
                "power-evaluation-totalcases=600",
                #number of replications in power step (integer)
                "power-evaluation-replications=1000",
                #power evaluation alternative hypothesis filename
                "alternative-hypothesis-filename=",
                #power baseline probability (integer/integer)
                "baseline-probability=1/2",
                #power z value (0 < z <= 0.01)
                "power-z=0.001",
                
                '[Miscellaneous Analysis]',
                #frequency of prospective analyses type (0=Daily, 1=Weekly, 2=Monthy, 3=Quarterly, 4=Yearly)
                "prospective-frequency-type=0",
                #frequency of prospective (integer)
                "prospective-frequency=1",
                
                '[Additional Output]',
                #create LLR results (y/n)
                "results-llr=n",
                #report critical values (y/n)
                "report-critical-values=y",
                #report attributable risk (y/n)
                "report-attributable-risk=n",
                #number of exposed attributable risk is based upon (positive integer)
                "attributable-risk-exposed=0",
                #report parent cuts that match child cuts (y/n)
                "include-identical-parent-cuts=n",
                #output temporal graph HTML file (y/n)
                "output-temporal-graph-html=y",
                #temporal graph cluster reporting type (0=Only most likely cluster, 1=X most likely clusters, 2=Only significant clusters)
                "temporal-graph-type=1",
                #number of most likely clusters to report in temporal graph (positive integer)
                "temporal-graph-most-mlc=100",
                #significant clusters p-value cutoff to report in temporal graph (0.000-1.000)
                "temporal-graph-significance-cutoff=0.05",
                
                '[Power Simulations]',
                #input simulation data (y/n)
                "input-simulations=n",
                #input simulation filename
                "input-simulations-file=",
                #output simulation data (y/n)
                "output-simulations=n",
                #output simulation filename
                "output-simulations-file=",
                
                '[Run Options]',
                #number of parallel processes to execute (0=All Processors, x=At Most X Processors)
                "parallel-processes=0",
                
                '[System]',
                #;parameters version - do not modify
                "parameters-version=2.3.0")
  
  write(prminput,paramfile)
}

input_file <- read.delim("TreeScan input files/Analysis_Count_File.txt")
input_file<-mutate(input_file,date=ymd(date))
class(input_file$date)

treefile = paste0(directory,"TreeScan input files/Final_Tree_File_",year,".csv")
countfile = paste0(directory,"TreeScan input files/Analysis_Count_File.txt")
date_range = paste0("[",format(min(input_file$date), "%Y/%m/%d"),",",format(max(input_file$date), "%Y/%m/%d"),"]")
resultsfile = paste0(directory,"TreeScan output files/Results_",format(max(input_file$date), "%Y%m%d"),".txt")
paramfile = paste0("TreeScan input files/Param_File.prm")
makeprm_2_3_0(treefile, countfile, date_range, resultsfile, paramfile)

#Run in batch mode
system(paste("/opt/treescan.2.3/treescan64" ,' ', paramfile, sep=''))
```
