## Supplementary material for "Asyndromic Surveillance of New York City Emergency Department Diagnoses with the Tree-Temporal Scan Statistic": Preparing TreeScan input files: How to prepare TreeScan input files.txt

How to create input files and run a sample TreeScan analysis using synthetic data:

1. Download the "Preparing TreeScan Input Files" folder to your computer.

2. Download ICD-10 Files from https://www.cms.gov/medicare/coding-billing/icd-10-codes and save "icd10cm_tabular_2025.xml" from the "Code Tables, Tabular and Index" ZIP file 
	and "icd10cm_codes_2025.txt" from the "Code Descriptions in Tabular Order" ZIP file in the "files for creating the tree" folder.

3. Run the R code in the "code" folder. At the beginning of each code file, set the working directory to point to where you saved the "Preparing TreeScan Input Files" folder.
	1_Create_Tree_File.R 
		- Reads in files in the "files for creating tree" folder and outputs two intermediate tree files ("Tree_File_2025_long_format.txt" and "Tree_File_2025_wide_format.txt") 
			in the same folder as well as the final tree file for the TreeScan analysis ("Final_Tree_2025.csv") in the "TreeScan input files" folder.
		- There is also a portion of the code (Lines 287-362) that can be uncommented to merge multiple years of ICD-10 Files
			-To do this for 2026, download "icd10cm_tabular_2026.xml" and "icd10cm_codes_2026.txt" from the website above, and set the year <- 2026 at the top of the code
			-This code uses the "Tree_File_2025_long_format.txt" and "Tree_File_2025_wide_format.txt" files to update the tree file
			-This will also output a list of nodes that were newly added that can be referenced for updating the the "Do_not_evaluate_nodes.csv" file

	2_Create_Count_File.R
		- Reads in "Synthetic_Dataset.txt" in the "synthetic dataset for count file" folder, which contains 15 months of anonymized ED visit data
		- Uses "Tree_File_2025_wide_format.txt" created in 1_Create_Tree_File.R to merge in parents
		- Outputs the count file for the TreeScan analysis ("Analysis_Count_File.txt") in the "TreeScan input files" folder

	3_Run_TreeScan.R
		- Creates the parameter settings file for the TreeScan analysis ("Param_file.prm") in the "TreeScan input files" folder
			-This file can be opened in the TreeScan software and run from there
		- The parameter file points to the files in the "TreeScan input files" folder ("Final_Tree_2025.csv","Analysis_Count_File.txt", and "Do_not_evaluate_nodes.csv")
 		- The last line of this code file prompts R to run TreeScan in batch mode
		- When run, TreeScan will create four output files in the "TreeScan output files" folder:
			1. "Results_20250820.txt"
			2. "Results_20250820.html"
			3. "Results_20250820.temporal.html"
			4. "Results_20250820.csv"

4. You can review the resulting output files to inspect the results of the analysis. These output files do not represent real results.
