## Supplementary material for "Asyndromic Surveillance of New York City Emergency Department Diagnoses with the Tree-Temporal Scan Statistic": Reproducing a TreeScan analysis: How to reproduce an analysis.txt

For reproducibility of signals related to an extreme heat event in New York City, 
TreeScan input and output files (which contain no confidential patient data) 
for prospective surveillance on June 29, 2025 are provided.
These input files can be used to recreate the top 50 nodes detected by the TreeScan analysis.

ICD-10 codes that did not appear in the top 50 most likely clusters in the original June 29, 2025 analysis were substituted with "other" for confidentiality.
The Tree File has been modified to add nodes for "other" visits and "other" admits.
We also provide a results file and temporal graph output file that you can review without needing to run the program.
Recurrence intervals for certain nodes in the results file are stronger than as presented in Table 3 due to the modifications in the count file.

To run analysis,
- Open parameter file in TreeScan, and update file locations in four places:
	1. Input tab > Tree File
	2. Input tab > Count File
	3. Analysis tab > Advanced button > Inference tab > Do not evaluate nodes file
	4. Output tab > Results File

- Run by clicking green arrow icon in toolbar

- Navigate to results file location to review results. Four output files will be created (.txt, .csv, .html, .temporal.html).
