## Supplementary material for "Asyndromic Surveillance of New York City Emergency Department Diagnoses with the Tree-Temporal Scan Statistic": Reproducing a TreeScan analysis: Results_20250629.html

xml version="1.0" encoding="UTF-8"?


Tree Temporal scan
looking for high rate branches
conditioned on node and time

### SUMMARY STATISTICS

|  |  |
| --- | --- |
| Tree |  |
| Number of Nodes: | 279531 |
| Number of Root Nodes: | 1 |
| Number of Nodes with Children: | 138437 |
| Number of Leaf Nodes: | 141094 |
| Number of Levels in Tree: | 9 |
| Nodes per Levels: | 1, 26, 345, 2246, 11745, 32826, 58656, 93586, 80100 |
| Number of Nodes Evaluated: | 50838 |
| Tree Levels Included: | 4, 5, 6, 7, 8, 9 |
| Tree Levels Excluded: | 1, 2, 3 |
| Data Summary |  |
| Total Cases: | 1047210 |
| Data Time Range: | [2025/03/31,2025/06/28] |

#### MOST LIKELY CUTS

| No. | Node ID | Node Name | Tree Level | Node Cases | Time Window | Cases in Window | Expected | Relative Risk | Excess Cases | Test Statistic | P-value | Recurrence Interval | Parent Node | Parent Node Name | Branch Order |
| --- | --- | --- | --- | --- | --- | --- | --- | --- | --- | --- | --- | --- | --- | --- | --- |
| 1 | 2-B08.4 | 2-Enteroviral vesicular stomatitis with exanthem | 5 | 1872 | 2025/06/10 to 2025/06/28 | 1070 | 386.03 | 5.16 | 862.48 | 407.123304 | 0.00001 | 274 years | 2-B08,2-Other Enterovirus | 2-Other viral infections characterized by skin and mucous membrane lesions, not elsewhere classified,2-Other Enterovirus | 32 |
| 2 | 2-B08 | 2-Other viral infections characterized by skin and mucous membrane lesions, not elsewhere classified | 4 | 2302 | 2025/06/10 to 2025/06/28 | 1217 | 475.35 | 4.34 | 936.35 | 402.701624 | 0.00001 | 274 years | 2-B00-B09 | 2-Viral infections characterized by skin and mucous membrane lesions | 30 |
| 3 | 2-Other Enterovirus |  | 4 | 2965 | 2025/06/10 to 2025/06/28 | 1423 | 611.24 | 3.57 | 1024.30 | 391.035689 | 0.00001 | 274 years | 2-dummy2 |  | 105 |
| 4 | 2-T67 | 2-Effects of heat and light | 4 | 425 | 2025/06/22 to 2025/06/28 | 282 | 31.55 | 24.52 | 270.50 | 367.225377 | 0.00001 | 274 years | 2-T66-T78 | 2-Other and unspecified effects of external causes | 96 |
| 5 | 2-T67.5 | 2-Heat exhaustion, unspecified | 5 | 195 | 2025/06/22 to 2025/06/28 | 155 | 14.46 | 48.12 | 151.78 | 227.115164 | 0.00001 | 274 years | 2-T67 | 2-Effects of heat and light | 101 |
| 6 | 2-T67.9 | 2-Effect of heat and light, unspecified | 5 | 54 | 2025/06/21 to 2025/06/28 | 47 | 4.24 | 72.31 | 46.35 | 70.344038 | 0.00001 | 274 years | 2-T67 | 2-Effects of heat and light | 103 |
| 7 | 2-T67.01 | 2-Heatstroke and sunstroke | 6 | 53 | 2025/06/23 to 2025/06/28 | 39 | 3.81 | 40.37 | 38.03 | 55.512761 | 0.00001 | 274 years | 2-T67.0 | 2-Heatstroke and sunstroke | 98 |
| 8 | 2-T67.0 | 2-Heatstroke and sunstroke | 5 | 56 | 2025/06/23 to 2025/06/28 | 39 | 3.95 | 33.24 | 37.83 | 54.219221 | 0.00001 | 274 years | 2-T67 | 2-Effects of heat and light | 97 |
| 9 | 1-T67 | 1-Effects of heat and light | 5 | 81 | 2025/06/22 to 2025/06/28 | 44 | 5.99 | 14.75 | 41.02 | 49.748824 | 0.00001 | 274 years | 1-T66-T78,2-T67 | 1-Other and unspecified effects of external causes,2-Effects of heat and light | 25 |
| 10 | 2-W57 | 2-Bitten or stung by nonvenomous insect and other nonvenomous arthropods | 4 | 1281 | 2025/06/07 to 2025/06/28 | 474 | 305.32 | 1.88 | 221.51 | 39.822115 | 0.00001 | 274 years | 2-W50-W64 | 2-Exposure to animate mechanical forces | 104 |
| 11 | 2-N17 | 2-Acute kidney failure | 4 | 5284 | 2025/06/23 to 2025/06/28 | 519 | 344.34 | 1.58 | 190.92 | 38.285122 | 0.00001 | 274 years | 2-N17-N19 | 2-Acute kidney failure and chronic kidney disease | 69 |
| 12 | 2-E86 | 2-Volume depletion | 4 | 2452 | 2025/06/23 to 2025/06/28 | 281 | 159.39 | 1.88 | 131.39 | 37.725494 | 0.00001 | 274 years | 2-E70-E88 | 2-Metabolic disorders | 35 |
| 13 | 2-N17.9 | 2-Acute kidney failure, unspecified | 5 | 5218 | 2025/06/23 to 2025/06/28 | 512 | 340.02 | 1.58 | 187.97 | 37.605543 | 0.00001 | 274 years | 2-N17 | 2-Acute kidney failure | 70 |
| 14 | 1-T67.01 | 1-Heatstroke and sunstroke | 7 | 35 | 2025/06/23 to 2025/06/28 | 25 | 2.57 | 36.22 | 24.31 | 34.483553 | 0.00001 | 274 years | 1-T67.0,2-T67.01 | 1-Heatstroke and sunstroke,2-Heatstroke and sunstroke | 26 |
| 15 | 2-E86.0 | 2-Dehydration | 5 | 2264 | 2025/06/23 to 2025/06/28 | 251 | 147.05 | 1.81 | 112.24 | 30.258656 | 0.00001 | 274 years | 2-E86 | 2-Volume depletion | 36 |
| 16 | 2-R50 | 2-Fever of other and unknown origin | 4 | 9286 | 2025/06/18 to 2025/06/28 | 1366 | 1105.78 | 1.29 | 305.39 | 28.496387 | 0.00001 | 274 years | 2-R50-R69 | 2-General symptoms and signs | 75 |
| 17 | 2-R50.9 | 2-Fever, unspecified | 5 | 9016 | 2025/06/18 to 2025/06/28 | 1331 | 1074.57 | 1.29 | 301.83 | 28.447640 | 0.00001 | 274 years | 2-R50 | 2-Fever of other and unknown origin | 76 |
| 18 | 2-L74.0 | 2-Miliaria rubra | 5 | 112 | 2025/06/20 to 2025/06/28 | 46 | 11.88 | 6.55 | 38.98 | 28.159526 | 0.00001 | 274 years | 2-L74 | 2-Eccrine sweat disorders | 64 |
| 19 | 2-H60 | 2-Otitis externa | 4 | 2146 | 2025/06/11 to 2025/06/28 | 579 | 417.50 | 1.53 | 200.36 | 27.854217 | 0.00001 | 274 years | 2-H60-H62 | 2-Diseases of external ear | 44 |
| 20 | 2-T67.1 | 2-Heat syncope | 5 | 110 | 2025/06/21 to 2025/06/28 | 40 | 9.35 | 6.15 | 33.50 | 27.506563 | 0.00001 | 274 years | 2-T67 | 2-Effects of heat and light | 99 |
| 21 | 2-L74 | 2-Eccrine sweat disorders | 4 | 139 | 2025/06/20 to 2025/06/28 | 49 | 14.52 | 5.12 | 39.43 | 25.123450 | 0.00001 | 274 years | 2-L60-L75 | 2-Disorders of skin appendages | 63 |
| 22 | 2-M79.89 | 2-Other specified soft tissue disorders | 6 | 4060 | 2025/06/06 to 2025/06/28 | 1224 | 1010.48 | 1.30 | 283.53 | 21.140132 | 0.00001 | 274 years | 2-M79.8 | 2-Other specified soft tissue disorders | 68 |
| 23 | 2-B34.1 | 2-Enterovirus infection, unspecified | 5 | 701 | 2025/06/07 to 2025/06/28 | 259 | 168.00 | 1.87 | 120.65 | 21.113116 | 0.00001 | 274 years | 2-B34,2-Other Enterovirus | 2-Viral infection of unspecified site,2-Other Enterovirus | 34 |
| 24 | 2-B08.5 | 2-Enteroviral vesicular pharyngitis | 5 | 299 | 2025/06/17 to 2025/06/28 | 85 | 39.16 | 2.67 | 53.18 | 20.034221 | 0.00001 | 274 years | 2-B08,2-Other Enterovirus | 2-Other viral infections characterized by skin and mucous membrane lesions, not elsewhere classified,2-Other Enterovirus | 33 |
| 25 | 2-M79.8 | 2-Other specified soft tissue disorders | 5 | 4098 | 2025/06/06 to 2025/06/28 | 1228 | 1020.02 | 1.29 | 276.23 | 19.914310 | 0.00001 | 274 years | 2-M79 | 2-Other and unspecified soft tissue disorders, not elsewhere classified | 67 |
| 26 | 2-R55 | 2-Syncope and collapse | 4 | 11998 | 2025/06/19 to 2025/06/28 | 1513 | 1285.69 | 1.21 | 261.14 | 19.029581 | 0.00001 | 274 years | 2-R50-R69 | 2-General symptoms and signs | 81 |
| 27 | 2-R41.8 | 2-Other symptoms and signs involving cognitive functions and awareness | 5 | 5028 | 2025/06/23 to 2025/06/28 | 433 | 321.46 | 1.37 | 116.28 | 17.440664 | 0.00001 | 274 years | 2-R41 | 2-Other symptoms and signs involving cognitive functions and awareness | 72 |
| 28 | 2-R41.82 | 2-Altered mental status, unspecified | 6 | 4732 | 2025/06/23 to 2025/06/28 | 409 | 303.49 | 1.37 | 111.00 | 16.528701 | 0.00001 | 274 years | 2-R41.8 | 2-Other symptoms and signs involving cognitive functions and awareness | 73 |
| 29 | 1-T67.5 | 1-Heat exhaustion, unspecified | 6 | 15 | 2025/06/24 to 2025/06/28 | 11 | 1.10 | 49.85 | 10.78 | 15.463901 | 0.00001 | 274 years | 1-T67,2-T67.5 | 1-Effects of heat and light,2-Heat exhaustion, unspecified | 28 |
| 30 | 2-R53 | 2-Malaise and fatigue | 4 | 7412 | 2025/06/23 to 2025/06/28 | 616 | 488.47 | 1.32 | 147.79 | 15.371062 | 0.00001 | 274 years | 2-R50-R69 | 2-General symptoms and signs | 77 |
| 31 | 2-R41 | 2-Other symptoms and signs involving cognitive functions and awareness | 4 | 5882 | 2025/06/21 to 2025/06/28 | 630 | 501.10 | 1.29 | 142.88 | 15.323118 | 0.00001 | 274 years | 2-R40-R46 | 2-Symptoms and signs involving cognition, perception, emotional state and behavior | 71 |
| 32 | 2-E87.1 | 2-Hypo-osmolality and hyponatremia | 5 | 1929 | 2025/06/21 to 2025/06/28 | 239 | 163.67 | 1.52 | 82.15 | 15.159563 | 0.00001 | 274 years | 2-E87 | 2-Other disorders of fluid, electrolyte and acid-base balance | 40 |
| 33 | 1-N17 | 1-Acute kidney failure | 5 | 3578 | 2025/06/23 to 2025/06/28 | 322 | 233.08 | 1.43 | 97.51 | 15.142849 | 0.00001 | 274 years | 1-N17-N19,2-N17 | 1-Acute kidney failure and chronic kidney disease,2-Acute kidney failure | 15 |
| 34 | 2-L55 | 2-Sunburn | 4 | 106 | 2025/06/20 to 2025/06/28 | 31 | 9.60 | 3.89 | 23.02 | 14.946308 | 0.00001 | 274 years | 2-L55-L59 | 2-Radiation-related disorders of the skin and subcutaneous tissue | 60 |
| 35 | 2-S91 | 2-Open wound of ankle, foot and toes | 4 | 1269 | 2025/06/18 to 2025/06/28 | 223 | 151.07 | 1.59 | 82.67 | 14.916589 | 0.00001 | 274 years | 2-S90-S99 | 2-Injuries to the ankle and foot | 86 |
| 36 | 2-R53.1 | 2-Weakness | 5 | 4974 | 2025/06/23 to 2025/06/28 | 431 | 328.01 | 1.38 | 117.87 | 14.704348 | 0.00001 | 274 years | 2-R53 | 2-Malaise and fatigue | 78 |
| 37 | 2-S81.81 | 2-Laceration without foreign body of lower leg | 6 | 667 | 2025/06/23 to 2025/06/28 | 82 | 42.22 | 2.03 | 41.63 | 14.650801 | 0.00001 | 274 years | 2-S81.8 | 2-Open wound of lower leg | 84 |
| 38 | 1-N17.9 | 1-Acute kidney failure, unspecified | 6 | 3521 | 2025/06/23 to 2025/06/28 | 316 | 229.35 | 1.43 | 95.02 | 14.630578 | 0.00001 | 274 years | 1-N17,2-N17.9 | 1-Acute kidney failure,2-Acute kidney failure, unspecified | 16 |
| 39 | 2-S81.8 | 2-Open wound of lower leg | 5 | 1121 | 2025/06/23 to 2025/06/28 | 122 | 72.12 | 1.77 | 53.08 | 14.257700 | 0.00001 | 274 years | 2-S81 | 2-Open wound of knee and lower leg | 82 |
| 40 | 2-L03 | 2-Cellulitis and acute lymphangitis | 4 | 8916 | 2025/06/11 to 2025/06/28 | 1964 | 1740.47 | 1.17 | 284.86 | 13.800424 | 0.00002 | 137 years | 2-L00-L08 | 2-Infections of the skin and subcutaneous tissue | 52 |
| 41 | 2-L55.9 | 2-Sunburn, unspecified | 5 | 84 | 2025/06/21 to 2025/06/28 | 25 | 7.04 | 4.56 | 19.52 | 13.713549 | 0.00002 | 137 years | 2-L55 | 2-Sunburn | 62 |
| 42 | 2-M62.82 | 2-Rhabdomyolysis | 6 | 554 | 2025/06/23 to 2025/06/28 | 69 | 35.18 | 2.06 | 35.53 | 12.663518 | 0.00003 | 91 years | 2-M62.8 | 2-Other specified disorders of muscle | 66 |
| 43 | 2-H60.50 | 2-Unspecified acute noninfective otitis externa | 6 | 782 | 2025/06/09 to 2025/06/28 | 240 | 170.93 | 1.58 | 88.26 | 12.384404 | 0.00006 | 46 years | 2-H60.5 | 2-Acute noninfective otitis externa | 50 |
| 44 | 2-E87 | 2-Other disorders of fluid, electrolyte and acid-base balance | 4 | 7779 | 2025/06/20 to 2025/06/28 | 882 | 745.65 | 1.20 | 149.20 | 11.778796 | 0.00012 | 23 years | 2-E70-E88 | 2-Metabolic disorders | 39 |
| 45 | 1-R41.8 | 1-Other symptoms and signs involving cognitive functions and awareness | 6 | 3007 | 2025/06/23 to 2025/06/28 | 262 | 191.30 | 1.38 | 72.67 | 11.703433 | 0.00014 | 20 years | 1-R41,2-R41.8 | 1-Other symptoms and signs involving cognitive functions and awareness,2-Other symptoms and signs involving cognitive functions and awareness | 18 |
| 46 | 1-R41.82 | 1-Altered mental status, unspecified | 7 | 2877 | 2025/06/23 to 2025/06/28 | 252 | 183.76 | 1.39 | 70.94 | 11.342890 | 0.00019 | 14 years | 1-R41.8,2-R41.82 | 1-Other symptoms and signs involving cognitive functions and awareness,2-Altered mental status, unspecified | 19 |
| 47 | 2-H60.9 | 2-Unspecified otitis externa | 5 | 735 | 2025/06/12 to 2025/06/28 | 193 | 134.44 | 1.59 | 71.58 | 11.226658 | 0.00021 | 13 years | 2-H60 | 2-Otitis externa | 51 |
| 48 | 2-L23.7 | 2-Allergic contact dermatitis due to plants, except food | 5 | 117 | 2025/06/02 to 2025/06/28 | 66 | 34.64 | 3.08 | 44.55 | 11.186537 | 0.00021 | 13 years | 2-L23 | 2-Allergic contact dermatitis | 59 |
| 49 | 2-H60.5 | 2-Acute noninfective otitis externa | 5 | 817 | 2025/06/09 to 2025/06/28 | 245 | 178.46 | 1.53 | 84.86 | 11.101681 | 0.00026 | 11 years | 2-H60 | 2-Otitis externa | 49 |
| 50 | 2-H60.3 | 2-Other infective otitis externa | 5 | 393 | 2025/06/27 to 2025/06/28 | 23 | 7.17 | 3.11 | 15.61 | 10.973759 | 0.00028 | 9.8 years | 2-H60 | 2-Otitis externa | 45 |
| 51 | 1-R50.9 | 1-Fever, unspecified | 6 | 1746 | 2025/06/16 to 2025/06/28 | 323 | 247.05 | 1.37 | 87.62 | 10.637271 | 0.00042 | 6.5 years | 1-R50,2-R50.9 | 1-Fever of other and unknown origin,2-Fever, unspecified | 21 |
| 52 | 1-R50 | 1-Fever of other and unknown origin | 5 | 1838 | 2025/06/15 to 2025/06/28 | 356 | 278.26 | 1.35 | 92.08 | 9.973026 | 0.00114 | 2.4 years | 1-R50-R69,2-R50 | 1-General symptoms and signs,2-Fever of other and unknown origin | 20 |
| 53 | 1-R41 | 1-Other symptoms and signs involving cognitive functions and awareness | 5 | 3416 | 2025/06/21 to 2025/06/28 | 365 | 286.50 | 1.29 | 81.83 | 9.890554 | 0.00124 | 2.2 years | 1-R40-R46,2-R41 | 1-Symptoms and signs involving cognition, perception, emotional state and behavior,2-Other symptoms and signs involving cognitive functions and awareness | 17 |
| 54 | 2-T67.8 | 2-Other effects of heat and light | 5 | 6 | 2025/06/19 to 2025/06/28 | 6 | 0.52 | infinity | 6.00 | 9.229156 | 0.00263 | 1.0 year | 2-T67 | 2-Effects of heat and light | 102 |
| 55 | 2-S91.31 | 2-Laceration without foreign body of foot | 6 | 359 | 2025/06/18 to 2025/06/28 | 73 | 42.80 | 1.90 | 34.62 | 8.774111 | 0.00463 | 216 days | 2-S91.3 | 2-Open wound of foot | 94 |
| 56 | 2-L03.90 | 2-Cellulitis, unspecified | 6 | 2389 | 2025/06/06 to 2025/06/28 | 701 | 596.29 | 1.25 | 140.92 | 8.704358 | 0.00495 | 202 days | 2-L03.9 | 2-Cellulitis and acute lymphangitis, unspecified | 58 |
| 57 | 1-E87.1 | 1-Hypo-osmolality and hyponatremia | 6 | 1275 | 2025/06/21 to 2025/06/28 | 152 | 107.44 | 1.46 | 47.73 | 8.175672 | 0.00914 | 109 days | 1-E87,2-E87.1 | 1-Other disorders of fluid, electrolyte and acid-base balance,2-Hypo-osmolality and hyponatremia | 6 |
| 58 | 2-L03.11 | 2-Cellulitis of other parts of limb | 6 | 3092 | 2025/06/11 to 2025/06/28 | 704 | 603.48 | 1.22 | 126.77 | 7.946791 | 0.01196 | 84 days | 2-L03.1 | 2-Cellulitis and acute lymphangitis of other parts of limb | 56 |
| 59 | 2-L03.1 | 2-Cellulitis and acute lymphangitis of other parts of limb | 5 | 3094 | 2025/06/11 to 2025/06/28 | 704 | 603.86 | 1.22 | 126.29 | 7.883940 | 0.01293 | 77 days | 2-L03 | 2-Cellulitis and acute lymphangitis | 55 |
| 60 | 2-S91.3 | 2-Open wound of foot | 5 | 729 | 2025/06/18 to 2025/06/28 | 126 | 87.10 | 1.56 | 45.08 | 7.624393 | 0.01748 | 57 days | 2-S91 | 2-Open wound of ankle, foot and toes | 93 |
| 61 | 2-H60.39 | 2-Other infective otitis externa | 6 | 237 | 2025/06/27 to 2025/06/28 | 14 | 4.23 | 3.14 | 9.55 | 6.982732 | 0.03778 | 26 days | 2-H60.3 | 2-Other infective otitis externa | 48 |
| 62 | 2-S91.2 | 2-Open wound of toe with damage to nail | 5 | 147 | 2025/06/19 to 2025/06/28 | 32 | 15.77 | 2.33 | 18.24 | 6.413791 | 0.07334 | 14 days | 2-S91 | 2-Open wound of ankle, foot and toes | 91 |
| 63 | 2-S91.20 | 2-Unspecified open wound of toe with damage to nail | 6 | 125 | 2025/06/19 to 2025/06/28 | 28 | 13.29 | 2.41 | 16.39 | 6.160817 | 0.09774 | 10 days | 2-S91.2 | 2-Open wound of toe with damage to nail | 92 |
| 64 | 2-H60.33 | 2-Swimmer's ear | 6 | 53 | 2025/06/26 to 2025/06/28 | 8 | 1.72 | 5.75 | 6.61 | 6.013661 | 0.11582 | 9 days | 2-H60.3 | 2-Other infective otitis externa | 47 |
| 65 | 2-S91.33 | 2-Puncture wound without foreign body of foot | 6 | 142 | 2025/06/21 to 2025/06/28 | 25 | 11.84 | 2.30 | 14.13 | 5.524635 | 0.19513 | 5 days | 2-S91.3 | 2-Open wound of foot | 95 |
| 66 | 1-M62.82 | 1-Rhabdomyolysis | 7 | 334 | 2025/06/23 to 2025/06/28 | 37 | 20.75 | 1.80 | 16.50 | 5.152454 | 0.28592 | 3 days | 1-M62.8,2-M62.82 | 1-Other specified disorders of muscle,2-Rhabdomyolysis | 12 |
| 67 | 2-T67.2 | 2-Heat cramp | 5 | 3 | 2025/06/23 to 2025/06/28 | 3 | 0.23 | infinity | 3.00 | 4.956953 | 0.35509 | 3 days | 2-T67 | 2-Effects of heat and light | 100 |
| 68 | 2-S81.80 | 2-Unspecified open wound of lower leg | 6 | 348 | 2025/06/16 to 2025/06/28 | 73 | 49.73 | 1.60 | 27.49 | 4.753251 | 0.42778 | 2 days | 2-S81.8 | 2-Open wound of lower leg | 83 |
| 69 | 1-B08.4 | 1-Enteroviral vesicular stomatitis with exanthem | 6 | 8 | 2025/06/15 to 2025/06/28 | 6 | 1.24 | 16.84 | 5.64 | 4.715100 | 0.44121 | 2 days | 1-B08,1-Other Enterovirus,2-B08.4 | 1-Other viral infections characterized by skin and mucous membrane lesions, not elsewhere classified,1-Other Enterovirus,2-Enteroviral vesicular stomatitis with exanthem | 2 |
| 70 | 1-W57 | 1-Bitten or stung by nonvenomous insect and other nonvenomous arthropods | 5 | 11 | 2025/06/15 to 2025/06/28 | 7 | 1.71 | 9.82 | 6.29 | 4.563453 | 0.49526 | 2 days | 1-W50-W64,2-W57 | 1-Exposure to animate mechanical forces,2-Bitten or stung by nonvenomous insect and other nonvenomous arthropods | 29 |
| 71 | 2-R53.83 | 2-Other fatigue | 6 | 1726 | 2025/06/25 to 2025/06/28 | 101 | 73.88 | 1.44 | 30.94 | 4.459116 | 0.53454 | 2 days | 2-R53.8 | 2-Other malaise and fatigue | 80 |
| 72 | 2-S91.01 | 2-Laceration without foreign body of ankle | 6 | 73 | 2025/06/18 to 2025/06/28 | 19 | 9.07 | 2.62 | 11.75 | 4.116207 | 0.69042 | 1 day | 2-S91.0 | 2-Open wound of ankle | 88 |
| 73 | 2-S91.0 | 2-Open wound of ankle | 5 | 156 | 2025/06/20 to 2025/06/28 | 28 | 15.48 | 2.06 | 14.38 | 4.075143 | 0.70626 | 1 day | 2-S91 | 2-Open wound of ankle, foot and toes | 87 |
| 74 | 1-R55 | 1-Syncope and collapse | 5 | 3066 | 2025/06/24 to 2025/06/28 | 198 | 161.37 | 1.25 | 39.87 | 3.875467 | 0.78236 | 1 day | 1-R50-R69,2-R55 | 1-General symptoms and signs,2-Syncope and collapse | 24 |
| 75 | 2-E86.1 | 2-Hypovolemia | 5 | 101 | 2025/06/24 to 2025/06/28 | 13 | 5.49 | 2.68 | 8.15 | 3.694184 | 0.85001 | 1 day | 2-E86 | 2-Volume depletion | 37 |
| 76 | 2-B08.1 | 2-Molluscum contagiosum | 5 | 76 | 2025/06/27 to 2025/06/28 | 6 | 1.58 | 4.29 | 4.60 | 3.593200 | 0.88055 | 1 day | 2-B08 | 2-Other viral infections characterized by skin and mucous membrane lesions, not elsewhere classified | 31 |
| 77 | 1-R53.1 | 1-Weakness | 6 | 2001 | 2025/06/23 to 2025/06/28 | 164 | 132.21 | 1.29 | 37.23 | 3.549196 | 0.89370 | 1 day | 1-R53,2-R53.1 | 1-Malaise and fatigue,2-Weakness | 23 |
| 78 | 1-M79.89 | 1-Other specified soft tissue disorders | 7 | 724 | 2025/06/11 to 2025/06/28 | 173 | 140.45 | 1.30 | 39.76 | 3.511003 | 0.90357 | 1 day | 1-M79.8,2-M79.89 | 1-Other specified soft tissue disorders,2-Other specified soft tissue disorders | 14 |
| 79 | 2-H60.31 | 2-Diffuse otitis externa | 6 | 97 | 2025/06/20 to 2025/06/28 | 18 | 8.97 | 2.14 | 9.59 | 3.510139 | 0.90384 | 1 day | 2-H60.3 | 2-Other infective otitis externa | 46 |
| 80 | 2-S91.11 | 2-Laceration without foreign body of toe without damage to nail | 6 | 158 | 2025/06/24 to 2025/06/28 | 16 | 7.72 | 2.04 | 8.17 | 3.377562 | 0.93378 | 1 day | 2-S91.1 | 2-Open wound of toe without damage to nail | 90 |
| 81 | 1-B08 | 1-Other viral infections characterized by skin and mucous membrane lesions, not elsewhere classified | 5 | 11 | 2025/06/15 to 2025/06/28 | 6 | 1.67 | 6.73 | 5.11 | 3.341275 | 0.93997 | 1 day | 1-B00-B09,2-B08 | 1-Viral infections characterized by skin and mucous membrane lesions,2-Other viral infections characterized by skin and mucous membrane lesions, not elsewhere classified | 1 |
| 82 | 1-M79.8 | 1-Other specified soft tissue disorders | 6 | 735 | 2025/06/11 to 2025/06/28 | 174 | 142.56 | 1.28 | 38.34 | 3.237271 | 0.96134 | 1 day | 1-M79,2-M79.8 | 1-Other and unspecified soft tissue disorders, not elsewhere classified,2-Other specified soft tissue disorders | 13 |
| 83 | 2-E87.6 | 2-Hypokalemia | 5 | 1716 | 2025/06/20 to 2025/06/28 | 197 | 163.94 | 1.22 | 35.42 | 3.130485 | 0.97444 | 1 day | 2-E87 | 2-Other disorders of fluid, electrolyte and acid-base balance | 41 |
| 84 | 1-E86 | 1-Volume depletion | 5 | 752 | 2025/06/24 to 2025/06/28 | 57 | 40.27 | 1.49 | 18.67 | 3.073322 | 0.98007 | 1 day | 1-E70-E88,2-E86 | 1-Metabolic disorders,2-Volume depletion | 3 |
| 85 | 2-E87.70 | 2-Fluid overload, unspecified | 6 | 638 | 2025/06/18 to 2025/06/28 | 96 | 74.08 | 1.32 | 23.25 | 2.963095 | 0.98760 | 1 day | 2-E87.7 | 2-Fluid overload | 43 |
| 86 | 1-T67.1 | 1-Heat syncope | 6 | 28 | 2025/06/21 to 2025/06/28 | 7 | 2.43 | 3.59 | 5.05 | 2.839945 | 0.99382 | 1 day | 1-T67,2-T67.1 | 1-Effects of heat and light,2-Heat syncope | 27 |
| 87 | 2-L03.0 | 2-Cellulitis and acute lymphangitis of finger and toe | 5 | 1592 | 2025/06/27 to 2025/06/28 | 45 | 31.13 | 1.46 | 14.11 | 2.714356 | 0.99716 | 1 day | 2-L03 | 2-Cellulitis and acute lymphangitis | 53 |
| 88 | 2-E87.7 | 2-Fluid overload | 5 | 741 | 2025/06/18 to 2025/06/28 | 109 | 86.49 | 1.28 | 24.17 | 2.704547 | 0.99742 | 1 day | 2-E87 | 2-Other disorders of fluid, electrolyte and acid-base balance | 42 |
| 89 | 1-R53 | 1-Malaise and fatigue | 5 | 2579 | 2025/06/23 to 2025/06/28 | 202 | 171.02 | 1.23 | 37.97 | 2.651245 | 0.99809 | 1 day | 1-R50-R69,2-R53 | 1-General symptoms and signs,2-Malaise and fatigue | 22 |
| 90 | 1-E86.1 | 1-Hypovolemia | 6 | 71 | 2025/06/24 to 2025/06/28 | 9 | 3.76 | 2.63 | 5.58 | 2.611426 | 0.99852 | 1 day | 1-E86,2-E86.1 | 1-Volume depletion,2-Hypovolemia | 4 |
| 91 | 2-L03.317 | 2-Cellulitis of buttock | 7 | 185 | 2025/06/25 to 2025/06/28 | 15 | 7.86 | 2.05 | 7.67 | 2.552336 | 0.99889 | 1 day | 2-L03.31 | 2-Cellulitis of trunk | 57 |
| 92 | 2-L03.01 | 2-Cellulitis of finger | 6 | 1119 | 2025/06/28 to 2025/06/28 | 18 | 10.08 | 1.76 | 7.76 | 2.516054 | 0.99932 | 1 day | 2-L03.0 | 2-Cellulitis and acute lymphangitis of finger and toe | 54 |
| 93 | 2-L55.0 | 2-Sunburn of first degree | 5 | 18 | 2025/06/03 to 2025/06/28 | 11 | 5.14 | 3.98 | 8.24 | 2.506756 | 0.99934 | 1 day | 2-L55 | 2-Sunburn | 61 |
| 94 | 2-S91.1 | 2-Open wound of toe without damage to nail | 5 | 237 | 2025/06/24 to 2025/06/28 | 20 | 11.71 | 1.67 | 8.03 | 2.415958 | 0.99968 | 1 day | 2-S91 | 2-Open wound of ankle, foot and toes | 89 |
| 95 | 1-E87 | 1-Other disorders of fluid, electrolyte and acid-base balance | 5 | 4630 | 2025/06/20 to 2025/06/28 | 487 | 440.23 | 1.11 | 46.36 | 2.402393 | 0.99974 | 1 day | 1-E70-E88,2-E87 | 1-Metabolic disorders,2-Other disorders of fluid, electrolyte and acid-base balance | 5 |
| 96 | 1-L03.90 | 1-Cellulitis, unspecified | 7 | 678 | 2025/06/26 to 2025/06/28 | 32 | 21.16 | 1.60 | 12.03 | 2.397069 | 0.99975 | 1 day | 1-L03.9,2-L03.90 | 1-Cellulitis and acute lymphangitis, unspecified,2-Cellulitis, unspecified | 11 |
| 97 | 2-E86.9 | 2-Volume depletion, unspecified | 5 | 27 | 2025/06/06 to 2025/06/28 | 13 | 6.62 | 2.80 | 8.35 | 2.392062 | 0.99976 | 1 day | 2-E86 | 2-Volume depletion | 38 |
| 98 | 1-L03.311 | 1-Cellulitis of abdominal wall | 8 | 27 | 2025/06/21 to 2025/06/28 | 6 | 2.17 | 3.08 | 4.05 | 2.272754 | 0.99994 | 1 day | 1-L03.31,2-L03.311 | 1-Cellulitis of trunk,2-Cellulitis of abdominal wall | 10 |
| 99 | 2-R53.8 | 2-Other malaise and fatigue | 5 | 2347 | 2025/06/25 to 2025/06/28 | 122 | 100.17 | 1.27 | 26.06 | 2.223041 | 0.99997 | 1 day | 2-R53 | 2-Malaise and fatigue | 79 |
| 100 | 1-H60.9 | 1-Unspecified otitis externa | 6 | 8 | 2025/06/16 to 2025/06/28 | 4 | 1.12 | 6.04 | 3.34 | 2.212166 | 0.99997 | 1 day | 1-H60,2-H60.9 | 1-Otitis externa,2-Unspecified otitis externa | 9 |
| 101 | 1-E87.70 | 1-Fluid overload, unspecified | 7 | 456 | 2025/06/18 to 2025/06/28 | 69 | 52.97 | 1.33 | 17.05 | 2.212158 | 0.99997 | 1 day | 1-E87.7,2-E87.70 | 1-Fluid overload,2-Fluid overload, unspecified | 8 |
| 102 | 2-R41.89 | 2-Other symptoms and signs involving cognitive functions and awareness | 6 | 275 | 2025/06/24 to 2025/06/28 | 22 | 13.58 | 1.58 | 8.04 | 2.191546 | 0.99997 | 1 day | 2-R41.8 | 2-Other symptoms and signs involving cognitive functions and awareness | 74 |
| 103 | 2-S81.85 | 2-Open bite of lower leg | 6 | 79 | 2025/06/16 to 2025/06/28 | 19 | 11.29 | 1.91 | 9.07 | 2.178469 | 0.99998 | 1 day | 2-S81.8 | 2-Open wound of lower leg | 85 |
| 104 | 2-L74.3 | 2-Miliaria, unspecified | 5 | 14 | 2025/06/14 to 2025/06/28 | 6 | 2.27 | 3.91 | 4.47 | 2.100153 | 0.99999 | 1 day | 2-L74 | 2-Eccrine sweat disorders | 65 |
| 105 | 1-E87.21 | 1-Acute metabolic acidosis | 7 | 18 | 2025/06/23 to 2025/06/28 | 4 | 1.17 | 4.14 | 3.03 | 2.093564 | 0.99999 | 1 day | 1-E87.2,2-E87.21 | 1-Acidosis,2-Acute metabolic acidosis | 7 |

|  |  |  |  |  |  |  |  |  |  |  |  |  |  |  |  |
| --- | --- | --- | --- | --- | --- | --- | --- | --- | --- | --- | --- | --- | --- | --- | --- |
| 106 | 2-R41.9 | 2-Unspecified symptoms and signs involving cognitive functions and awareness | 5 | 42 | 2025/06/27 to 2025/06/28 | 3 | 0.72 | 3.85 | 2.22 | 2.008207 | 1.00000 | 1 day | 2-R41 | 2-Other symptoms and signs involving cognitive functions and awareness | 0 |
| 107 | 2-L03.818 | 2-Cellulitis of other sites | 7 | 112 | 2025/06/20 to 2025/06/28 | 18 | 10.81 | 1.80 | 8.00 | 1.985883 | 1.00000 | 1 day | 2-L03.81 | 2-Cellulitis of other sites | 0 |
| 108 | 1-H60 | 1-Otitis externa | 5 | 35 | 2025/06/26 to 2025/06/28 | 4 | 1.22 | 4.17 | 3.04 | 1.971783 | 1.00000 | 1 day | 1-H60-H62,2-H60 | 1-Diseases of external ear,2-Otitis externa | 0 |
| 109 | 2-H60.6 | 2-Unspecified chronic otitis externa | 5 | 20 | 2025/06/15 to 2025/06/28 | 7 | 3.00 | 3.02 | 4.68 | 1.930382 | 1.00000 | 1 day | 2-H60 | 2-Otitis externa | 0 |
| 110 | 2-L03.81 | 2-Cellulitis of other sites | 6 | 147 | 2025/06/20 to 2025/06/28 | 22 | 14.06 | 1.65 | 8.70 | 1.907787 | 1.00000 | 1 day | 2-L03.8 | 2-Cellulitis and acute lymphangitis of other sites | 0 |
| 111 | 1-L03.818 | 1-Cellulitis of other sites | 8 | 16 | 2025/06/19 to 2025/06/28 | 5 | 1.80 | 3.80 | 3.68 | 1.902194 | 1.00000 | 1 day | 1-L03.81,2-L03.818 | 1-Cellulitis of other sites,2-Cellulitis of other sites | 0 |
| 112 | 2-B97.11 | 2-Coxsackievirus as the cause of diseases classified elsewhere | 5 | 27 | 2025/06/13 to 2025/06/28 | 10 | 5.15 | 2.84 | 6.48 | 1.787560 | 1.00000 | 1 day | 2-B97.1,2-Coxsackievirus | 2-Enterovirus as the cause of diseases classified elsewhere,2-Coxsackievirus | 0 |
| 113 | 1-S81.81 | 1-Laceration without foreign body of lower leg | 7 | 21 | 2025/06/07 to 2025/06/28 | 10 | 5.15 | 2.90 | 6.56 | 1.787209 | 1.00000 | 1 day | 1-S81.8,2-S81.81 | 1-Open wound of lower leg,2-Laceration without foreign body of lower leg | 0 |
| 114 | 2-L03.31 | 2-Cellulitis of trunk | 6 | 474 | 2025/06/19 to 2025/06/28 | 65 | 50.94 | 1.33 | 16.06 | 1.783924 | 1.00000 | 1 day | 2-L03.3 | 2-Cellulitis and acute lymphangitis of trunk | 0 |
| 115 | 1-L03.81 | 1-Cellulitis of other sites | 7 | 23 | 2025/06/19 to 2025/06/28 | 6 | 2.48 | 2.95 | 3.97 | 1.782556 | 1.00000 | 1 day | 1-L03.8,2-L03.81 | 1-Cellulitis and acute lymphangitis of other sites,2-Cellulitis of other sites | 0 |
| 116 | 2-E87.21 | 2-Acute metabolic acidosis | 6 | 21 | 2025/06/23 to 2025/06/28 | 4 | 1.32 | 3.41 | 2.83 | 1.756197 | 1.00000 | 1 day | 2-E87.2 | 2-Acidosis | 0 |
| 117 | 2-L03.3 | 2-Cellulitis and acute lymphangitis of trunk | 5 | 476 | 2025/06/19 to 2025/06/28 | 65 | 51.09 | 1.32 | 15.82 | 1.742278 | 1.00000 | 1 day | 2-L03 | 2-Cellulitis and acute lymphangitis | 0 |
| 118 | 2-E87.0 | 2-Hyperosmolality and hypernatremia | 5 | 310 | 2025/06/23 to 2025/06/28 | 30 | 20.91 | 1.55 | 10.67 | 1.739484 | 1.00000 | 1 day | 2-E87 | 2-Other disorders of fluid, electrolyte and acid-base balance | 0 |
| 119 | 2-E87.5 | 2-Hyperkalemia | 5 | 1855 | 2025/06/06 to 2025/06/28 | 496 | 458.81 | 1.10 | 44.92 | 1.468650 | 1.00000 | 1 day | 2-E87 | 2-Other disorders of fluid, electrolyte and acid-base balance | 0 |
| 120 | 2-S91.21 | 2-Laceration without foreign body of toe with damage to nail | 6 | 20 | 2025/06/22 to 2025/06/28 | 4 | 1.50 | 3.10 | 2.71 | 1.429120 | 1.00000 | 1 day | 2-S91.2 | 2-Open wound of toe with damage to nail | 0 |
| 121 | 2-B08.3 | 2-Erythema infectiosum [fifth disease] | 5 | 24 | 2025/06/20 to 2025/06/28 | 5 | 2.11 | 2.47 | 2.98 | 1.420824 | 1.00000 | 1 day | 2-B08 | 2-Other viral infections characterized by skin and mucous membrane lesions, not elsewhere classified | 0 |
| 122 | 1-E87.7 | 1-Fluid overload | 6 | 534 | 2025/06/18 to 2025/06/28 | 76 | 62.34 | 1.24 | 14.52 | 1.398330 | 1.00000 | 1 day | 1-E87,2-E87.7 | 1-Other disorders of fluid, electrolyte and acid-base balance,2-Fluid overload | 0 |
| 123 | 1-S81.8 | 1-Open wound of lower leg | 6 | 116 | 2025/06/07 to 2025/06/28 | 37 | 27.84 | 1.50 | 12.26 | 1.364130 | 1.00000 | 1 day | 1-S81,2-S81.8 | 1-Open wound of knee and lower leg,2-Open wound of lower leg | 0 |
| 124 | 2-R50.83 | 2-Postvaccination fever | 6 | 57 | 2025/06/07 to 2025/06/28 | 20 | 13.49 | 1.73 | 8.41 | 1.363339 | 1.00000 | 1 day | 2-R50.8 | 2-Other specified fever | 0 |
| 125 | 2-S91.12 | 2-Laceration with foreign body of toe without damage to nail | 6 | 5 | 2025/06/09 to 2025/06/28 | 3 | 1.01 | 5.36 | 2.44 | 1.273732 | 1.00000 | 1 day | 2-S91.1 | 2-Open wound of toe without damage to nail | 0 |
| 126 | 1-R41.9 | 1-Unspecified symptoms and signs involving cognitive functions and awareness | 6 | 9 | 2025/06/19 to 2025/06/28 | 3 | 1.04 | 4.18 | 2.28 | 1.226298 | 1.00000 | 1 day | 1-R41,2-R41.9 | 1-Other symptoms and signs involving cognitive functions and awareness,2-Unspecified symptoms and signs involving cognitive functions and awareness | 0 |
| 127 | 2-H60.0 | 2-Abscess of external ear | 5 | 97 | 2025/06/13 to 2025/06/28 | 23 | 16.36 | 1.50 | 7.67 | 1.193595 | 1.00000 | 1 day | 2-H60 | 2-Otitis externa | 0 |
| 128 | 2-R41.3 | 2-Other amnesia | 5 | 180 | 2025/06/02 to 2025/06/28 | 65 | 53.33 | 1.34 | 16.63 | 1.193400 | 1.00000 | 1 day | 2-R41 | 2-Other symptoms and signs involving cognitive functions and awareness | 0 |
| 129 | 2-L74.5 | 2-Focal hyperhidrosis | 5 | 6 | 2025/06/04 to 2025/06/28 | 4 | 1.68 | 5.37 | 3.26 | 1.146669 | 1.00000 | 1 day | 2-L74 | 2-Eccrine sweat disorders | 0 |
| 130 | 1-L03 | 1-Cellulitis and acute lymphangitis | 5 | 2111 | 2025/06/20 to 2025/06/28 | 224 | 202.55 | 1.12 | 23.26 | 1.098079 | 1.00000 | 1 day | 1-L00-L08,2-L03 | 1-Infections of the skin and subcutaneous tissue,2-Cellulitis and acute lymphangitis | 0 |
| 131 | 1-L03.31 | 1-Cellulitis of trunk | 7 | 114 | 2025/06/22 to 2025/06/28 | 13 | 8.46 | 1.60 | 4.85 | 1.044838 | 1.00000 | 1 day | 1-L03.3,2-L03.31 | 1-Cellulitis and acute lymphangitis of trunk,2-Cellulitis of trunk | 0 |
| 132 | 2-L03.314 | 2-Cellulitis of groin | 7 | 45 | 2025/06/18 to 2025/06/28 | 9 | 5.35 | 1.86 | 4.17 | 1.033094 | 1.00000 | 1 day | 2-L03.31 | 2-Cellulitis of trunk | 0 |
| 133 | 1-L03.3 | 1-Cellulitis and acute lymphangitis of trunk | 6 | 115 | 2025/06/22 to 2025/06/28 | 13 | 8.54 | 1.58 | 4.77 | 1.004788 | 1.00000 | 1 day | 1-L03,2-L03.3 | 1-Cellulitis and acute lymphangitis,2-Cellulitis and acute lymphangitis of trunk | 0 |
| 134 | 1-S91.33 | 1-Puncture wound without foreign body of foot | 7 | 8 | 2025/06/09 to 2025/06/28 | 4 | 1.80 | 3.57 | 2.88 | 0.993090 | 1.00000 | 1 day | 1-S91.3,2-S91.33 | 1-Open wound of foot,2-Puncture wound without foreign body of foot | 0 |
| 135 | 1-R41.89 | 1-Other symptoms and signs involving cognitive functions and awareness | 7 | 125 | 2025/06/22 to 2025/06/28 | 14 | 9.47 | 1.56 | 5.04 | 0.943503 | 1.00000 | 1 day | 1-R41.8,2-R41.89 | 1-Other symptoms and signs involving cognitive functions and awareness,2-Other symptoms and signs involving cognitive functions and awareness | 0 |
| 136 | 1-E87.0 | 1-Hyperosmolality and hypernatremia | 6 | 257 | 2025/06/24 to 2025/06/28 | 18 | 12.92 | 1.37 | 4.81 | 0.889475 | 1.00000 | 1 day | 1-E87,2-E87.0 | 1-Other disorders of fluid, electrolyte and acid-base balance,2-Hyperosmolality and hypernatremia | 0 |
| 137 | 1-E87.6 | 1-Hypokalemia | 6 | 641 | 2025/06/04 to 2025/06/28 | 191 | 173.50 | 1.14 | 23.54 | 0.854523 | 1.00000 | 1 day | 1-E87,2-E87.6 | 1-Other disorders of fluid, electrolyte and acid-base balance,2-Hypokalemia | 0 |
| 138 | 1-E86.0 | 1-Dehydration | 6 | 643 | 2025/06/24 to 2025/06/28 | 42 | 34.33 | 1.27 | 8.85 | 0.798969 | 1.00000 | 1 day | 1-E86,2-E86.0 | 1-Volume depletion,2-Dehydration | 0 |
| 139 | 1-L03.314 | 1-Cellulitis of groin | 8 | 18 | 2025/06/22 to 2025/06/28 | 3 | 1.33 | 2.48 | 1.79 | 0.764948 | 1.00000 | 1 day | 1-L03.31,2-L03.314 | 1-Cellulitis of trunk,2-Cellulitis of groin | 0 |
| 140 | 2-H60.1 | 2-Cellulitis of external ear | 5 | 60 | 2025/06/15 to 2025/06/28 | 13 | 9.04 | 1.55 | 4.63 | 0.763509 | 1.00000 | 1 day | 2-H60 | 2-Otitis externa | 0 |
| 141 | 2-L03.03 | 2-Cellulitis of toe | 6 | 471 | 2025/06/01 to 2025/06/28 | 158 | 143.59 | 1.14 | 19.97 | 0.700553 | 1.00000 | 1 day | 2-L03.0 | 2-Cellulitis and acute lymphangitis of finger and toe | 0 |
| 142 | 1-R53.83 | 1-Other fatigue | 7 | 292 | 2025/06/10 to 2025/06/28 | 71 | 61.53 | 1.24 | 13.59 | 0.693995 | 1.00000 | 1 day | 1-R53.8,2-R53.83 | 1-Other malaise and fatigue,2-Other fatigue | 0 |
| 143 | 2-L03.313 | 2-Cellulitis of chest wall | 7 | 39 | 2025/06/02 to 2025/06/28 | 16 | 11.76 | 1.65 | 6.33 | 0.686856 | 1.00000 | 1 day | 2-L03.31 | 2-Cellulitis of trunk | 0 |
| 144 | 2-L03.211 | 2-Cellulitis of face | 7 | 395 | 2025/06/28 to 2025/06/28 | 6 | 3.57 | 1.66 | 2.38 | 0.681912 | 1.00000 | 1 day | 2-L03.21 | 2-Cellulitis and acute lymphangitis of face | 0 |
| 145 | 1-H60.2 | 1-Malignant otitis externa | 6 | 5 | 2025/06/03 to 2025/06/28 | 3 | 1.42 | 3.80 | 2.21 | 0.664730 | 1.00000 | 1 day | 1-H60,2-H60.2 | 1-Otitis externa,2-Malignant otitis externa | 0 |
| 146 | 2-S91.00 | 2-Unspecified open wound of ankle | 6 | 56 | 2025/06/01 to 2025/06/28 | 22 | 17.06 | 1.47 | 7.01 | 0.655113 | 1.00000 | 1 day | 2-S91.0 | 2-Open wound of ankle | 0 |
| 147 | 1-Other Enterovirus |  | 5 | 49 | 2025/06/07 to 2025/06/28 | 16 | 11.85 | 1.55 | 5.67 | 0.652633 | 1.00000 | 1 day | 1-dummy2,2-Other Enterovirus |  | 0 |
| 148 | 1-S81.80 | 1-Unspecified open wound of lower leg | 7 | 87 | 2025/06/08 to 2025/06/28 | 25 | 19.75 | 1.36 | 6.64 | 0.643217 | 1.00000 | 1 day | 1-S81.8,2-S81.80 | 1-Open wound of lower leg,2-Unspecified open wound of lower leg | 0 |
| 149 | 2-L74.51 | 2-Primary focal hyperhidrosis | 6 | 5 | 2025/06/04 to 2025/06/28 | 3 | 1.45 | 4.03 | 2.26 | 0.630606 | 1.00000 | 1 day | 2-L74.5 | 2-Focal hyperhidrosis | 0 |
| 150 | 2-L03.311 | 2-Cellulitis of abdominal wall | 7 | 117 | 2025/06/19 to 2025/06/28 | 17 | 12.89 | 1.42 | 5.03 | 0.595391 | 1.00000 | 1 day | 2-L03.31 | 2-Cellulitis of trunk | 0 |
| 151 | 1-N17.0 | 1-Acute kidney failure with tubular necrosis | 6 | 28 | 2025/06/24 to 2025/06/28 | 3 | 1.52 | 2.17 | 1.62 | 0.556753 | 1.00000 | 1 day | 1-N17,2-N17.0 | 1-Acute kidney failure,2-Acute kidney failure with tubular necrosis | 0 |
| 152 | 1-N17.8 | 1-Other acute kidney failure | 6 | 22 | 2025/06/23 to 2025/06/28 | 3 | 1.55 | 2.29 | 1.69 | 0.534431 | 1.00000 | 1 day | 1-N17,2-N17.8 | 1-Acute kidney failure,2-Other acute kidney failure | 0 |
| 153 | 1-R41.3 | 1-Other amnesia | 6 | 55 | 2025/06/01 to 2025/06/28 | 21 | 16.72 | 1.40 | 6.01 | 0.505665 | 1.00000 | 1 day | 1-R41,2-R41.3 | 1-Other symptoms and signs involving cognitive functions and awareness,2-Other amnesia | 0 |
| 154 | 2-R41.840 | 2-Attention and concentration deficit | 7 | 13 | 2025/06/08 to 2025/06/28 | 5 | 3.08 | 2.11 | 2.63 | 0.501017 | 1.00000 | 1 day | 2-R41.84 | 2-Other specified cognitive deficit | 0 |
| 155 | 2-R50.8 | 2-Other specified fever | 5 | 268 | 2025/06/10 to 2025/06/28 | 62 | 54.51 | 1.16 | 8.49 | 0.492361 | 1.00000 | 1 day | 2-R50 | 2-Fever of other and unknown origin | 0 |
| 156 | 2-E87.8 | 2-Other disorders of electrolyte and fluid balance, not elsewhere classified | 5 | 214 | 2025/06/27 to 2025/06/28 | 6 | 3.94 | 1.44 | 1.84 | 0.461279 | 1.00000 | 1 day | 2-E87 | 2-Other disorders of fluid, electrolyte and acid-base balance | 0 |
| 157 | 2-R50.81 | 2-Fever presenting with conditions classified elsewhere | 6 | 181 | 2025/06/28 to 2025/06/28 | 3 | 1.64 | 1.81 | 1.34 | 0.448109 | 1.00000 | 1 day | 2-R50.8 | 2-Other specified fever | 0 |
| 158 | 2-L03.312 | 2-Cellulitis of back [any part except buttock] | 7 | 39 | 2025/06/08 to 2025/06/28 | 12 | 9.02 | 1.50 | 4.00 | 0.445733 | 1.00000 | 1 day | 2-L03.31 | 2-Cellulitis of trunk | 0 |
| 159 | 1-H60.50 | 1-Unspecified acute noninfective otitis externa | 7 | 8 | 2025/06/02 to 2025/06/28 | 4 | 2.40 | 2.38 | 2.32 | 0.444784 | 1.00000 | 1 day | 1-H60.5,2-H60.50 | 1-Acute noninfective otitis externa,2-Unspecified acute noninfective otitis externa | 0 |
| 160 | 2-L03.213 | 2-Periorbital cellulitis | 7 | 751 | 2025/06/15 to 2025/06/28 | 124 | 113.85 | 1.11 | 12.29 | 0.439788 | 1.00000 | 1 day | 2-L03.21 | 2-Cellulitis and acute lymphangitis of face | 0 |
| 161 | 2-N17.0 | 2-Acute kidney failure with tubular necrosis | 5 | 30 | 2025/06/24 to 2025/06/28 | 3 | 1.67 | 2.01 | 1.51 | 0.430206 | 1.00000 | 1 day | 2-N17 | 2-Acute kidney failure | 0 |
| 162 | 1-E87.20 | 1-Acidosis, unspecified | 7 | 462 | 2025/06/10 to 2025/06/28 | 104 | 95.03 | 1.12 | 11.00 | 0.410484 | 1.00000 | 1 day | 1-E87.2,2-E87.20 | 1-Acidosis,2-Acidosis, unspecified | 0 |
| 163 | 1-L03.2 | 1-Cellulitis and acute lymphangitis of face and neck | 6 | 177 | 2025/06/19 to 2025/06/28 | 22 | 18.03 | 1.19 | 3.45 | 0.408820 | 1.00000 | 1 day | 1-L03,2-L03.2 | 1-Cellulitis and acute lymphangitis,2-Cellulitis and acute lymphangitis of face and neck | 0 |
| 164 | 2-E87.20 | 2-Acidosis, unspecified | 6 | 609 | 2025/06/28 to 2025/06/28 | 8 | 5.79 | 1.43 | 2.41 | 0.376035 | 1.00000 | 1 day | 2-E87.2 | 2-Acidosis | 0 |
| 165 | 2-N17.8 | 2-Other acute kidney failure | 5 | 26 | 2025/06/23 to 2025/06/28 | 3 | 1.77 | 1.89 | 1.41 | 0.356302 | 1.00000 | 1 day | 2-N17 | 2-Acute kidney failure | 0 |
| 166 | 1-L03.211 | 1-Cellulitis of face | 8 | 86 | 2025/06/25 to 2025/06/28 | 5 | 3.37 | 1.43 | 1.51 | 0.342291 | 1.00000 | 1 day | 1-L03.21,2-L03.211 | 1-Cellulitis and acute lymphangitis of face,2-Cellulitis of face | 0 |
| 167 | 2-L03.811 | 2-Cellulitis of head [any part, except face] | 7 | 35 | 2025/06/04 to 2025/06/28 | 12 | 9.44 | 1.40 | 3.44 | 0.319875 | 1.00000 | 1 day | 2-L03.81 | 2-Cellulitis of other sites | 0 |
| 168 | 1-R53.8 | 1-Other malaise and fatigue | 6 | 521 | 2025/06/04 to 2025/06/28 | 152 | 142.57 | 1.11 | 14.68 | 0.305116 | 1.00000 | 1 day | 1-R53,2-R53.8 | 1-Malaise and fatigue,2-Other malaise and fatigue | 0 |
| 169 | 2-B08.2 | 2-Exanthema subitum [sixth disease] | 5 | 28 | 2025/06/01 to 2025/06/28 | 11 | 8.63 | 1.47 | 3.50 | 0.299445 | 1.00000 | 1 day | 2-B08 | 2-Other viral infections characterized by skin and mucous membrane lesions, not elsewhere classified | 0 |
| 170 | 1-L03.21 | 1-Cellulitis and acute lymphangitis of face | 7 | 158 | 2025/06/20 to 2025/06/28 | 18 | 14.96 | 1.21 | 3.10 | 0.290665 | 1.00000 | 1 day | 1-L03.2,2-L03.21 | 1-Cellulitis and acute lymphangitis of face and neck,2-Cellulitis and acute lymphangitis of face | 0 |
| 171 | 2-S91.35 | 2-Open bite of foot | 6 | 19 | 2025/06/16 to 2025/06/28 | 4 | 2.73 | 1.61 | 1.52 | 0.259703 | 1.00000 | 1 day | 2-S91.3 | 2-Open wound of foot | 0 |
| 172 | 2-H60.2 | 2-Malignant otitis externa | 5 | 16 | 2025/06/03 to 2025/06/28 | 6 | 4.41 | 1.52 | 2.05 | 0.257857 | 1.00000 | 1 day | 2-H60 | 2-Otitis externa | 0 |
| 173 | 2-R41.84 | 2-Other specified cognitive deficit | 6 | 15 | 2025/06/08 to 2025/06/28 | 5 | 3.56 | 1.69 | 2.04 | 0.257174 | 1.00000 | 1 day | 2-R41.8 | 2-Other symptoms and signs involving cognitive functions and awareness | 0 |
| 174 | 2-S81.83 | 2-Puncture wound without foreign body of lower leg | 6 | 26 | 2025/06/17 to 2025/06/28 | 5 | 3.57 | 1.60 | 1.88 | 0.253506 | 1.00000 | 1 day | 2-S81.8 | 2-Open wound of lower leg | 0 |
| 175 | 2-R53.81 | 2-Other malaise | 6 | 600 | 2025/06/07 to 2025/06/28 | 152 | 143.41 | 1.08 | 11.71 | 0.252464 | 1.00000 | 1 day | 2-R53.8 | 2-Other malaise and fatigue | 0 |
| 176 | 2-R41.0 | 2-Disorientation, unspecified | 5 | 622 | 2025/06/19 to 2025/06/28 | 71 | 65.70 | 1.08 | 5.06 | 0.208494 | 1.00000 | 1 day | 2-R41 | 2-Other symptoms and signs involving cognitive functions and awareness | 0 |
| 177 | 2-R50.82 | 2-Postprocedural fever | 6 | 28 | 2025/06/20 to 2025/06/28 | 4 | 2.87 | 1.57 | 1.45 | 0.199267 | 1.00000 | 1 day | 2-R50.8 | 2-Other specified fever | 0 |
| 178 | 1-L03.213 | 1-Periorbital cellulitis | 8 | 72 | 2025/06/19 to 2025/06/28 | 9 | 7.29 | 1.19 | 1.46 | 0.185900 | 1.00000 | 1 day | 1-L03.21,2-L03.213 | 1-Cellulitis and acute lymphangitis of face,2-Periorbital cellulitis | 0 |
| 179 | 2-S91.05 | 2-Open bite of ankle | 6 | 25 | 2025/06/02 to 2025/06/28 | 9 | 7.31 | 1.34 | 2.27 | 0.182243 | 1.00000 | 1 day | 2-S91.0 | 2-Open wound of ankle | 0 |
| 180 | 1-L03.221 | 1-Cellulitis of neck | 8 | 19 | 2025/06/19 to 2025/06/28 | 3 | 2.11 | 1.57 | 1.09 | 0.165172 | 1.00000 | 1 day | 1-L03.22,2-L03.221 | 1-Cellulitis and acute lymphangitis of neck,2-Cellulitis of neck | 0 |
| 181 | 2-B97.1 | 2-Enterovirus as the cause of diseases classified elsewhere | 5 | 91 | 2025/06/18 to 2025/06/28 | 13 | 11.04 | 1.24 | 2.53 | 0.164242 | 1.00000 | 1 day | 2-B97,2-Other Enterovirus | 2-Viral agents as the cause of diseases classified elsewhere,2-Other Enterovirus | 0 |
| 182 | 2-B08.20 | 2-Exanthema subitum [sixth disease], unspecified | 6 | 27 | 2025/06/01 to 2025/06/28 | 10 | 8.33 | 1.33 | 2.50 | 0.158128 | 1.00000 | 1 day | 2-B08.2 | 2-Exanthema subitum [sixth disease] | 0 |
| 183 | 2-L03.316 | 2-Cellulitis of umbilicus | 7 | 20 | 2025/06/12 to 2025/06/28 | 5 | 3.87 | 1.49 | 1.64 | 0.151058 | 1.00000 | 1 day | 2-L03.31 | 2-Cellulitis of trunk | 0 |
| 184 | 1-E87.8 | 1-Other disorders of electrolyte and fluid balance, not elsewhere classified | 6 | 126 | 2025/06/01 to 2025/06/28 | 42 | 38.78 | 1.13 | 4.96 | 0.129990 | 1.00000 | 1 day | 1-E87,2-E87.8 | 1-Other disorders of fluid, electrolyte and acid-base balance,2-Other disorders of electrolyte and fluid balance, not elsewhere classified | 0 |
| 185 | 2-S91.10 | 2-Unspecified open wound of toe without damage to nail | 6 | 65 | 2025/06/25 to 2025/06/28 | 3 | 2.26 | 1.12 | 0.32 | 0.110250 | 1.00000 | 1 day | 2-S91.1 | 2-Open wound of toe without damage to nail | 0 |
| 186 | 1-B34.1 | 1-Enterovirus infection, unspecified | 6 | 36 | 2025/06/07 to 2025/06/28 | 10 | 8.61 | 1.23 | 1.86 | 0.106506 | 1.00000 | 1 day | 1-B34,1-Other Enterovirus,2-B34.1 | 1-Viral infection of unspecified site,1-Other Enterovirus,2-Enterovirus infection, unspecified | 0 |
| 187 | 1-S91.00 | 1-Unspecified open wound of ankle | 7 | 11 | 2025/06/12 to 2025/06/28 | 3 | 2.27 | 1.67 | 1.21 | 0.106030 | 1.00000 | 1 day | 1-S91.0,2-S91.00 | 1-Open wound of ankle,2-Unspecified open wound of ankle | 0 |
| 188 | 2-S91.34 | 2-Puncture wound with foreign body of foot | 6 | 9 | 2025/06/07 to 2025/06/28 | 3 | 2.31 | 1.60 | 1.12 | 0.094214 | 1.00000 | 1 day | 2-S91.3 | 2-Open wound of foot | 0 |
| 189 | 1-R53.81 | 1-Other malaise | 7 | 226 | 2025/06/04 to 2025/06/28 | 64 | 60.69 | 1.06 | 3.71 | 0.088547 | 1.00000 | 1 day | 1-R53.8,2-R53.81 | 1-Other malaise and fatigue,2-Other malaise | 0 |
| 190 | 1-R50.81 | 1-Fever presenting with conditions classified elsewhere | 7 | 76 | 2025/06/11 to 2025/06/28 | 16 | 14.39 | 1.10 | 1.49 | 0.086338 | 1.00000 | 1 day | 1-R50.8,2-R50.81 | 1-Other specified fever,2-Fever presenting with conditions classified elsewhere | 0 |
| 191 | 1-S91.1 | 1-Open wound of toe without damage to nail | 6 | 14 | 2025/06/08 to 2025/06/28 | 4 | 3.23 | 1.35 | 1.04 | 0.084266 | 1.00000 | 1 day | 1-S91,2-S91.1 | 1-Open wound of ankle, foot and toes,2-Open wound of toe without damage to nail | 0 |
| 192 | 2-L03.315 | 2-Cellulitis of perineum | 7 | 11 | 2025/06/11 to 2025/06/28 | 3 | 2.34 | 1.55 | 1.07 | 0.084202 | 1.00000 | 1 day | 2-L03.31 | 2-Cellulitis of trunk | 0 |
| 193 | 1-S91.3 | 1-Open wound of foot | 6 | 96 | 2025/06/04 to 2025/06/28 | 28 | 25.99 | 1.11 | 2.69 | 0.075843 | 1.00000 | 1 day | 1-S91,2-S91.3 | 1-Open wound of ankle, foot and toes,2-Open wound of foot | 0 |
| 194 | 2-L03.221 | 2-Cellulitis of neck | 7 | 72 | 2025/06/02 to 2025/06/28 | 23 | 21.22 | 1.12 | 2.39 | 0.072744 | 1.00000 | 1 day | 2-L03.22 | 2-Cellulitis and acute lymphangitis of neck | 0 |
| 195 | 2-E87.29 | 2-Other acidosis | 6 | 255 | 2025/06/24 to 2025/06/28 | 14 | 12.66 | 1.05 | 0.70 | 0.068216 | 1.00000 | 1 day | 2-E87.2 | 2-Acidosis | 0 |
| 196 | 1-S91.30 | 1-Unspecified open wound of foot | 7 | 84 | 2025/06/04 to 2025/06/28 | 24 | 22.76 | 1.07 | 1.67 | 0.033403 | 1.00000 | 1 day | 1-S91.3,2-S91.30 | 1-Open wound of foot,2-Unspecified open wound of foot | 0 |
| 197 | 1-E87.2 | 1-Acidosis | 6 | 727 | 2025/06/10 to 2025/06/28 | 152 | 149.13 | 1.02 | 2.62 | 0.027388 | 1.00000 | 1 day | 1-E87,2-E87.2 | 1-Other disorders of fluid, electrolyte and acid-base balance,2-Acidosis | 0 |
| 198 | 2-L03.21 | 2-Cellulitis and acute lymphangitis of face | 6 | 1146 | 2025/06/16 to 2025/06/28 | 165 | 162.19 | 1.02 | 2.64 | 0.024238 | 1.00000 | 1 day | 2-L03.2 | 2-Cellulitis and acute lymphangitis of face and neck | 0 |
| 199 | 1-E87.5 | 1-Hyperkalemia | 6 | 1022 | 2025/06/06 to 2025/06/28 | 255 | 251.78 | 1.00 | 0.37 | 0.020463 | 1.00000 | 1 day | 1-E87,2-E87.5 | 1-Other disorders of fluid, electrolyte and acid-base balance,2-Hyperkalemia | 0 |
| 200 | 2-L03.2 | 2-Cellulitis and acute lymphangitis of face and neck | 5 | 1218 | 2025/06/09 to 2025/06/28 | 269 | 265.85 | 1.01 | 3.22 | 0.018618 | 1.00000 | 1 day | 2-L03 | 2-Cellulitis and acute lymphangitis | 0 |
| 201 | 1-L03.11 | 1-Cellulitis of other parts of limb | 7 | 1030 | 2025/06/16 to 2025/06/28 | 149 | 146.90 | 1.02 | 3.19 | 0.014920 | 1.00000 | 1 day | 1-L03.1,2-L03.11 | 1-Cellulitis and acute lymphangitis of other parts of limb,2-Cellulitis of other parts of limb | 0 |
| 202 | 1-L03.01 | 1-Cellulitis of finger | 7 | 56 | 2025/06/10 to 2025/06/28 | 12 | 11.42 | 1.05 | 0.57 | 0.014331 | 1.00000 | 1 day | 1-L03.0,2-L03.01 | 1-Cellulitis and acute lymphangitis of finger and toe,2-Cellulitis of finger | 0 |
| 203 | 2-E87.79 | 2-Other fluid overload | 6 | 101 | 2025/06/22 to 2025/06/28 | 8 | 7.55 | 1.07 | 0.50 | 0.013424 | 1.00000 | 1 day | 2-E87.7 | 2-Fluid overload | 0 |
| 204 | 1-L03.1 | 1-Cellulitis and acute lymphangitis of other parts of limb | 6 | 1031 | 2025/06/16 to 2025/06/28 | 149 | 147.06 | 1.02 | 3.02 | 0.012803 | 1.00000 | 1 day | 1-L03,2-L03.1 | 1-Cellulitis and acute lymphangitis,2-Cellulitis and acute lymphangitis of other parts of limb | 0 |
| 205 | 1-S91.10 | 1-Unspecified open wound of toe without damage to nail | 7 | 12 | 2025/06/08 to 2025/06/28 | 3 | 2.78 | 1.13 | 0.33 | 0.008185 | 1.00000 | 1 day | 1-S91.1,2-S91.10 | 1-Open wound of toe without damage to nail,2-Unspecified open wound of toe without damage to nail | 0 |
| 206 | 1-S91.0 | 1-Open wound of ankle | 6 | 14 | 2025/06/12 to 2025/06/28 | 3 | 2.79 | 1.22 | 0.53 | 0.007475 | 1.00000 | 1 day | 1-S91,2-S91.0 | 1-Open wound of ankle, foot and toes,2-Open wound of ankle | 0 |
| 207 | 1-R50.8 | 1-Other specified fever | 6 | 91 | 2025/06/11 to 2025/06/28 | 18 | 17.50 | 1.02 | 0.34 | 0.007118 | 1.00000 | 1 day | 1-R50,2-R50.8 | 1-Fever of other and unknown origin,2-Other specified fever | 0 |
| 208 | 1-S91 | 1-Open wound of ankle, foot and toes | 5 | 127 | 2025/06/04 to 2025/06/28 | 35 | 34.32 | 1.02 | 0.76 | 0.006623 | 1.00000 | 1 day | 1-S90-S99,2-S91 | 1-Injuries to the ankle and foot,2-Open wound of ankle, foot and toes | 0 |
| 209 | 2-R53.0 | 2-Neoplastic (malignant) related fatigue | 5 | 10 | 2025/06/04 to 2025/06/28 | 3 | 2.83 | 1.15 | 0.39 | 0.004776 | 1.00000 | 1 day | 2-R53 | 2-Malaise and fatigue | 0 |
| 210 | 2-S91.30 | 2-Unspecified open wound of foot | 6 | 194 | 2025/06/04 to 2025/06/28 | 53 | 52.57 | 1.01 | 0.53 | 0.001784 | 1.00000 | 1 day | 2-S91.3 | 2-Open wound of foot | 0 |

Tree Visualization

#### Visualization of Analysis Tree ... loading

Color Nodes by P-Value

Color Nodes by Recurrence Interval

Color Nodes by Relative Risk

P-Value Legend

- > 0.05
- 0.05
- 0.01
- 0.001

Recurrence Interval Legend

- < 100 days
- 100 days
- 1 year
- 5 years
- 100 years

Relative Risk Legend

- < 2
- 2
- 4
- 8

1. Selecting the circle in the upper right corner of each node will expand/collapse children under node. The color of circle indicates best p-value / relative risk found in descendent nodes.
2. The visualization tree displays nodes with recurrence interval ≥ 100 days. Siblings and ancestry to root are also displayed for significant nodes, regardless of their recurrence interval.

A cut is statistically significant when its log likelihood ratio is greater than the critical value, which is, for significance level:

... 0.00001: 14.24847

... 0.0001: 11.85986

... 0.001: 10.05381

... 0.01: 8.09809

... 0.05: 6.75095

\_\_\_\_\_\_\_\_\_\_\_\_\_\_\_\_\_\_\_\_\_\_\_\_\_\_\_\_\_\_\_\_\_\_\_\_\_\_\_\_\_\_\_\_\_\_\_\_\_\_\_\_\_\_\_\_\_\_\_\_\_\_\_\_\_\_\_\_\_\_\_\_\_\_\_\_\_\_\_\_

##### Additional Results Files

|  |  |
| --- | --- |
| Results HTML : | \\nasprgshare220\Share\DIS\BCD\COMDISshared\Analyst\_of\_the\_week\Asyndromic\Manuscript\Draft\Appendix\june 29 analysis\Results\_20250629.html |
| Temporal Graph File : | \\nasprgshare220\Share\DIS\BCD\COMDISshared\Analyst\_of\_the\_week\Asyndromic\Manuscript\Draft\Appendix\june 29 analysis\Results\_20250629.temporal.html |

##### Parameter Settings

##### Input

|  |  |
| --- | --- |
| Tree File : | \\nasprgshare220\Share\DIS\BCD\COMDISshared\Analyst\_of\_the\_week\Asyndromic\Manuscript\Draft\Appendix\june 29 analysis\Tree\_File\_20250629.csv |
| Count File : | \\nasprgshare220\Share\DIS\BCD\COMDISshared\Analyst\_of\_the\_week\Asyndromic\Manuscript\Draft\Appendix\june 29 analysis\Count\_File\_20250629.txt |
| Data Time Range : | [2025/03/31,2025/06/28] |
| Time Precision : | Day |

##### Analysis

|  |  |
| --- | --- |
| Type of Scan : | Tree and Time |
| Conditional Analysis : | Node and Time |
| Scan Rate : | High Rates |

##### Output

|  |  |
| --- | --- |
| Results File : | \\nasprgshare220\Share\DIS\BCD\COMDISshared\Analyst\_of\_the\_week\Asyndromic\Manuscript\Draft\Appendix\june 29 analysis\Results\_20250629.txt |
| Report Results as HTML : | Yes |
| Report Results as CSV Table : | No |
| Generate NCBI Genome Workbench ASN1 File : | No |
| Generate Newick Tree Format File : | No |

##### Advanced Input

|  |  |
| --- | --- |
| Cut File : |  |
| Only Allow Data on Leaves of Tree : | No |
| Relaxed Study Data Period Checking : | Yes |
| Allow Multiple Parents for the Same Node : | Yes |
| Allow Multiple Root Nodes : | No |

##### Temporal Window

|  |  |
| --- | --- |
| Maximum Temporal Window : | 28 Time Units |
| Minimum Temporal Window : | 1 Time Units |
| Apply Risk Window Restriction : | No |
| Prospective Analysis : | Yes |

##### Adjustments

|  |  |
| --- | --- |
| Perform Day of Week Adjustment : | Yes |
| Apply Data Time Range Exclusions : | No |

##### Inference

|  |  |
| --- | --- |
| P-Value Reporting : | Sequential Monte Carlo Early Termination |
| Termination Cutoff : | 5000 |
| Number of Replications : | 99999 |
| Restrict Tree Levels : | Yes |
| Tree Levels Excluded From Evaluation : | 1,2,3 |
| Restrict Evaluated Tree Nodes : | Yes |
| Not Evaluated Nodes File : | \\nasprgshare220\Share\DIS\BCD\COMDISshared\Analyst\_of\_the\_week\Asyndromic\Manuscript\Draft\Appendix\june 29 analysis\Do\_not\_evaluate\_nodes.csv |
| Minimum Number of High Rate Node Cases : | 3 |

##### Miscellaneous

|  |  |
| --- | --- |
| Prospective Analysis Frequency : | Daily |

##### Additional Output

|  |  |
| --- | --- |
| Attributable Risk : | No |
| Report Simulated Log Likelihood Ratios : | No |
| Report Critical Values : | Yes |
| Temporal Graph File : | Yes |

##### Run Options

|  |  |
| --- | --- |
| Processer Usage : | All Available Processors |

##### COMPUTATIONAL INFORMATION

|  |  |
| --- | --- |
| Program run on | Tue Oct 7 15:25:13 2025 |
| Program completed | Tue Oct 7 17:54:43 2025 |
| Total Running Time | 2 hours 29 minutes 30 seconds |
| Processor Usage | 8 processors |
| Version | TreeScan v2.3 |
