## Supplementary material for "Asyndromic Surveillance of New York City Emergency Department Diagnoses with the Tree-Temporal Scan Statistic": Reproducing a TreeScan analysis: Results_20250629.temporal.html

 


Cluster Temporal Graph


Search

Choose which graphs to display:

2-B08.4 (2-Enteroviral vesicular stomatitis with e..
2-B08 (2-Other viral infections characterized by s..
2-Other Enterovirus
2-T67 (2-Effects of heat and light)
2-T67.5 (2-Heat exhaustion, unspecified)
2-T67.9 (2-Effect of heat and light, unspecified)
2-T67.01 (2-Heatstroke and sunstroke)
2-T67.0 (2-Heatstroke and sunstroke)
1-T67 (1-Effects of heat and light)
2-W57 (2-Bitten or stung by nonvenomous insect and..
2-N17 (2-Acute kidney failure)
2-E86 (2-Volume depletion)
2-N17.9 (2-Acute kidney failure, unspecified)
1-T67.01 (1-Heatstroke and sunstroke)
2-E86.0 (2-Dehydration)
2-R50 (2-Fever of other and unknown origin)
2-R50.9 (2-Fever, unspecified)
2-L74.0 (2-Miliaria rubra)
2-H60 (2-Otitis externa)
2-T67.1 (2-Heat syncope)
2-L74 (2-Eccrine sweat disorders)
2-M79.89 (2-Other specified soft tissue disorders)
2-B34.1 (2-Enterovirus infection, unspecified)
2-B08.5 (2-Enteroviral vesicular pharyngitis)
2-M79.8 (2-Other specified soft tissue disorders)
2-R55 (2-Syncope and collapse)
2-R41.8 (2-Other symptoms and signs involving cogn..
2-R41.82 (2-Altered mental status, unspecified)
1-T67.5 (1-Heat exhaustion, unspecified)
2-R53 (2-Malaise and fatigue)
2-R41 (2-Other symptoms and signs involving cognit..
2-E87.1 (2-Hypo-osmolality and hyponatremia)
1-N17 (1-Acute kidney failure)
2-L55 (2-Sunburn)
2-S91 (2-Open wound of ankle, foot and toes)
2-R53.1 (2-Weakness)
2-S81.81 (2-Laceration without foreign body of low..
1-N17.9 (1-Acute kidney failure, unspecified)
2-S81.8 (2-Open wound of lower leg)
2-L03 (2-Cellulitis and acute lymphangitis)
2-L55.9 (2-Sunburn, unspecified)
2-M62.82 (2-Rhabdomyolysis)
2-H60.50 (2-Unspecified acute noninfective otitis ..
2-E87 (2-Other disorders of fluid, electrolyte and..
1-R41.8 (1-Other symptoms and signs involving cogn..
1-R41.82 (1-Altered mental status, unspecified)
2-H60.9 (2-Unspecified otitis externa)
2-L23.7 (2-Allergic contact dermatitis due to plan..
2-H60.5 (2-Acute noninfective otitis externa)
2-H60.3 (2-Other infective otitis externa)
1-R50.9 (1-Fever, unspecified)
1-R50 (1-Fever of other and unknown origin)
1-R41 (1-Other symptoms and signs involving cognit..
2-T67.8 (2-Other effects of heat and light)
2-S91.31 (2-Laceration without foreign body of foo..
2-L03.90 (2-Cellulitis, unspecified)
1-E87.1 (1-Hypo-osmolality and hyponatremia)
2-L03.11 (2-Cellulitis of other parts of limb)
2-L03.1 (2-Cellulitis and acute lymphangitis of ot..
2-S91.3 (2-Open wound of foot)
2-H60.39 (2-Other infective otitis externa)
2-S91.2 (2-Open wound of toe with damage to nail)
2-S91.20 (2-Unspecified open wound of toe with dam..
2-H60.33 (2-Swimmer's ear)
2-S91.33 (2-Puncture wound without foreign body of..
1-M62.82 (1-Rhabdomyolysis)
2-T67.2 (2-Heat cramp)
2-S81.80 (2-Unspecified open wound of lower leg)
1-B08.4 (1-Enteroviral vesicular stomatitis with e..
1-W57 (1-Bitten or stung by nonvenomous insect and..
2-R53.83 (2-Other fatigue)
2-S91.01 (2-Laceration without foreign body of ank..
2-S91.0 (2-Open wound of ankle)
1-R55 (1-Syncope and collapse)
2-E86.1 (2-Hypovolemia)
2-B08.1 (2-Molluscum contagiosum)
1-R53.1 (1-Weakness)
1-M79.89 (1-Other specified soft tissue disorders)
2-H60.31 (2-Diffuse otitis externa)
2-S91.11 (2-Laceration without foreign body of toe..
1-B08 (1-Other viral infections characterized by s..
1-M79.8 (1-Other specified soft tissue disorders)
2-E87.6 (2-Hypokalemia)
1-E86 (1-Volume depletion)
2-E87.70 (2-Fluid overload, unspecified)
1-T67.1 (1-Heat syncope)
2-L03.0 (2-Cellulitis and acute lymphangitis of fi..
2-E87.7 (2-Fluid overload)
1-R53 (1-Malaise and fatigue)
1-E86.1 (1-Hypovolemia)
2-L03.317 (2-Cellulitis of buttock)
2-L03.01 (2-Cellulitis of finger)
2-L55.0 (2-Sunburn of first degree)
2-S91.1 (2-Open wound of toe without damage to nai..
1-E87 (1-Other disorders of fluid, electrolyte and..
1-L03.90 (1-Cellulitis, unspecified)
2-E86.9 (2-Volume depletion, unspecified)
1-L03.311 (1-Cellulitis of abdominal wall)
2-R53.8 (2-Other malaise and fatigue)
1-H60.9 (1-Unspecified otitis externa)

Apply
Cancel

0 % Complete

Show Chart Options

#### Chart Options

Title

Title can be changed by editing this text.

Series Inside Cluster

Observed

Expected

Observed / Expected

Percent Cases

Series Outside Cluster

Observed

Expected

Observed Chart Type

Histogram

Line

Switch the series type between line and histogram.

Cluster Band

Show Cluster Band

Band stretching across the plot area marking cluster interval.

To zoom a portion of the chart, select and drag mouse within the chart. Hold down shift key to pan zoomed chart.

#### 2 x 2 Table

|  | Cluster Time Period | |
| --- | --- | --- |
| Cluster Node | Inside | Outside |
| Inside | 1070 | 802 |
| Outside | 214882 | 830456 |
|  | 0.5% | 0.1% |

Close Chart Options

Show Chart Options

#### Chart Options

Title

Title can be changed by editing this text.

Series Inside Cluster

Observed

Expected

Observed / Expected

Percent Cases

Series Outside Cluster

Observed

Expected

Observed Chart Type

Histogram

Line

Switch the series type between line and histogram.

Cluster Band

Show Cluster Band

Band stretching across the plot area marking cluster interval.

To zoom a portion of the chart, select and drag mouse within the chart. Hold down shift key to pan zoomed chart.

#### 2 x 2 Table

|  | Cluster Time Period | |
| --- | --- | --- |
| Cluster Node | Inside | Outside |
| Inside | 1217 | 1085 |
| Outside | 214735 | 830173 |
|  | 0.6% | 0.1% |

Close Chart Options

Show Chart Options

#### Chart Options

Title

Title can be changed by editing this text.

Series Inside Cluster

Observed

Expected

Observed / Expected

Percent Cases

Series Outside Cluster

Observed

Expected

Observed Chart Type

Histogram

Line

Switch the series type between line and histogram.

Cluster Band

Show Cluster Band

Band stretching across the plot area marking cluster interval.

To zoom a portion of the chart, select and drag mouse within the chart. Hold down shift key to pan zoomed chart.

#### 2 x 2 Table

|  | Cluster Time Period | |
| --- | --- | --- |
| Cluster Node | Inside | Outside |
| Inside | 1423 | 1542 |
| Outside | 214529 | 829716 |
|  | 0.7% | 0.2% |

Close Chart Options

Show Chart Options

#### Chart Options

Title

Title can be changed by editing this text.

Series Inside Cluster

Observed

Expected

Observed / Expected

Percent Cases

Series Outside Cluster

Observed

Expected

Observed Chart Type

Histogram

Line

Switch the series type between line and histogram.

Cluster Band

Show Cluster Band

Band stretching across the plot area marking cluster interval.

To zoom a portion of the chart, select and drag mouse within the chart. Hold down shift key to pan zoomed chart.

#### 2 x 2 Table

|  | Cluster Time Period | |
| --- | --- | --- |
| Cluster Node | Inside | Outside |
| Inside | 282 | 143 |
| Outside | 77909 | 968876 |
|  | 0.4% | 0.0% |

Close Chart Options

Show Chart Options

#### Chart Options

Title

Title can be changed by editing this text.

Series Inside Cluster

Observed

Expected

Observed / Expected

Percent Cases

Series Outside Cluster

Observed

Expected

Observed Chart Type

Histogram

Line

Switch the series type between line and histogram.

Cluster Band

Show Cluster Band

Band stretching across the plot area marking cluster interval.

To zoom a portion of the chart, select and drag mouse within the chart. Hold down shift key to pan zoomed chart.

#### 2 x 2 Table

|  | Cluster Time Period | |
| --- | --- | --- |
| Cluster Node | Inside | Outside |
| Inside | 155 | 40 |
| Outside | 78036 | 968979 |
|  | 0.2% | 0.0% |

Close Chart Options

Show Chart Options

#### Chart Options

Title

Title can be changed by editing this text.

Series Inside Cluster

Observed

Expected

Observed / Expected

Percent Cases

Series Outside Cluster

Observed

Expected

Observed Chart Type

Histogram

Line

Switch the series type between line and histogram.

Cluster Band

Show Cluster Band

Band stretching across the plot area marking cluster interval.

To zoom a portion of the chart, select and drag mouse within the chart. Hold down shift key to pan zoomed chart.

#### 2 x 2 Table

|  | Cluster Time Period | |
| --- | --- | --- |
| Cluster Node | Inside | Outside |
| Inside | 47 | 7 |
| Outside | 88968 | 958188 |
|  | 0.1% | 0.0% |

Close Chart Options

Show Chart Options

#### Chart Options

Title

Title can be changed by editing this text.

Series Inside Cluster

Observed

Expected

Observed / Expected

Percent Cases

Series Outside Cluster

Observed

Expected

Observed Chart Type

Histogram

Line

Switch the series type between line and histogram.

Cluster Band

Show Cluster Band

Band stretching across the plot area marking cluster interval.

To zoom a portion of the chart, select and drag mouse within the chart. Hold down shift key to pan zoomed chart.

#### 2 x 2 Table

|  | Cluster Time Period | |
| --- | --- | --- |
| Cluster Node | Inside | Outside |
| Inside | 39 | 14 |
| Outside | 67597 | 979560 |
|  | 0.1% | 0.0% |

Close Chart Options

Show Chart Options

#### Chart Options

Title

Title can be changed by editing this text.

Series Inside Cluster

Observed

Expected

Observed / Expected

Percent Cases

Series Outside Cluster

Observed

Expected

Observed Chart Type

Histogram

Line

Switch the series type between line and histogram.

Cluster Band

Show Cluster Band

Band stretching across the plot area marking cluster interval.

To zoom a portion of the chart, select and drag mouse within the chart. Hold down shift key to pan zoomed chart.

#### 2 x 2 Table

|  | Cluster Time Period | |
| --- | --- | --- |
| Cluster Node | Inside | Outside |
| Inside | 39 | 17 |
| Outside | 67597 | 979557 |
|  | 0.1% | 0.0% |

Close Chart Options

Show Chart Options

#### Chart Options

Title

Title can be changed by editing this text.

Series Inside Cluster

Observed

Expected

Observed / Expected

Percent Cases

Series Outside Cluster

Observed

Expected

Observed Chart Type

Histogram

Line

Switch the series type between line and histogram.

Cluster Band

Show Cluster Band

Band stretching across the plot area marking cluster interval.

To zoom a portion of the chart, select and drag mouse within the chart. Hold down shift key to pan zoomed chart.

#### 2 x 2 Table

|  | Cluster Time Period | |
| --- | --- | --- |
| Cluster Node | Inside | Outside |
| Inside | 44 | 37 |
| Outside | 78147 | 968982 |
|  | 0.1% | 0.0% |

Close Chart Options

Show Chart Options

#### Chart Options

Title

Title can be changed by editing this text.

Series Inside Cluster

Observed

Expected

Observed / Expected

Percent Cases

Series Outside Cluster

Observed

Expected

Observed Chart Type

Histogram

Line

Switch the series type between line and histogram.

Cluster Band

Show Cluster Band

Band stretching across the plot area marking cluster interval.

To zoom a portion of the chart, select and drag mouse within the chart. Hold down shift key to pan zoomed chart.

#### 2 x 2 Table

|  | Cluster Time Period | |
| --- | --- | --- |
| Cluster Node | Inside | Outside |
| Inside | 474 | 807 |
| Outside | 249258 | 796671 |
|  | 0.2% | 0.1% |

Close Chart Options

Show Chart Options

#### Chart Options

Title

Title can be changed by editing this text.

Series Inside Cluster

Observed

Expected

Observed / Expected

Percent Cases

Series Outside Cluster

Observed

Expected

Observed Chart Type

Histogram

Line

Switch the series type between line and histogram.

Cluster Band

Show Cluster Band

Band stretching across the plot area marking cluster interval.

To zoom a portion of the chart, select and drag mouse within the chart. Hold down shift key to pan zoomed chart.

#### 2 x 2 Table

|  | Cluster Time Period | |
| --- | --- | --- |
| Cluster Node | Inside | Outside |
| Inside | 519 | 4765 |
| Outside | 67117 | 974809 |
|  | 0.8% | 0.5% |

Close Chart Options

Show Chart Options

#### Chart Options

Title

Title can be changed by editing this text.

Series Inside Cluster

Observed

Expected

Observed / Expected

Percent Cases

Series Outside Cluster

Observed

Expected

Observed Chart Type

Histogram

Line

Switch the series type between line and histogram.

Cluster Band

Show Cluster Band

Band stretching across the plot area marking cluster interval.

To zoom a portion of the chart, select and drag mouse within the chart. Hold down shift key to pan zoomed chart.

#### 2 x 2 Table

|  | Cluster Time Period | |
| --- | --- | --- |
| Cluster Node | Inside | Outside |
| Inside | 281 | 2171 |
| Outside | 67355 | 977403 |
|  | 0.4% | 0.2% |

Close Chart Options

Show Chart Options

#### Chart Options

Title

Title can be changed by editing this text.

Series Inside Cluster

Observed

Expected

Observed / Expected

Percent Cases

Series Outside Cluster

Observed

Expected

Observed Chart Type

Histogram

Line

Switch the series type between line and histogram.

Cluster Band

Show Cluster Band

Band stretching across the plot area marking cluster interval.

To zoom a portion of the chart, select and drag mouse within the chart. Hold down shift key to pan zoomed chart.

#### 2 x 2 Table

|  | Cluster Time Period | |
| --- | --- | --- |
| Cluster Node | Inside | Outside |
| Inside | 512 | 4706 |
| Outside | 67124 | 974868 |
|  | 0.8% | 0.5% |

Close Chart Options

Show Chart Options

#### Chart Options

Title

Title can be changed by editing this text.

Series Inside Cluster

Observed

Expected

Observed / Expected

Percent Cases

Series Outside Cluster

Observed

Expected

Observed Chart Type

Histogram

Line

Switch the series type between line and histogram.

Cluster Band

Show Cluster Band

Band stretching across the plot area marking cluster interval.

To zoom a portion of the chart, select and drag mouse within the chart. Hold down shift key to pan zoomed chart.

#### 2 x 2 Table

|  | Cluster Time Period | |
| --- | --- | --- |
| Cluster Node | Inside | Outside |
| Inside | 25 | 10 |
| Outside | 67611 | 979564 |
|  | 0.0% | 0.0% |

Close Chart Options

Show Chart Options

#### Chart Options

Title

Title can be changed by editing this text.

Series Inside Cluster

Observed

Expected

Observed / Expected

Percent Cases

Series Outside Cluster

Observed

Expected

Observed Chart Type

Histogram

Line

Switch the series type between line and histogram.

Cluster Band

Show Cluster Band

Band stretching across the plot area marking cluster interval.

To zoom a portion of the chart, select and drag mouse within the chart. Hold down shift key to pan zoomed chart.

#### 2 x 2 Table

|  | Cluster Time Period | |
| --- | --- | --- |
| Cluster Node | Inside | Outside |
| Inside | 251 | 2013 |
| Outside | 67385 | 977561 |
|  | 0.4% | 0.2% |

Close Chart Options

Show Chart Options

#### Chart Options

Title

Title can be changed by editing this text.

Series Inside Cluster

Observed

Expected

Observed / Expected

Percent Cases

Series Outside Cluster

Observed

Expected

Observed Chart Type

Histogram

Line

Switch the series type between line and histogram.

Cluster Band

Show Cluster Band

Band stretching across the plot area marking cluster interval.

To zoom a portion of the chart, select and drag mouse within the chart. Hold down shift key to pan zoomed chart.

#### 2 x 2 Table

|  | Cluster Time Period | |
| --- | --- | --- |
| Cluster Node | Inside | Outside |
| Inside | 1366 | 7920 |
| Outside | 122579 | 915345 |
|  | 1.1% | 0.9% |

Close Chart Options

Show Chart Options

#### Chart Options

Title

Title can be changed by editing this text.

Series Inside Cluster

Observed

Expected

Observed / Expected

Percent Cases

Series Outside Cluster

Observed

Expected

Observed Chart Type

Histogram

Line

Switch the series type between line and histogram.

Cluster Band

Show Cluster Band

Band stretching across the plot area marking cluster interval.

To zoom a portion of the chart, select and drag mouse within the chart. Hold down shift key to pan zoomed chart.

#### 2 x 2 Table

|  | Cluster Time Period | |
| --- | --- | --- |
| Cluster Node | Inside | Outside |
| Inside | 1331 | 7685 |
| Outside | 122614 | 915580 |
|  | 1.1% | 0.8% |

Close Chart Options

Show Chart Options

#### Chart Options

Title

Title can be changed by editing this text.

Series Inside Cluster

Observed

Expected

Observed / Expected

Percent Cases

Series Outside Cluster

Observed

Expected

Observed Chart Type

Histogram

Line

Switch the series type between line and histogram.

Cluster Band

Show Cluster Band

Band stretching across the plot area marking cluster interval.

To zoom a portion of the chart, select and drag mouse within the chart. Hold down shift key to pan zoomed chart.

#### 2 x 2 Table

|  | Cluster Time Period | |
| --- | --- | --- |
| Cluster Node | Inside | Outside |
| Inside | 46 | 66 |
| Outside | 100668 | 946430 |
|  | 0.0% | 0.0% |

Close Chart Options

Show Chart Options

#### Chart Options

Title

Title can be changed by editing this text.

Series Inside Cluster

Observed

Expected

Observed / Expected

Percent Cases

Series Outside Cluster

Observed

Expected

Observed Chart Type

Histogram

Line

Switch the series type between line and histogram.

Cluster Band

Show Cluster Band

Band stretching across the plot area marking cluster interval.

To zoom a portion of the chart, select and drag mouse within the chart. Hold down shift key to pan zoomed chart.

#### 2 x 2 Table

|  | Cluster Time Period | |
| --- | --- | --- |
| Cluster Node | Inside | Outside |
| Inside | 579 | 1567 |
| Outside | 203379 | 841685 |
|  | 0.3% | 0.2% |

Close Chart Options

Show Chart Options

#### Chart Options

Title

Title can be changed by editing this text.

Series Inside Cluster

Observed

Expected

Observed / Expected

Percent Cases

Series Outside Cluster

Observed

Expected

Observed Chart Type

Histogram

Line

Switch the series type between line and histogram.

Cluster Band

Show Cluster Band

Band stretching across the plot area marking cluster interval.

To zoom a portion of the chart, select and drag mouse within the chart. Hold down shift key to pan zoomed chart.

#### 2 x 2 Table

|  | Cluster Time Period | |
| --- | --- | --- |
| Cluster Node | Inside | Outside |
| Inside | 40 | 70 |
| Outside | 88975 | 958125 |
|  | 0.0% | 0.0% |

Close Chart Options

Show Chart Options

#### Chart Options

Title

Title can be changed by editing this text.

Series Inside Cluster

Observed

Expected

Observed / Expected

Percent Cases

Series Outside Cluster

Observed

Expected

Observed Chart Type

Histogram

Line

Switch the series type between line and histogram.

Cluster Band

Show Cluster Band

Band stretching across the plot area marking cluster interval.

To zoom a portion of the chart, select and drag mouse within the chart. Hold down shift key to pan zoomed chart.

#### 2 x 2 Table

|  | Cluster Time Period | |
| --- | --- | --- |
| Cluster Node | Inside | Outside |
| Inside | 49 | 90 |
| Outside | 100665 | 946406 |
|  | 0.0% | 0.0% |

Close Chart Options

Show Chart Options

#### Chart Options

Title

Title can be changed by editing this text.

Series Inside Cluster

Observed

Expected

Observed / Expected

Percent Cases

Series Outside Cluster

Observed

Expected

Observed Chart Type

Histogram

Line

Switch the series type between line and histogram.

Cluster Band

Show Cluster Band

Band stretching across the plot area marking cluster interval.

To zoom a portion of the chart, select and drag mouse within the chart. Hold down shift key to pan zoomed chart.

#### 2 x 2 Table

|  | Cluster Time Period | |
| --- | --- | --- |
| Cluster Node | Inside | Outside |
| Inside | 1224 | 2836 |
| Outside | 259780 | 783370 |
|  | 0.5% | 0.4% |

Close Chart Options

Show Chart Options

#### Chart Options

Title

Title can be changed by editing this text.

Series Inside Cluster

Observed

Expected

Observed / Expected

Percent Cases

Series Outside Cluster

Observed

Expected

Observed Chart Type

Histogram

Line

Switch the series type between line and histogram.

Cluster Band

Show Cluster Band

Band stretching across the plot area marking cluster interval.

To zoom a portion of the chart, select and drag mouse within the chart. Hold down shift key to pan zoomed chart.

#### 2 x 2 Table

|  | Cluster Time Period | |
| --- | --- | --- |
| Cluster Node | Inside | Outside |
| Inside | 259 | 442 |
| Outside | 249473 | 797036 |
|  | 0.1% | 0.1% |

Close Chart Options

Show Chart Options

#### Chart Options

Title

Title can be changed by editing this text.

Series Inside Cluster

Observed

Expected

Observed / Expected

Percent Cases

Series Outside Cluster

Observed

Expected

Observed Chart Type

Histogram

Line

Switch the series type between line and histogram.

Cluster Band

Show Cluster Band

Band stretching across the plot area marking cluster interval.

To zoom a portion of the chart, select and drag mouse within the chart. Hold down shift key to pan zoomed chart.

#### 2 x 2 Table

|  | Cluster Time Period | |
| --- | --- | --- |
| Cluster Node | Inside | Outside |
| Inside | 85 | 214 |
| Outside | 135524 | 911387 |
|  | 0.1% | 0.0% |

Close Chart Options

Show Chart Options

#### Chart Options

Title

Title can be changed by editing this text.

Series Inside Cluster

Observed

Expected

Observed / Expected

Percent Cases

Series Outside Cluster

Observed

Expected

Observed Chart Type

Histogram

Line

Switch the series type between line and histogram.

Cluster Band

Show Cluster Band

Band stretching across the plot area marking cluster interval.

To zoom a portion of the chart, select and drag mouse within the chart. Hold down shift key to pan zoomed chart.

#### 2 x 2 Table

|  | Cluster Time Period | |
| --- | --- | --- |
| Cluster Node | Inside | Outside |
| Inside | 1228 | 2870 |
| Outside | 259776 | 783336 |
|  | 0.5% | 0.4% |

Close Chart Options

Show Chart Options

#### Chart Options

Title

Title can be changed by editing this text.

Series Inside Cluster

Observed

Expected

Observed / Expected

Percent Cases

Series Outside Cluster

Observed

Expected

Observed Chart Type

Histogram

Line

Switch the series type between line and histogram.

Cluster Band

Show Cluster Band

Band stretching across the plot area marking cluster interval.

To zoom a portion of the chart, select and drag mouse within the chart. Hold down shift key to pan zoomed chart.

#### 2 x 2 Table

|  | Cluster Time Period | |
| --- | --- | --- |
| Cluster Node | Inside | Outside |
| Inside | 1513 | 10485 |
| Outside | 110416 | 924796 |
|  | 1.4% | 1.1% |

Close Chart Options

Show Chart Options

#### Chart Options

Title

Title can be changed by editing this text.

Series Inside Cluster

Observed

Expected

Observed / Expected

Percent Cases

Series Outside Cluster

Observed

Expected

Observed Chart Type

Histogram

Line

Switch the series type between line and histogram.

Cluster Band

Show Cluster Band

Band stretching across the plot area marking cluster interval.

To zoom a portion of the chart, select and drag mouse within the chart. Hold down shift key to pan zoomed chart.

#### 2 x 2 Table

|  | Cluster Time Period | |
| --- | --- | --- |
| Cluster Node | Inside | Outside |
| Inside | 433 | 4595 |
| Outside | 67203 | 974979 |
|  | 0.6% | 0.5% |

Close Chart Options

Show Chart Options

#### Chart Options

Title

Title can be changed by editing this text.

Series Inside Cluster

Observed

Expected

Observed / Expected

Percent Cases

Series Outside Cluster

Observed

Expected

Observed Chart Type

Histogram

Line

Switch the series type between line and histogram.

Cluster Band

Show Cluster Band

Band stretching across the plot area marking cluster interval.

To zoom a portion of the chart, select and drag mouse within the chart. Hold down shift key to pan zoomed chart.

#### 2 x 2 Table

|  | Cluster Time Period | |
| --- | --- | --- |
| Cluster Node | Inside | Outside |
| Inside | 409 | 4323 |
| Outside | 67227 | 975251 |
|  | 0.6% | 0.4% |

Close Chart Options

Show Chart Options

#### Chart Options

Title

Title can be changed by editing this text.

Series Inside Cluster

Observed

Expected

Observed / Expected

Percent Cases

Series Outside Cluster

Observed

Expected

Observed Chart Type

Histogram

Line

Switch the series type between line and histogram.

Cluster Band

Show Cluster Band

Band stretching across the plot area marking cluster interval.

To zoom a portion of the chart, select and drag mouse within the chart. Hold down shift key to pan zoomed chart.

#### 2 x 2 Table

|  | Cluster Time Period | |
| --- | --- | --- |
| Cluster Node | Inside | Outside |
| Inside | 11 | 4 |
| Outside | 54747 | 992448 |
|  | 0.0% | 0.0% |

Close Chart Options

Show Chart Options

#### Chart Options

Title

Title can be changed by editing this text.

Series Inside Cluster

Observed

Expected

Observed / Expected

Percent Cases

Series Outside Cluster

Observed

Expected

Observed Chart Type

Histogram

Line

Switch the series type between line and histogram.

Cluster Band

Show Cluster Band

Band stretching across the plot area marking cluster interval.

To zoom a portion of the chart, select and drag mouse within the chart. Hold down shift key to pan zoomed chart.

#### 2 x 2 Table

|  | Cluster Time Period | |
| --- | --- | --- |
| Cluster Node | Inside | Outside |
| Inside | 616 | 6796 |
| Outside | 67020 | 972778 |
|  | 0.9% | 0.7% |

Close Chart Options

Show Chart Options

#### Chart Options

Title

Title can be changed by editing this text.

Series Inside Cluster

Observed

Expected

Observed / Expected

Percent Cases

Series Outside Cluster

Observed

Expected

Observed Chart Type

Histogram

Line

Switch the series type between line and histogram.

Cluster Band

Show Cluster Band

Band stretching across the plot area marking cluster interval.

To zoom a portion of the chart, select and drag mouse within the chart. Hold down shift key to pan zoomed chart.

#### 2 x 2 Table

|  | Cluster Time Period | |
| --- | --- | --- |
| Cluster Node | Inside | Outside |
| Inside | 630 | 5252 |
| Outside | 88385 | 952943 |
|  | 0.7% | 0.5% |

Close Chart Options

Show Chart Options

#### Chart Options

Title

Title can be changed by editing this text.

Series Inside Cluster

Observed

Expected

Observed / Expected

Percent Cases

Series Outside Cluster

Observed

Expected

Observed Chart Type

Histogram

Line

Switch the series type between line and histogram.

Cluster Band

Show Cluster Band

Band stretching across the plot area marking cluster interval.

To zoom a portion of the chart, select and drag mouse within the chart. Hold down shift key to pan zoomed chart.

#### 2 x 2 Table

|  | Cluster Time Period | |
| --- | --- | --- |
| Cluster Node | Inside | Outside |
| Inside | 239 | 1690 |
| Outside | 88776 | 956505 |
|  | 0.3% | 0.2% |

Close Chart Options

Show Chart Options

#### Chart Options

Title

Title can be changed by editing this text.

Series Inside Cluster

Observed

Expected

Observed / Expected

Percent Cases

Series Outside Cluster

Observed

Expected

Observed Chart Type

Histogram

Line

Switch the series type between line and histogram.

Cluster Band

Show Cluster Band

Band stretching across the plot area marking cluster interval.

To zoom a portion of the chart, select and drag mouse within the chart. Hold down shift key to pan zoomed chart.

#### 2 x 2 Table

|  | Cluster Time Period | |
| --- | --- | --- |
| Cluster Node | Inside | Outside |
| Inside | 322 | 3256 |
| Outside | 67314 | 976318 |
|  | 0.5% | 0.3% |

Close Chart Options

Show Chart Options

#### Chart Options

Title

Title can be changed by editing this text.

Series Inside Cluster

Observed

Expected

Observed / Expected

Percent Cases

Series Outside Cluster

Observed

Expected

Observed Chart Type

Histogram

Line

Switch the series type between line and histogram.

Cluster Band

Show Cluster Band

Band stretching across the plot area marking cluster interval.

To zoom a portion of the chart, select and drag mouse within the chart. Hold down shift key to pan zoomed chart.

#### 2 x 2 Table

|  | Cluster Time Period | |
| --- | --- | --- |
| Cluster Node | Inside | Outside |
| Inside | 31 | 75 |
| Outside | 100683 | 946421 |
|  | 0.0% | 0.0% |

Close Chart Options

Show Chart Options

#### Chart Options

Title

Title can be changed by editing this text.

Series Inside Cluster

Observed

Expected

Observed / Expected

Percent Cases

Series Outside Cluster

Observed

Expected

Observed Chart Type

Histogram

Line

Switch the series type between line and histogram.

Cluster Band

Show Cluster Band

Band stretching across the plot area marking cluster interval.

To zoom a portion of the chart, select and drag mouse within the chart. Hold down shift key to pan zoomed chart.

#### 2 x 2 Table

|  | Cluster Time Period | |
| --- | --- | --- |
| Cluster Node | Inside | Outside |
| Inside | 223 | 1046 |
| Outside | 123722 | 922219 |
|  | 0.2% | 0.1% |

Close Chart Options

Show Chart Options

#### Chart Options

Title

Title can be changed by editing this text.

Series Inside Cluster

Observed

Expected

Observed / Expected

Percent Cases

Series Outside Cluster

Observed

Expected

Observed Chart Type

Histogram

Line

Switch the series type between line and histogram.

Cluster Band

Show Cluster Band

Band stretching across the plot area marking cluster interval.

To zoom a portion of the chart, select and drag mouse within the chart. Hold down shift key to pan zoomed chart.

#### 2 x 2 Table

|  | Cluster Time Period | |
| --- | --- | --- |
| Cluster Node | Inside | Outside |
| Inside | 431 | 4543 |
| Outside | 67205 | 975031 |
|  | 0.6% | 0.5% |

Close Chart Options

Show Chart Options

#### Chart Options

Title

Title can be changed by editing this text.

Series Inside Cluster

Observed

Expected

Observed / Expected

Percent Cases

Series Outside Cluster

Observed

Expected

Observed Chart Type

Histogram

Line

Switch the series type between line and histogram.

Cluster Band

Show Cluster Band

Band stretching across the plot area marking cluster interval.

To zoom a portion of the chart, select and drag mouse within the chart. Hold down shift key to pan zoomed chart.

#### 2 x 2 Table

|  | Cluster Time Period | |
| --- | --- | --- |
| Cluster Node | Inside | Outside |
| Inside | 82 | 585 |
| Outside | 67554 | 978989 |
|  | 0.1% | 0.1% |

Close Chart Options

Show Chart Options

#### Chart Options

Title

Title can be changed by editing this text.

Series Inside Cluster

Observed

Expected

Observed / Expected

Percent Cases

Series Outside Cluster

Observed

Expected

Observed Chart Type

Histogram

Line

Switch the series type between line and histogram.

Cluster Band

Show Cluster Band

Band stretching across the plot area marking cluster interval.

To zoom a portion of the chart, select and drag mouse within the chart. Hold down shift key to pan zoomed chart.

#### 2 x 2 Table

|  | Cluster Time Period | |
| --- | --- | --- |
| Cluster Node | Inside | Outside |
| Inside | 316 | 3205 |
| Outside | 67320 | 976369 |
|  | 0.5% | 0.3% |

Close Chart Options

Show Chart Options

#### Chart Options

Title

Title can be changed by editing this text.

Series Inside Cluster

Observed

Expected

Observed / Expected

Percent Cases

Series Outside Cluster

Observed

Expected

Observed Chart Type

Histogram

Line

Switch the series type between line and histogram.

Cluster Band

Show Cluster Band

Band stretching across the plot area marking cluster interval.

To zoom a portion of the chart, select and drag mouse within the chart. Hold down shift key to pan zoomed chart.

#### 2 x 2 Table

|  | Cluster Time Period | |
| --- | --- | --- |
| Cluster Node | Inside | Outside |
| Inside | 122 | 999 |
| Outside | 67514 | 978575 |
|  | 0.2% | 0.1% |

Close Chart Options

Show Chart Options

#### Chart Options

Title

Title can be changed by editing this text.

Series Inside Cluster

Observed

Expected

Observed / Expected

Percent Cases

Series Outside Cluster

Observed

Expected

Observed Chart Type

Histogram

Line

Switch the series type between line and histogram.

Cluster Band

Show Cluster Band

Band stretching across the plot area marking cluster interval.

To zoom a portion of the chart, select and drag mouse within the chart. Hold down shift key to pan zoomed chart.

#### 2 x 2 Table

|  | Cluster Time Period | |
| --- | --- | --- |
| Cluster Node | Inside | Outside |
| Inside | 1964 | 6952 |
| Outside | 201994 | 836300 |
|  | 1.0% | 0.8% |

Close Chart Options

Show Chart Options

#### Chart Options

Title

Title can be changed by editing this text.

Series Inside Cluster

Observed

Expected

Observed / Expected

Percent Cases

Series Outside Cluster

Observed

Expected

Observed Chart Type

Histogram

Line

Switch the series type between line and histogram.

Cluster Band

Show Cluster Band

Band stretching across the plot area marking cluster interval.

To zoom a portion of the chart, select and drag mouse within the chart. Hold down shift key to pan zoomed chart.

#### 2 x 2 Table

|  | Cluster Time Period | |
| --- | --- | --- |
| Cluster Node | Inside | Outside |
| Inside | 25 | 59 |
| Outside | 88990 | 958136 |
|  | 0.0% | 0.0% |

Close Chart Options

Show Chart Options

#### Chart Options

Title

Title can be changed by editing this text.

Series Inside Cluster

Observed

Expected

Observed / Expected

Percent Cases

Series Outside Cluster

Observed

Expected

Observed Chart Type

Histogram

Line

Switch the series type between line and histogram.

Cluster Band

Show Cluster Band

Band stretching across the plot area marking cluster interval.

To zoom a portion of the chart, select and drag mouse within the chart. Hold down shift key to pan zoomed chart.

#### 2 x 2 Table

|  | Cluster Time Period | |
| --- | --- | --- |
| Cluster Node | Inside | Outside |
| Inside | 69 | 485 |
| Outside | 67567 | 979089 |
|  | 0.1% | 0.0% |

Close Chart Options

Show Chart Options

#### Chart Options

Title

Title can be changed by editing this text.

Series Inside Cluster

Observed

Expected

Observed / Expected

Percent Cases

Series Outside Cluster

Observed

Expected

Observed Chart Type

Histogram

Line

Switch the series type between line and histogram.

Cluster Band

Show Cluster Band

Band stretching across the plot area marking cluster interval.

To zoom a portion of the chart, select and drag mouse within the chart. Hold down shift key to pan zoomed chart.

#### 2 x 2 Table

|  | Cluster Time Period | |
| --- | --- | --- |
| Cluster Node | Inside | Outside |
| Inside | 240 | 542 |
| Outside | 228877 | 817551 |
|  | 0.1% | 0.1% |

Close Chart Options

Show Chart Options

#### Chart Options

Title

Title can be changed by editing this text.

Series Inside Cluster

Observed

Expected

Observed / Expected

Percent Cases

Series Outside Cluster

Observed

Expected

Observed Chart Type

Histogram

Line

Switch the series type between line and histogram.

Cluster Band

Show Cluster Band

Band stretching across the plot area marking cluster interval.

To zoom a portion of the chart, select and drag mouse within the chart. Hold down shift key to pan zoomed chart.

#### 2 x 2 Table

|  | Cluster Time Period | |
| --- | --- | --- |
| Cluster Node | Inside | Outside |
| Inside | 882 | 6897 |
| Outside | 99832 | 939599 |
|  | 0.9% | 0.7% |

Close Chart Options

Show Chart Options

#### Chart Options

Title

Title can be changed by editing this text.

Series Inside Cluster

Observed

Expected

Observed / Expected

Percent Cases

Series Outside Cluster

Observed

Expected

Observed Chart Type

Histogram

Line

Switch the series type between line and histogram.

Cluster Band

Show Cluster Band

Band stretching across the plot area marking cluster interval.

To zoom a portion of the chart, select and drag mouse within the chart. Hold down shift key to pan zoomed chart.

#### 2 x 2 Table

|  | Cluster Time Period | |
| --- | --- | --- |
| Cluster Node | Inside | Outside |
| Inside | 262 | 2745 |
| Outside | 67374 | 976829 |
|  | 0.4% | 0.3% |

Close Chart Options

Show Chart Options

#### Chart Options

Title

Title can be changed by editing this text.

Series Inside Cluster

Observed

Expected

Observed / Expected

Percent Cases

Series Outside Cluster

Observed

Expected

Observed Chart Type

Histogram

Line

Switch the series type between line and histogram.

Cluster Band

Show Cluster Band

Band stretching across the plot area marking cluster interval.

To zoom a portion of the chart, select and drag mouse within the chart. Hold down shift key to pan zoomed chart.

#### 2 x 2 Table

|  | Cluster Time Period | |
| --- | --- | --- |
| Cluster Node | Inside | Outside |
| Inside | 252 | 2625 |
| Outside | 67384 | 976949 |
|  | 0.4% | 0.3% |

Close Chart Options

Show Chart Options

#### Chart Options

Title

Title can be changed by editing this text.

Series Inside Cluster

Observed

Expected

Observed / Expected

Percent Cases

Series Outside Cluster

Observed

Expected

Observed Chart Type

Histogram

Line

Switch the series type between line and histogram.

Cluster Band

Show Cluster Band

Band stretching across the plot area marking cluster interval.

To zoom a portion of the chart, select and drag mouse within the chart. Hold down shift key to pan zoomed chart.

#### 2 x 2 Table

|  | Cluster Time Period | |
| --- | --- | --- |
| Cluster Node | Inside | Outside |
| Inside | 193 | 542 |
| Outside | 191526 | 854949 |
|  | 0.1% | 0.1% |

Close Chart Options

Show Chart Options

#### Chart Options

Title

Title can be changed by editing this text.

Series Inside Cluster

Observed

Expected

Observed / Expected

Percent Cases

Series Outside Cluster

Observed

Expected

Observed Chart Type

Histogram

Line

Switch the series type between line and histogram.

Cluster Band

Show Cluster Band

Band stretching across the plot area marking cluster interval.

To zoom a portion of the chart, select and drag mouse within the chart. Hold down shift key to pan zoomed chart.

#### 2 x 2 Table

|  | Cluster Time Period | |
| --- | --- | --- |
| Cluster Node | Inside | Outside |
| Inside | 66 | 51 |
| Outside | 310012 | 737081 |
|  | 0.0% | 0.0% |

Close Chart Options

Show Chart Options

#### Chart Options

Title

Title can be changed by editing this text.

Series Inside Cluster

Observed

Expected

Observed / Expected

Percent Cases

Series Outside Cluster

Observed

Expected

Observed Chart Type

Histogram

Line

Switch the series type between line and histogram.

Cluster Band

Show Cluster Band

Band stretching across the plot area marking cluster interval.

To zoom a portion of the chart, select and drag mouse within the chart. Hold down shift key to pan zoomed chart.

#### 2 x 2 Table

|  | Cluster Time Period | |
| --- | --- | --- |
| Cluster Node | Inside | Outside |
| Inside | 245 | 572 |
| Outside | 228872 | 817521 |
|  | 0.1% | 0.1% |

Close Chart Options

Show Chart Options

#### Chart Options

Title

Title can be changed by editing this text.

Series Inside Cluster

Observed

Expected

Observed / Expected

Percent Cases

Series Outside Cluster

Observed

Expected

Observed Chart Type

Histogram

Line

Switch the series type between line and histogram.

Cluster Band

Show Cluster Band

Band stretching across the plot area marking cluster interval.

To zoom a portion of the chart, select and drag mouse within the chart. Hold down shift key to pan zoomed chart.

#### 2 x 2 Table

|  | Cluster Time Period | |
| --- | --- | --- |
| Cluster Node | Inside | Outside |
| Inside | 23 | 370 |
| Outside | 20495 | 1026322 |
|  | 0.1% | 0.0% |

Close Chart Options

Show Chart Options

#### Chart Options

Title

Title can be changed by editing this text.

Series Inside Cluster

Observed

Expected

Observed / Expected

Percent Cases

Series Outside Cluster

Observed

Expected

Observed Chart Type

Histogram

Line

Switch the series type between line and histogram.

Cluster Band

Show Cluster Band

Band stretching across the plot area marking cluster interval.

To zoom a portion of the chart, select and drag mouse within the chart. Hold down shift key to pan zoomed chart.

#### 2 x 2 Table

|  | Cluster Time Period | |
| --- | --- | --- |
| Cluster Node | Inside | Outside |
| Inside | 323 | 1423 |
| Outside | 148389 | 897075 |
|  | 0.2% | 0.2% |

Close Chart Options

Show Chart Options

#### Chart Options

Title

Title can be changed by editing this text.

Series Inside Cluster

Observed

Expected

Observed / Expected

Percent Cases

Series Outside Cluster

Observed

Expected

Observed Chart Type

Histogram

Line

Switch the series type between line and histogram.

Cluster Band

Show Cluster Band

Band stretching across the plot area marking cluster interval.

To zoom a portion of the chart, select and drag mouse within the chart. Hold down shift key to pan zoomed chart.

#### 2 x 2 Table

|  | Cluster Time Period | |
| --- | --- | --- |
| Cluster Node | Inside | Outside |
| Inside | 356 | 1482 |
| Outside | 158021 | 887351 |
|  | 0.2% | 0.2% |

Close Chart Options

Show Chart Options

#### Chart Options

Title

Title can be changed by editing this text.

Series Inside Cluster

Observed

Expected

Observed / Expected

Percent Cases

Series Outside Cluster

Observed

Expected

Observed Chart Type

Histogram

Line

Switch the series type between line and histogram.

Cluster Band

Show Cluster Band

Band stretching across the plot area marking cluster interval.

To zoom a portion of the chart, select and drag mouse within the chart. Hold down shift key to pan zoomed chart.

#### 2 x 2 Table

|  | Cluster Time Period | |
| --- | --- | --- |
| Cluster Node | Inside | Outside |
| Inside | 365 | 3051 |
| Outside | 88650 | 955144 |
|  | 0.4% | 0.3% |

Close Chart Options

Show Chart Options

#### Chart Options

Title

Title can be changed by editing this text.

Series Inside Cluster

Observed

Expected

Observed / Expected

Percent Cases

Series Outside Cluster

Observed

Expected

Observed Chart Type

Histogram

Line

Switch the series type between line and histogram.

Cluster Band

Show Cluster Band

Band stretching across the plot area marking cluster interval.

To zoom a portion of the chart, select and drag mouse within the chart. Hold down shift key to pan zoomed chart.

#### 2 x 2 Table

|  | Cluster Time Period | |
| --- | --- | --- |
| Cluster Node | Inside | Outside |
| Inside | 6 | 0 |
| Outside | 111923 | 935281 |
|  | 0.0% | 0.0% |

Close Chart Options

Show Chart Options

#### Chart Options

Title

Title can be changed by editing this text.

Series Inside Cluster

Observed

Expected

Observed / Expected

Percent Cases

Series Outside Cluster

Observed

Expected

Observed Chart Type

Histogram

Line

Switch the series type between line and histogram.

Cluster Band

Show Cluster Band

Band stretching across the plot area marking cluster interval.

To zoom a portion of the chart, select and drag mouse within the chart. Hold down shift key to pan zoomed chart.

#### 2 x 2 Table

|  | Cluster Time Period | |
| --- | --- | --- |
| Cluster Node | Inside | Outside |
| Inside | 73 | 286 |
| Outside | 123872 | 922979 |
|  | 0.1% | 0.0% |

Close Chart Options

Show Chart Options

#### Chart Options

Title

Title can be changed by editing this text.

Series Inside Cluster

Observed

Expected

Observed / Expected

Percent Cases

Series Outside Cluster

Observed

Expected

Observed Chart Type

Histogram

Line

Switch the series type between line and histogram.

Cluster Band

Show Cluster Band

Band stretching across the plot area marking cluster interval.

To zoom a portion of the chart, select and drag mouse within the chart. Hold down shift key to pan zoomed chart.

#### 2 x 2 Table

|  | Cluster Time Period | |
| --- | --- | --- |
| Cluster Node | Inside | Outside |
| Inside | 701 | 1688 |
| Outside | 260303 | 784518 |
|  | 0.3% | 0.2% |

Close Chart Options

Show Chart Options

#### Chart Options

Title

Title can be changed by editing this text.

Series Inside Cluster

Observed

Expected

Observed / Expected

Percent Cases

Series Outside Cluster

Observed

Expected

Observed Chart Type

Histogram

Line

Switch the series type between line and histogram.

Cluster Band

Show Cluster Band

Band stretching across the plot area marking cluster interval.

To zoom a portion of the chart, select and drag mouse within the chart. Hold down shift key to pan zoomed chart.

#### 2 x 2 Table

|  | Cluster Time Period | |
| --- | --- | --- |
| Cluster Node | Inside | Outside |
| Inside | 152 | 1123 |
| Outside | 88863 | 957072 |
|  | 0.2% | 0.1% |

Close Chart Options

Show Chart Options

#### Chart Options

Title

Title can be changed by editing this text.

Series Inside Cluster

Observed

Expected

Observed / Expected

Percent Cases

Series Outside Cluster

Observed

Expected

Observed Chart Type

Histogram

Line

Switch the series type between line and histogram.

Cluster Band

Show Cluster Band

Band stretching across the plot area marking cluster interval.

To zoom a portion of the chart, select and drag mouse within the chart. Hold down shift key to pan zoomed chart.

#### 2 x 2 Table

|  | Cluster Time Period | |
| --- | --- | --- |
| Cluster Node | Inside | Outside |
| Inside | 704 | 2388 |
| Outside | 203254 | 840864 |
|  | 0.3% | 0.3% |

Close Chart Options

Show Chart Options

#### Chart Options

Title

Title can be changed by editing this text.

Series Inside Cluster

Observed

Expected

Observed / Expected

Percent Cases

Series Outside Cluster

Observed

Expected

Observed Chart Type

Histogram

Line

Switch the series type between line and histogram.

Cluster Band

Show Cluster Band

Band stretching across the plot area marking cluster interval.

To zoom a portion of the chart, select and drag mouse within the chart. Hold down shift key to pan zoomed chart.

#### 2 x 2 Table

|  | Cluster Time Period | |
| --- | --- | --- |
| Cluster Node | Inside | Outside |
| Inside | 704 | 2390 |
| Outside | 203254 | 840862 |
|  | 0.3% | 0.3% |

Close Chart Options

Show Chart Options

#### Chart Options

Title

Title can be changed by editing this text.

Series Inside Cluster

Observed

Expected

Observed / Expected

Percent Cases

Series Outside Cluster

Observed

Expected

Observed Chart Type

Histogram

Line

Switch the series type between line and histogram.

Cluster Band

Show Cluster Band

Band stretching across the plot area marking cluster interval.

To zoom a portion of the chart, select and drag mouse within the chart. Hold down shift key to pan zoomed chart.

#### 2 x 2 Table

|  | Cluster Time Period | |
| --- | --- | --- |
| Cluster Node | Inside | Outside |
| Inside | 126 | 603 |
| Outside | 123819 | 922662 |
|  | 0.1% | 0.1% |

Close Chart Options

Show Chart Options

#### Chart Options

Title

Title can be changed by editing this text.

Series Inside Cluster

Observed

Expected

Observed / Expected

Percent Cases

Series Outside Cluster

Observed

Expected

Observed Chart Type

Histogram

Line

Switch the series type between line and histogram.

Cluster Band

Show Cluster Band

Band stretching across the plot area marking cluster interval.

To zoom a portion of the chart, select and drag mouse within the chart. Hold down shift key to pan zoomed chart.

#### 2 x 2 Table

|  | Cluster Time Period | |
| --- | --- | --- |
| Cluster Node | Inside | Outside |
| Inside | 14 | 223 |
| Outside | 20504 | 1026469 |
|  | 0.1% | 0.0% |

Close Chart Options

Show Chart Options

#### Chart Options

Title

Title can be changed by editing this text.

Series Inside Cluster

Observed

Expected

Observed / Expected

Percent Cases

Series Outside Cluster

Observed

Expected

Observed Chart Type

Histogram

Line

Switch the series type between line and histogram.

Cluster Band

Show Cluster Band

Band stretching across the plot area marking cluster interval.

To zoom a portion of the chart, select and drag mouse within the chart. Hold down shift key to pan zoomed chart.

#### 2 x 2 Table

|  | Cluster Time Period | |
| --- | --- | --- |
| Cluster Node | Inside | Outside |
| Inside | 32 | 115 |
| Outside | 111897 | 935166 |
|  | 0.0% | 0.0% |

Close Chart Options

Show Chart Options

#### Chart Options

Title

Title can be changed by editing this text.

Series Inside Cluster

Observed

Expected

Observed / Expected

Percent Cases

Series Outside Cluster

Observed

Expected

Observed Chart Type

Histogram

Line

Switch the series type between line and histogram.

Cluster Band

Show Cluster Band

Band stretching across the plot area marking cluster interval.

To zoom a portion of the chart, select and drag mouse within the chart. Hold down shift key to pan zoomed chart.

#### 2 x 2 Table

|  | Cluster Time Period | |
| --- | --- | --- |
| Cluster Node | Inside | Outside |
| Inside | 28 | 97 |
| Outside | 111901 | 935184 |
|  | 0.0% | 0.0% |

Close Chart Options

Show Chart Options

#### Chart Options

Title

Title can be changed by editing this text.

Series Inside Cluster

Observed

Expected

Observed / Expected

Percent Cases

Series Outside Cluster

Observed

Expected

Observed Chart Type

Histogram

Line

Switch the series type between line and histogram.

Cluster Band

Show Cluster Band

Band stretching across the plot area marking cluster interval.

To zoom a portion of the chart, select and drag mouse within the chart. Hold down shift key to pan zoomed chart.

#### 2 x 2 Table

|  | Cluster Time Period | |
| --- | --- | --- |
| Cluster Node | Inside | Outside |
| Inside | 8 | 45 |
| Outside | 31412 | 1015745 |
|  | 0.0% | 0.0% |

Close Chart Options

Show Chart Options

#### Chart Options

Title

Title can be changed by editing this text.

Series Inside Cluster

Observed

Expected

Observed / Expected

Percent Cases

Series Outside Cluster

Observed

Expected

Observed Chart Type

Histogram

Line

Switch the series type between line and histogram.

Cluster Band

Show Cluster Band

Band stretching across the plot area marking cluster interval.

To zoom a portion of the chart, select and drag mouse within the chart. Hold down shift key to pan zoomed chart.

#### 2 x 2 Table

|  | Cluster Time Period | |
| --- | --- | --- |
| Cluster Node | Inside | Outside |
| Inside | 25 | 117 |
| Outside | 88990 | 958078 |
|  | 0.0% | 0.0% |

Close Chart Options

Show Chart Options

#### Chart Options

Title

Title can be changed by editing this text.

Series Inside Cluster

Observed

Expected

Observed / Expected

Percent Cases

Series Outside Cluster

Observed

Expected

Observed Chart Type

Histogram

Line

Switch the series type between line and histogram.

Cluster Band

Show Cluster Band

Band stretching across the plot area marking cluster interval.

To zoom a portion of the chart, select and drag mouse within the chart. Hold down shift key to pan zoomed chart.

#### 2 x 2 Table

|  | Cluster Time Period | |
| --- | --- | --- |
| Cluster Node | Inside | Outside |
| Inside | 37 | 297 |
| Outside | 67599 | 979277 |
|  | 0.1% | 0.0% |

Close Chart Options

Show Chart Options

#### Chart Options

Title

Title can be changed by editing this text.

Series Inside Cluster

Observed

Expected

Observed / Expected

Percent Cases

Series Outside Cluster

Observed

Expected

Observed Chart Type

Histogram

Line

Switch the series type between line and histogram.

Cluster Band

Show Cluster Band

Band stretching across the plot area marking cluster interval.

To zoom a portion of the chart, select and drag mouse within the chart. Hold down shift key to pan zoomed chart.

#### 2 x 2 Table

|  | Cluster Time Period | |
| --- | --- | --- |
| Cluster Node | Inside | Outside |
| Inside | 3 | 0 |
| Outside | 67633 | 979574 |
|  | 0.0% | 0.0% |

Close Chart Options

Show Chart Options

#### Chart Options

Title

Title can be changed by editing this text.

Series Inside Cluster

Observed

Expected

Observed / Expected

Percent Cases

Series Outside Cluster

Observed

Expected

Observed Chart Type

Histogram

Line

Switch the series type between line and histogram.

Cluster Band

Show Cluster Band

Band stretching across the plot area marking cluster interval.

To zoom a portion of the chart, select and drag mouse within the chart. Hold down shift key to pan zoomed chart.

#### 2 x 2 Table

|  | Cluster Time Period | |
| --- | --- | --- |
| Cluster Node | Inside | Outside |
| Inside | 73 | 275 |
| Outside | 148639 | 898223 |
|  | 0.0% | 0.0% |

Close Chart Options

Show Chart Options

#### Chart Options

Title

Title can be changed by editing this text.

Series Inside Cluster

Observed

Expected

Observed / Expected

Percent Cases

Series Outside Cluster

Observed

Expected

Observed Chart Type

Histogram

Line

Switch the series type between line and histogram.

Cluster Band

Show Cluster Band

Band stretching across the plot area marking cluster interval.

To zoom a portion of the chart, select and drag mouse within the chart. Hold down shift key to pan zoomed chart.

#### 2 x 2 Table

|  | Cluster Time Period | |
| --- | --- | --- |
| Cluster Node | Inside | Outside |
| Inside | 6 | 2 |
| Outside | 158371 | 888831 |
|  | 0.0% | 0.0% |

Close Chart Options

Show Chart Options

#### Chart Options

Title

Title can be changed by editing this text.

Series Inside Cluster

Observed

Expected

Observed / Expected

Percent Cases

Series Outside Cluster

Observed

Expected

Observed Chart Type

Histogram

Line

Switch the series type between line and histogram.

Cluster Band

Show Cluster Band

Band stretching across the plot area marking cluster interval.

To zoom a portion of the chart, select and drag mouse within the chart. Hold down shift key to pan zoomed chart.

#### 2 x 2 Table

|  | Cluster Time Period | |
| --- | --- | --- |
| Cluster Node | Inside | Outside |
| Inside | 7 | 4 |
| Outside | 158370 | 888829 |
|  | 0.0% | 0.0% |

Close Chart Options

Show Chart Options

#### Chart Options

Title

Title can be changed by editing this text.

Series Inside Cluster

Observed

Expected

Observed / Expected

Percent Cases

Series Outside Cluster

Observed

Expected

Observed Chart Type

Histogram

Line

Switch the series type between line and histogram.

Cluster Band

Show Cluster Band

Band stretching across the plot area marking cluster interval.

To zoom a portion of the chart, select and drag mouse within the chart. Hold down shift key to pan zoomed chart.

#### 2 x 2 Table

|  | Cluster Time Period | |
| --- | --- | --- |
| Cluster Node | Inside | Outside |
| Inside | 101 | 1625 |
| Outside | 43213 | 1002271 |
|  | 0.2% | 0.2% |

Close Chart Options

Show Chart Options

#### Chart Options

Title

Title can be changed by editing this text.

Series Inside Cluster

Observed

Expected

Observed / Expected

Percent Cases

Series Outside Cluster

Observed

Expected

Observed Chart Type

Histogram

Line

Switch the series type between line and histogram.

Cluster Band

Show Cluster Band

Band stretching across the plot area marking cluster interval.

To zoom a portion of the chart, select and drag mouse within the chart. Hold down shift key to pan zoomed chart.

#### 2 x 2 Table

|  | Cluster Time Period | |
| --- | --- | --- |
| Cluster Node | Inside | Outside |
| Inside | 19 | 54 |
| Outside | 123926 | 923211 |
|  | 0.0% | 0.0% |

Close Chart Options

Show Chart Options

#### Chart Options

Title

Title can be changed by editing this text.

Series Inside Cluster

Observed

Expected

Observed / Expected

Percent Cases

Series Outside Cluster

Observed

Expected

Observed Chart Type

Histogram

Line

Switch the series type between line and histogram.

Cluster Band

Show Cluster Band

Band stretching across the plot area marking cluster interval.

To zoom a portion of the chart, select and drag mouse within the chart. Hold down shift key to pan zoomed chart.

#### 2 x 2 Table

|  | Cluster Time Period | |
| --- | --- | --- |
| Cluster Node | Inside | Outside |
| Inside | 28 | 128 |
| Outside | 100686 | 946368 |
|  | 0.0% | 0.0% |

Close Chart Options

Show Chart Options

#### Chart Options

Title

Title can be changed by editing this text.

Series Inside Cluster

Observed

Expected

Observed / Expected

Percent Cases

Series Outside Cluster

Observed

Expected

Observed Chart Type

Histogram

Line

Switch the series type between line and histogram.

Cluster Band

Show Cluster Band

Band stretching across the plot area marking cluster interval.

To zoom a portion of the chart, select and drag mouse within the chart. Hold down shift key to pan zoomed chart.

#### 2 x 2 Table

|  | Cluster Time Period | |
| --- | --- | --- |
| Cluster Node | Inside | Outside |
| Inside | 198 | 2868 |
| Outside | 54560 | 989584 |
|  | 0.4% | 0.3% |

Close Chart Options

Show Chart Options

#### Chart Options

Title

Title can be changed by editing this text.

Series Inside Cluster

Observed

Expected

Observed / Expected

Percent Cases

Series Outside Cluster

Observed

Expected

Observed Chart Type

Histogram

Line

Switch the series type between line and histogram.

Cluster Band

Show Cluster Band

Band stretching across the plot area marking cluster interval.

To zoom a portion of the chart, select and drag mouse within the chart. Hold down shift key to pan zoomed chart.

#### 2 x 2 Table

|  | Cluster Time Period | |
| --- | --- | --- |
| Cluster Node | Inside | Outside |
| Inside | 13 | 88 |
| Outside | 54745 | 992364 |
|  | 0.0% | 0.0% |

Close Chart Options

Show Chart Options

#### Chart Options

Title

Title can be changed by editing this text.

Series Inside Cluster

Observed

Expected

Observed / Expected

Percent Cases

Series Outside Cluster

Observed

Expected

Observed Chart Type

Histogram

Line

Switch the series type between line and histogram.

Cluster Band

Show Cluster Band

Band stretching across the plot area marking cluster interval.

To zoom a portion of the chart, select and drag mouse within the chart. Hold down shift key to pan zoomed chart.

#### 2 x 2 Table

|  | Cluster Time Period | |
| --- | --- | --- |
| Cluster Node | Inside | Outside |
| Inside | 6 | 70 |
| Outside | 20512 | 1026622 |
|  | 0.0% | 0.0% |

Close Chart Options

Show Chart Options

#### Chart Options

Title

Title can be changed by editing this text.

Series Inside Cluster

Observed

Expected

Observed / Expected

Percent Cases

Series Outside Cluster

Observed

Expected

Observed Chart Type

Histogram

Line

Switch the series type between line and histogram.

Cluster Band

Show Cluster Band

Band stretching across the plot area marking cluster interval.

To zoom a portion of the chart, select and drag mouse within the chart. Hold down shift key to pan zoomed chart.

#### 2 x 2 Table

|  | Cluster Time Period | |
| --- | --- | --- |
| Cluster Node | Inside | Outside |
| Inside | 164 | 1837 |
| Outside | 67472 | 977737 |
|  | 0.2% | 0.2% |

Close Chart Options

Show Chart Options

#### Chart Options

Title

Title can be changed by editing this text.

Series Inside Cluster

Observed

Expected

Observed / Expected

Percent Cases

Series Outside Cluster

Observed

Expected

Observed Chart Type

Histogram

Line

Switch the series type between line and histogram.

Cluster Band

Show Cluster Band

Band stretching across the plot area marking cluster interval.

To zoom a portion of the chart, select and drag mouse within the chart. Hold down shift key to pan zoomed chart.

#### 2 x 2 Table

|  | Cluster Time Period | |
| --- | --- | --- |
| Cluster Node | Inside | Outside |
| Inside | 173 | 551 |
| Outside | 203785 | 842701 |
|  | 0.1% | 0.1% |

Close Chart Options

Show Chart Options

#### Chart Options

Title

Title can be changed by editing this text.

Series Inside Cluster

Observed

Expected

Observed / Expected

Percent Cases

Series Outside Cluster

Observed

Expected

Observed Chart Type

Histogram

Line

Switch the series type between line and histogram.

Cluster Band

Show Cluster Band

Band stretching across the plot area marking cluster interval.

To zoom a portion of the chart, select and drag mouse within the chart. Hold down shift key to pan zoomed chart.

#### 2 x 2 Table

|  | Cluster Time Period | |
| --- | --- | --- |
| Cluster Node | Inside | Outside |
| Inside | 18 | 79 |
| Outside | 100696 | 946417 |
|  | 0.0% | 0.0% |

Close Chart Options

Show Chart Options

#### Chart Options

Title

Title can be changed by editing this text.

Series Inside Cluster

Observed

Expected

Observed / Expected

Percent Cases

Series Outside Cluster

Observed

Expected

Observed Chart Type

Histogram

Line

Switch the series type between line and histogram.

Cluster Band

Show Cluster Band

Band stretching across the plot area marking cluster interval.

To zoom a portion of the chart, select and drag mouse within the chart. Hold down shift key to pan zoomed chart.

#### 2 x 2 Table

|  | Cluster Time Period | |
| --- | --- | --- |
| Cluster Node | Inside | Outside |
| Inside | 16 | 142 |
| Outside | 54742 | 992310 |
|  | 0.0% | 0.0% |

Close Chart Options

Show Chart Options

#### Chart Options

Title

Title can be changed by editing this text.

Series Inside Cluster

Observed

Expected

Observed / Expected

Percent Cases

Series Outside Cluster

Observed

Expected

Observed Chart Type

Histogram

Line

Switch the series type between line and histogram.

Cluster Band

Show Cluster Band

Band stretching across the plot area marking cluster interval.

To zoom a portion of the chart, select and drag mouse within the chart. Hold down shift key to pan zoomed chart.

#### 2 x 2 Table

|  | Cluster Time Period | |
| --- | --- | --- |
| Cluster Node | Inside | Outside |
| Inside | 6 | 5 |
| Outside | 158371 | 888828 |
|  | 0.0% | 0.0% |

Close Chart Options

Show Chart Options

#### Chart Options

Title

Title can be changed by editing this text.

Series Inside Cluster

Observed

Expected

Observed / Expected

Percent Cases

Series Outside Cluster

Observed

Expected

Observed Chart Type

Histogram

Line

Switch the series type between line and histogram.

Cluster Band

Show Cluster Band

Band stretching across the plot area marking cluster interval.

To zoom a portion of the chart, select and drag mouse within the chart. Hold down shift key to pan zoomed chart.

#### 2 x 2 Table

|  | Cluster Time Period | |
| --- | --- | --- |
| Cluster Node | Inside | Outside |
| Inside | 174 | 561 |
| Outside | 203784 | 842691 |
|  | 0.1% | 0.1% |

Close Chart Options

Show Chart Options

#### Chart Options

Title

Title can be changed by editing this text.

Series Inside Cluster

Observed

Expected

Observed / Expected

Percent Cases

Series Outside Cluster

Observed

Expected

Observed Chart Type

Histogram

Line

Switch the series type between line and histogram.

Cluster Band

Show Cluster Band

Band stretching across the plot area marking cluster interval.

To zoom a portion of the chart, select and drag mouse within the chart. Hold down shift key to pan zoomed chart.

#### 2 x 2 Table

|  | Cluster Time Period | |
| --- | --- | --- |
| Cluster Node | Inside | Outside |
| Inside | 197 | 1519 |
| Outside | 100517 | 944977 |
|  | 0.2% | 0.2% |

Close Chart Options

Show Chart Options

#### Chart Options

Title

Title can be changed by editing this text.

Series Inside Cluster

Observed

Expected

Observed / Expected

Percent Cases

Series Outside Cluster

Observed

Expected

Observed Chart Type

Histogram

Line

Switch the series type between line and histogram.

Cluster Band

Show Cluster Band

Band stretching across the plot area marking cluster interval.

To zoom a portion of the chart, select and drag mouse within the chart. Hold down shift key to pan zoomed chart.

#### 2 x 2 Table

|  | Cluster Time Period | |
| --- | --- | --- |
| Cluster Node | Inside | Outside |
| Inside | 57 | 695 |
| Outside | 54701 | 991757 |
|  | 0.1% | 0.1% |

Close Chart Options

Show Chart Options

#### Chart Options

Title

Title can be changed by editing this text.

Series Inside Cluster

Observed

Expected

Observed / Expected

Percent Cases

Series Outside Cluster

Observed

Expected

Observed Chart Type

Histogram

Line

Switch the series type between line and histogram.

Cluster Band

Show Cluster Band

Band stretching across the plot area marking cluster interval.

To zoom a portion of the chart, select and drag mouse within the chart. Hold down shift key to pan zoomed chart.

#### 2 x 2 Table

|  | Cluster Time Period | |
| --- | --- | --- |
| Cluster Node | Inside | Outside |
| Inside | 96 | 542 |
| Outside | 123849 | 922723 |
|  | 0.1% | 0.1% |

Close Chart Options

Show Chart Options

#### Chart Options

Title

Title can be changed by editing this text.

Series Inside Cluster

Observed

Expected

Observed / Expected

Percent Cases

Series Outside Cluster

Observed

Expected

Observed Chart Type

Histogram

Line

Switch the series type between line and histogram.

Cluster Band

Show Cluster Band

Band stretching across the plot area marking cluster interval.

To zoom a portion of the chart, select and drag mouse within the chart. Hold down shift key to pan zoomed chart.

#### 2 x 2 Table

|  | Cluster Time Period | |
| --- | --- | --- |
| Cluster Node | Inside | Outside |
| Inside | 7 | 21 |
| Outside | 89008 | 958174 |
|  | 0.0% | 0.0% |

Close Chart Options

Show Chart Options

#### Chart Options

Title

Title can be changed by editing this text.

Series Inside Cluster

Observed

Expected

Observed / Expected

Percent Cases

Series Outside Cluster

Observed

Expected

Observed Chart Type

Histogram

Line

Switch the series type between line and histogram.

Cluster Band

Show Cluster Band

Band stretching across the plot area marking cluster interval.

To zoom a portion of the chart, select and drag mouse within the chart. Hold down shift key to pan zoomed chart.

#### 2 x 2 Table

|  | Cluster Time Period | |
| --- | --- | --- |
| Cluster Node | Inside | Outside |
| Inside | 45 | 1547 |
| Outside | 20473 | 1025145 |
|  | 0.2% | 0.2% |

Close Chart Options

Show Chart Options

#### Chart Options

Title

Title can be changed by editing this text.

Series Inside Cluster

Observed

Expected

Observed / Expected

Percent Cases

Series Outside Cluster

Observed

Expected

Observed Chart Type

Histogram

Line

Switch the series type between line and histogram.

Cluster Band

Show Cluster Band

Band stretching across the plot area marking cluster interval.

To zoom a portion of the chart, select and drag mouse within the chart. Hold down shift key to pan zoomed chart.

#### 2 x 2 Table

|  | Cluster Time Period | |
| --- | --- | --- |
| Cluster Node | Inside | Outside |
| Inside | 109 | 632 |
| Outside | 123836 | 922633 |
|  | 0.1% | 0.1% |

Close Chart Options

Show Chart Options

#### Chart Options

Title

Title can be changed by editing this text.

Series Inside Cluster

Observed

Expected

Observed / Expected

Percent Cases

Series Outside Cluster

Observed

Expected

Observed Chart Type

Histogram

Line

Switch the series type between line and histogram.

Cluster Band

Show Cluster Band

Band stretching across the plot area marking cluster interval.

To zoom a portion of the chart, select and drag mouse within the chart. Hold down shift key to pan zoomed chart.

#### 2 x 2 Table

|  | Cluster Time Period | |
| --- | --- | --- |
| Cluster Node | Inside | Outside |
| Inside | 202 | 2377 |
| Outside | 67434 | 977197 |
|  | 0.3% | 0.2% |

Close Chart Options

Show Chart Options

#### Chart Options

Title

Title can be changed by editing this text.

Series Inside Cluster

Observed

Expected

Observed / Expected

Percent Cases

Series Outside Cluster

Observed

Expected

Observed Chart Type

Histogram

Line

Switch the series type between line and histogram.

Cluster Band

Show Cluster Band

Band stretching across the plot area marking cluster interval.

To zoom a portion of the chart, select and drag mouse within the chart. Hold down shift key to pan zoomed chart.

#### 2 x 2 Table

|  | Cluster Time Period | |
| --- | --- | --- |
| Cluster Node | Inside | Outside |
| Inside | 9 | 62 |
| Outside | 54749 | 992390 |
|  | 0.0% | 0.0% |

Close Chart Options

Show Chart Options

#### Chart Options

Title

Title can be changed by editing this text.

Series Inside Cluster

Observed

Expected

Observed / Expected

Percent Cases

Series Outside Cluster

Observed

Expected

Observed Chart Type

Histogram

Line

Switch the series type between line and histogram.

Cluster Band

Show Cluster Band

Band stretching across the plot area marking cluster interval.

To zoom a portion of the chart, select and drag mouse within the chart. Hold down shift key to pan zoomed chart.

#### 2 x 2 Table

|  | Cluster Time Period | |
| --- | --- | --- |
| Cluster Node | Inside | Outside |
| Inside | 15 | 170 |
| Outside | 43299 | 1003726 |
|  | 0.0% | 0.0% |

Close Chart Options

Show Chart Options

#### Chart Options

Title

Title can be changed by editing this text.

Series Inside Cluster

Observed

Expected

Observed / Expected

Percent Cases

Series Outside Cluster

Observed

Expected

Observed Chart Type

Histogram

Line

Switch the series type between line and histogram.

Cluster Band

Show Cluster Band

Band stretching across the plot area marking cluster interval.

To zoom a portion of the chart, select and drag mouse within the chart. Hold down shift key to pan zoomed chart.

#### 2 x 2 Table

|  | Cluster Time Period | |
| --- | --- | --- |
| Cluster Node | Inside | Outside |
| Inside | 18 | 1101 |
| Outside | 9635 | 1036456 |
|  | 0.2% | 0.1% |

Close Chart Options

Show Chart Options

#### Chart Options

Title

Title can be changed by editing this text.

Series Inside Cluster

Observed

Expected

Observed / Expected

Percent Cases

Series Outside Cluster

Observed

Expected

Observed Chart Type

Histogram

Line

Switch the series type between line and histogram.

Cluster Band

Show Cluster Band

Band stretching across the plot area marking cluster interval.

To zoom a portion of the chart, select and drag mouse within the chart. Hold down shift key to pan zoomed chart.

#### 2 x 2 Table

|  | Cluster Time Period | |
| --- | --- | --- |
| Cluster Node | Inside | Outside |
| Inside | 11 | 7 |
| Outside | 296462 | 750730 |
|  | 0.0% | 0.0% |

Close Chart Options

Show Chart Options

#### Chart Options

Title

Title can be changed by editing this text.

Series Inside Cluster

Observed

Expected

Observed / Expected

Percent Cases

Series Outside Cluster

Observed

Expected

Observed Chart Type

Histogram

Line

Switch the series type between line and histogram.

Cluster Band

Show Cluster Band

Band stretching across the plot area marking cluster interval.

To zoom a portion of the chart, select and drag mouse within the chart. Hold down shift key to pan zoomed chart.

#### 2 x 2 Table

|  | Cluster Time Period | |
| --- | --- | --- |
| Cluster Node | Inside | Outside |
| Inside | 20 | 217 |
| Outside | 54738 | 992235 |
|  | 0.0% | 0.0% |

Close Chart Options

Show Chart Options

#### Chart Options

Title

Title can be changed by editing this text.

Series Inside Cluster

Observed

Expected

Observed / Expected

Percent Cases

Series Outside Cluster

Observed

Expected

Observed Chart Type

Histogram

Line

Switch the series type between line and histogram.

Cluster Band

Show Cluster Band

Band stretching across the plot area marking cluster interval.

To zoom a portion of the chart, select and drag mouse within the chart. Hold down shift key to pan zoomed chart.

#### 2 x 2 Table

|  | Cluster Time Period | |
| --- | --- | --- |
| Cluster Node | Inside | Outside |
| Inside | 487 | 4143 |
| Outside | 100227 | 942353 |
|  | 0.5% | 0.4% |

Close Chart Options

Show Chart Options

#### Chart Options

Title

Title can be changed by editing this text.

Series Inside Cluster

Observed

Expected

Observed / Expected

Percent Cases

Series Outside Cluster

Observed

Expected

Observed Chart Type

Histogram

Line

Switch the series type between line and histogram.

Cluster Band

Show Cluster Band

Band stretching across the plot area marking cluster interval.

To zoom a portion of the chart, select and drag mouse within the chart. Hold down shift key to pan zoomed chart.

#### 2 x 2 Table

|  | Cluster Time Period | |
| --- | --- | --- |
| Cluster Node | Inside | Outside |
| Inside | 32 | 646 |
| Outside | 31388 | 1015144 |
|  | 0.1% | 0.1% |

Close Chart Options

Show Chart Options

#### Chart Options

Title

Title can be changed by editing this text.

Series Inside Cluster

Observed

Expected

Observed / Expected

Percent Cases

Series Outside Cluster

Observed

Expected

Observed Chart Type

Histogram

Line

Switch the series type between line and histogram.

Cluster Band

Show Cluster Band

Band stretching across the plot area marking cluster interval.

To zoom a portion of the chart, select and drag mouse within the chart. Hold down shift key to pan zoomed chart.

#### 2 x 2 Table

|  | Cluster Time Period | |
| --- | --- | --- |
| Cluster Node | Inside | Outside |
| Inside | 13 | 14 |
| Outside | 260991 | 786192 |
|  | 0.0% | 0.0% |

Close Chart Options

Show Chart Options

#### Chart Options

Title

Title can be changed by editing this text.

Series Inside Cluster

Observed

Expected

Observed / Expected

Percent Cases

Series Outside Cluster

Observed

Expected

Observed Chart Type

Histogram

Line

Switch the series type between line and histogram.

Cluster Band

Show Cluster Band

Band stretching across the plot area marking cluster interval.

To zoom a portion of the chart, select and drag mouse within the chart. Hold down shift key to pan zoomed chart.

#### 2 x 2 Table

|  | Cluster Time Period | |
| --- | --- | --- |
| Cluster Node | Inside | Outside |
| Inside | 6 | 21 |
| Outside | 89009 | 958174 |
|  | 0.0% | 0.0% |

Close Chart Options

Show Chart Options

#### Chart Options

Title

Title can be changed by editing this text.

Series Inside Cluster

Observed

Expected

Observed / Expected

Percent Cases

Series Outside Cluster

Observed

Expected

Observed Chart Type

Histogram

Line

Switch the series type between line and histogram.

Cluster Band

Show Cluster Band

Band stretching across the plot area marking cluster interval.

To zoom a portion of the chart, select and drag mouse within the chart. Hold down shift key to pan zoomed chart.

#### 2 x 2 Table

|  | Cluster Time Period | |
| --- | --- | --- |
| Cluster Node | Inside | Outside |
| Inside | 122 | 2225 |
| Outside | 43192 | 1001671 |
|  | 0.3% | 0.2% |

Close Chart Options

Show Chart Options

#### Chart Options

Title

Title can be changed by editing this text.

Series Inside Cluster

Observed

Expected

Observed / Expected

Percent Cases

Series Outside Cluster

Observed

Expected

Observed Chart Type

Histogram

Line

Switch the series type between line and histogram.

Cluster Band

Show Cluster Band

Band stretching across the plot area marking cluster interval.

To zoom a portion of the chart, select and drag mouse within the chart. Hold down shift key to pan zoomed chart.

#### 2 x 2 Table

|  | Cluster Time Period | |
| --- | --- | --- |
| Cluster Node | Inside | Outside |
| Inside | 4 | 4 |
| Outside | 148708 | 898494 |
|  | 0.0% | 0.0% |

Close Chart Options

Generated with TreeScan v2.3
